# Widely accessible prognostication using medical history for fetal growth restriction and small for gestational age in nationwide insured women

**DOI:** 10.1101/2024.01.08.24300958

**Authors:** Herdiantri Sufriyana, Fariska Zata Amani, Aufar Zimamuz Zaman Al Hajiri, Yu-Wei Wu, Emily Chia-Yu Su

## Abstract

**Objectives:** Prevention of fetal growth restriction/small for gestational age is adequate if screening is accurate. Ultrasound and biomarkers can achieve this goal; however, both are often inaccessible. This study aimed to develop, validate, and deploy a prognostic prediction model for screening fetal growth restriction/small for gestational age using only medical history.

**Methods:** From a nationwide health insurance database (*n*=1,697,452), we retrospectively selected visits of 12-to-55-year-old females to 22,024 healthcare providers of primary, secondary, and tertiary care. This study used machine learning (including deep learning) to develop prediction models using 54 medical-history predictors. After evaluating model calibration, clinical utility, and explainability, we selected the best by discrimination ability. We also externally validated and compared the models with those from previous studies, which were rigorously selected by a systematic review of Pubmed, Scopus, and Web of Science.

**Results:** We selected 169,746 subjects with 507,319 visits for predictive modeling. The best prediction model was a deep-insight visible neural network. It had an area under the receiver operating characteristics curve of 0.742 (95% confidence interval 0.734 to 0.750) and a sensitivity of 49.09% (95% confidence interval 47.60% to 50.58% using a threshold with 95% specificity). The model was competitive against the previous models in a systematic review of 30 eligible studies of 381 records, including those using either ultrasound or biomarker measurements. We deployed a web application to apply the model.

**Conclusions:** Our model used only medical history to improve accessibility for fetal growth restriction/small for gestational age screening. However, future studies are warranted to evaluate if this model’s usage impacts patient outcomes.

## Introduction

Fetal growth restriction (FGR) and small for gestational age (SGA) are two terms of a single condition with the same diagnostic criterion in principle but different measures in practice.^1^ This condition is the second leading cause of preventable perinatal deaths.^2^ The prevention method depends on FGR/SGA predictions with a clinically acceptable predictive performance.^3^ However, most settings lack accessibility to predictors in existing prediction models.^4^

A pregnancy with FGR likely results in delivering low-birth-weight infants,^5^ an indirect cause of neonatal deaths.^6–8^ Neonatal mortality rates varied from 20 to 30 deaths per 1000 live births worldwide in 2013.^9^ Low-birth-weight infants also need to spend time in a neonatal intensive care unit.^10^ But, this requires high costs and is a limited resource in many countries.^11,12^ Prevention of FGR/SGA may reduce neonatal mortality and associated costs.^13^ Several preventive strategies were found to be effective for FGR/SGA;^14^ yet, this intervention needs a screening method with a good predictive performance.^3^

Since a low-cost method such as symphysis fundal height was not recommended by a Cochrane review, mainly due to low sensitivity (∼17%), there is a trend to employ either ultrasound or biomarker measurements for FGR/SGA screening.^15^ Nonetheless, these methods are inaccessible in resource-limited settings.^15,16^ Meanwhile, there was an association detected of FGR/SGA with a woman’s medical history.^17^ Because a health insurance claim database abundantly records medical histories, this allows proactive screening for FGR/SGA, particularly in countries with universal health coverage.^18^ Screening by medical history is also independent of the number of pregnancy consultations on which FGR detection depends (hazard ratio 1.15, 95% confidence interval [CI] 1.05 to 1.26).^19^ However, studies have yet to develop a screening method for FGR/SGA using only medical history.

Prognostic predictions of FGR/SGA using medical histories can be either a prediction model for use in resource-limited settings or a preliminary prediction model before ordering ultrasound and biomarker measurements. Both statistical and computational machine learning can predict pregnancy outcomes in advance,^20^ including deep learning and those using only medical history.^20,21^ We aimed to develop, validate, and deploy a prognostic prediction model for screening FGR/SGA using only medical history in nationwide insured women.

## Materials and Methods

Report completeness of this study was according to the transparent reporting of a multivariable prediction model for individual prognosis or diagnosis (TRIPOD) checklist (Appendix A).^22^ We followed a protocol with the same software and hardware (Tables B1-B3, C1),^23^ except those stated otherwise. This study was under a single project that compared a deep-insight visible neural network (DI-VNN) to other machine learning algorithms to predict several outcomes in medicine. The Taipei Medical University Joint Institutional Review Board exempted this project from the ethical review (TMU-JIRB no.: N202106025).

### Study design and data source

We applied a retrospective design to select subjects from a public dataset version 2 (August 2019;^24^ access approval no.: 510/PPID/1223) of a nationwide health insurance database in Indonesia. The dataset was a cross-sectional, random sampling of ∼1% of insurance holders within 2 years up to 2016. This sampling included all affiliated healthcare providers (*n*=22,024) at all levels (i.e., primary, secondary, and tertiary care).

The inclusion criteria were females aged 12 to 55 years who had visited primary, secondary, or tertiary care facilities. All visits afterward were exprotocolcluded if a woman was pregnant and had a delivery. If a woman became pregnant twice within the dataset period, then different identifiers were assigned to differentiate the pregnancy periods of that woman. To determine a delivery, we used several codes of diagnoses and procedures (Table C2).

This study developed a prediction model for detecting in advance a visit by a subject who would be diagnosed with either FGR or SGA. We pursued to achieve an acceptable sensitivity at 95% specificity but using more-accessible predictors. Nevertheless, we compared our prediction models with those from previous studies selected by systematic review methods to evaluate if our predictive modeling was successful. Since there were different policies in choosing a prediction threshold (e.g., that at 90% vs. 95% specificity), the comparison was conducted using receiver operating characteristics (ROC) curves and the area under the ROC curve (AUROC).

The event outcome definition in this study utilized the International Classification of Disease version 10 (ICD-10) codes. These were codes preceded by either O365 (maternal care for known or suspected fetal growth) or P05 (disorders of newborns related to slow fetal growth and fetal malnutrition). Both codes indicating FGR and SGA were assigned with those respectively for mothers and fetuses/newborns. A nonevent outcome was assigned if the end of pregnancy was identified within the dataset period by the codes for determining delivery. Otherwise, we assigned an outcome to a censored one.

Candidate predictors were only medical histories of diagnoses and procedures. These were either single or multiple ICD-10 codes. As extensively described in the protocol,^22^ the preprocessing of candidate predictors consisted of (1) preventing zero variance, perfect separation and leakage of the outcome, and redundant predictors; (2) simulating real-world data; and (3) systematically determining the multiple ICD-10 codes for defining latent candidate predictors based on prior knowledge. After this preprocessing (Tables C3-C7), we identified 54 candidate predictors, including four latent candidate predictors of multiple pregnancies, varicella, urinary tract infections, and placenta previa.

### Statistical analysis

We developed five models using different algorithms and hyperparameter tuning, as described in the protocol.^22^ The first applied ridge regression (RR). The second to fourth models used 54 candidate predictors transformed into principal components (PCs). We applied three algorithms using these PCs: (1) elastic net regression (PC-ENR); (2) random forest (PC-RF); and (3) gradient boosting machine (PC-GBM). The fifth model was a deep-insight visible neural network (DI-VNN). However, unlike the protocol,^22^ we did not limit this model to only 22 of 54 candidate predictors, which had a false discovery rate of ≤5% based on differential analyses with Benjamini-Hochberg multiple testing corrections. Instead, we used all 54 candidate predictors considering the feasibility of constructing the data-driven network architecture. In addition, all model recalibration was by either a logistic regression or a general additive model using locally weighted scatterplot smoothing. The recalibration procedure also differed from the protocol.^22^ This is because the models only sometimes resulted in a wide range of predicted probabilities, as required for recalibration. Unlike the protocol, we chose 100 repetitions for bootstrapping, considering the sample size of this study compared to that of the protocol. Details on model development and validation are described in Table B2.

For deployment, this model will predict the outcome each time an insured woman visits a healthcare provider. We provided the best model in this study as a web application. A user is only required to upload a comma-separated value (.csv) file consisting a two-column table. It includes column headers of “admission_date” (yyyy-mm-dd) and “code” (ICD-10 code at discharge) from previous to current visits.

We computed an uncertainty interval (i.e., 95% confidence interval, CI) for each evaluation metric. This interval inference used subsets of an evaluated set, resampled by bootstrapping and cross-validation. All analytical codes were publicly shared (see “Data sharing statement”).

The selection of latent candidate predictors in the first model applied inverse probability weighting for the multivariate analyses, according to the protocol.^22^ Results were also compared to those by outcome regression. We selected a latent candidate predictor if its association with the outcome had an interval of odds ratio (OR) excluding a value of 1.

The evaluation metrics were those for assessing the models’ calibration, utility, explainability, and discrimination. To evaluate the model calibration, we assessed (1) a calibration plot with a regression line and histograms of either event or nonevent distribution of the predicted probabilities; (2) the intercept and slope of the linear regression; and (3) the Brier score. We measured the clinical utility using a decision curve analysis by comparing the net benefits of a model with those if we treated all predictions as either positive (i.e., treat all) or negative (i.e., treat none). Clinicians (i.e., FZA and AZZAH) assessed the explainability. They were given counterfactual quantities for each predictor in a model.^25^ These consisted of the probability of necessity (PN; Equation 1) and the probability of sufficiency (PS; Equation 2). Eventually, we evaluated the discrimination ability of well-calibrated models by the ROC curve and sensitivity at 95% specificity.

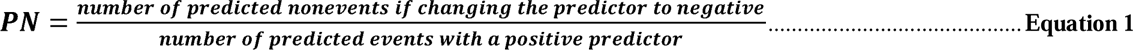

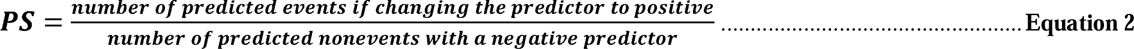

Furthermore, we compared our models with previous ones identified by a systematic review and meta-analysis (see “Comparison to previous models”). We compared the best model with those from previous studies. These were identified by following 11 of 14 items in section methods of the preferred reporting items for systematic reviews and meta-analyses (PRISMA)-extended checklist statements.^26^ Those items are described in Table B4.

## Results

### Subject characteristics

From the database (*n*=1,697,452), we selected 12-to-55-year-old females (*n*=169,746) that had visited (*n*=507,319) primary, secondary, or tertiary care (Figure 1). After removing subjects with no pregnancy and their visits, we split the selected data for internal and external validation. There were no overlapped visits between the internal and external validation sets. We only used the former to develop the prediction models in this study, including association tests to select candidate predictors.

**Figure 1.**
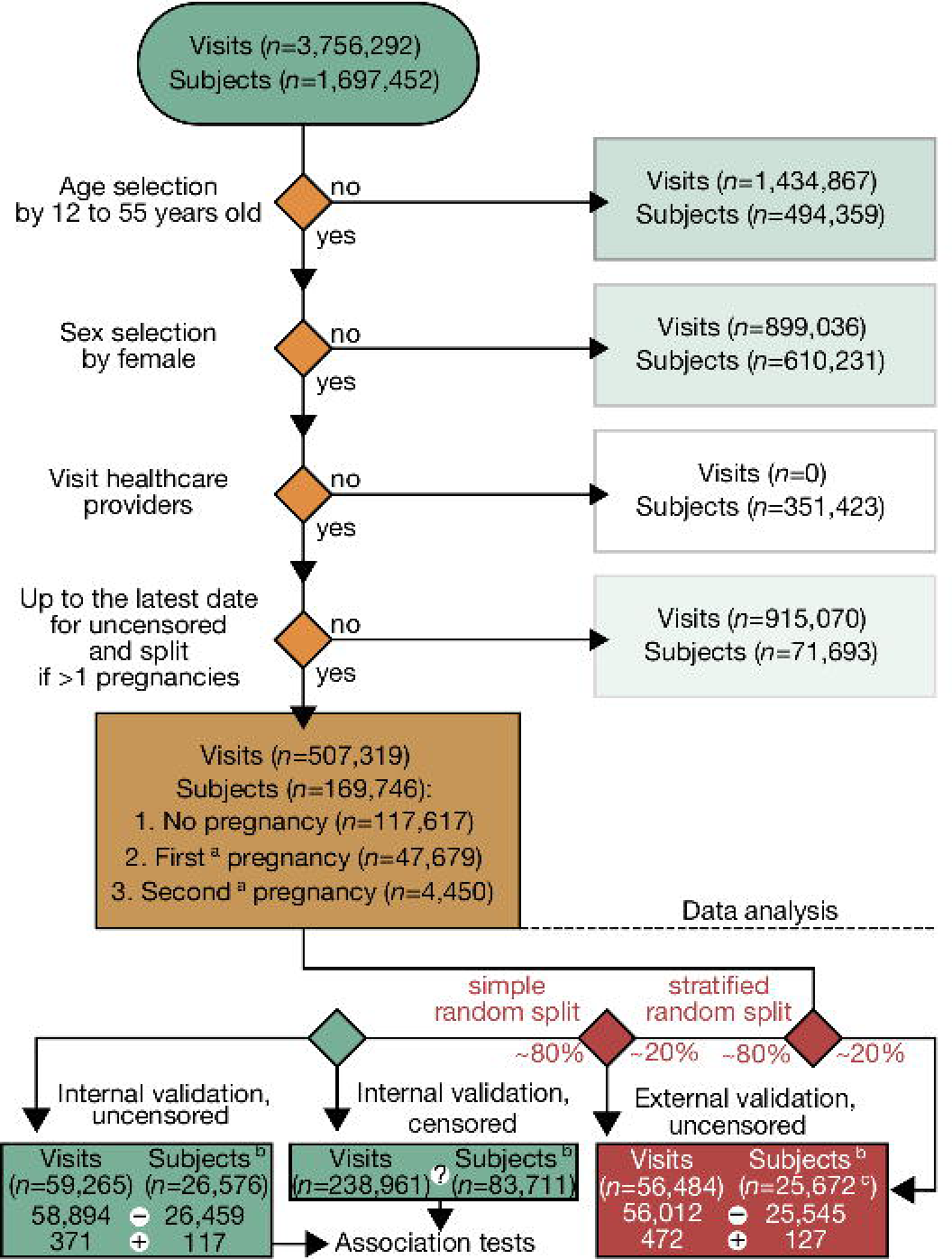
Subject selection by applying a retrospective design and data partitioning for internal and external validations. The set for association tests included censored outcomes. The summation of the internal and external validation numbers differs from the total because: (1) there were subject overlaps; (2) the numbers of subjects and visits in the censored internal validation are not shown; and (3) we excluded subjects with no pregnancy before data analysis. ^a^, the first and second pregnancies of a subject within the database period, not parity; ^b^, subjects per pregnancy episode; ^c^, only subjects in the external random split overlapped with those in the internal validation sets; *n*, sample size; (?), number of censoring; (–), number of nonevents; (+), number of events.

To characterize subjects in the internal validation set (Table 1), we also included subjects with uncensored outcomes (*n*=26,576). There were differences between subjects without and those with FGR/SGA based on multiple univariate analyses. These were in terms of subject characteristics, i.e.: (1) maternal age; (2) third vs. first categories of the insurance class; (3) single vs. married categories of the marital status; and (4) private company vs. central-government employee categories of the occupation segment of the householder. We also identified differences in terms of latent candidate predictors. Two of these variables were the risk of adverse pregnancy by maternal age and a low socioeconomic status. The former represented maternal age of either <20 or >25 years, while the latter represented either the third insurance class or unemployed householder (Table C7). Differences in the latent candidate predictors implied their associations with the outcome.

**Table 1.** Subject characteristics for association tests and internal validation set.

| Variable |  | Not FGR/SGA <sup>a</sup><br>(n=26,459) | FGR/SGA <sup>a</sup><br>(n=117) | p value |
| --- | --- | --- | --- | --- |
| Pregnancy episode within database period <sup>b</sup> | First pregnancy, <sup>c</sup> no. (%) | 25,096 (94.85) | 109 (93.16) | (reference) |
|  | Second pregnancy, <sup>c</sup> no. (%) | 1363 (5.15) | 8 (6.84) | 0.41 |
| Maternal age | Mean (SD), year | 29 (6) | 28 (6) | 0.006** |
| Insurance class | First, no. (%) | 3604 (13.62) | 21 (17.95) | (reference) |
|  | Unspecified, no. (%) | 87 ( 0.33) | 1 ( 0.85) | 0.51 |
|  | Second, no. (%) | 9226 (34.87) | 50 (42.74) | 0.78 |
|  | Third, no. (%) | 13,542 (51.18) | 45 (38.46) | 0.03* |
| Marital status | Married, no. (%) | 16,831 (63.61) | 77 (66) | (reference) |
|  | Single, no. (%) | 2397 ( 9.06) | 20 (17) | 0.02* |
|  | Unspecified, no. (%) | 7117 (26.90) | 20 (17) | 0.05 |
|  | Divorced/widowed, no. (%) | 114 ( 0.43) | 77 (66) | 0.97 |
| Occupation segment of the householder | Central-government employee, no. (%) | 7683 (29.04) | 20 (17.1) | (reference) |
|  | Private company employee, no. (%) | 9611 (36.32) | 57 (48.7) | 0.002** |
|  | Private company employer or self-employed, no. (%) | 7871 (29.75) | 35 (29.9) | 0.06 |
|  | Local-government employee, no. (%) | 1278 ( 4.83) | 5 ( 4.3) | 0.42 |
|  | Unemployed, no. (%) | 16 ( 0.06) | 5 ( 4.3) | 0.98 |
| Pregnancy-induced hypertension | Negative, no. (%) | 25,366 (9.6e-01) | 98 (8.4e-01) | (reference) |
|  | Positive, no. (%) | 1093 (4.1e-02) | 19 (1.6e-01) | <0.001*** |
| Multiple pregnancies | Negative, no. (%) | 26,271 (9.9e-01) | 112 (9.6e-01) | (reference) |
|  | Positive, no. (%) | 188 (7.1e-03) | 5 (4.3e-02) | <0.001*** |
| Malaria | Negative, no. (%) | 26,439 (1.0e+00) | 117 (1.0e+00) | (reference) |
|  | Positive, no. (%) | 20 (7.6e-04) | 0 (0.0e+00) | <0.001*** |
| Varicella | Negative, no. (%) | 26,446 (1.0e+00) | 117 (1.0e+00) | (reference) |
|  | Positive, no. (%) | 13 (4.9e-04) | 0 (0.0e+00) | <0.001*** |
| Risk of adverse pregnancy by maternal age | Negative, no. (%) | 19,660 (7.4e-01) | 93 (7.9e-01) | (reference) |
|  | Positive, no. (%) | 6799 (2.6e-01) | 24 (2.1e-01) | <0.001*** |
| Urinary tract infection | Negative, no. (%) | 26,294 (9.9e-01) | 116 (9.9e-01) | (reference) |
|  | Positive, no. (%) | 165 (6.2e-03) | 1 (8.5e-03) | <0.001*** |
| Placenta previa | Negative, no. (%) | 26,187 (9.9e-01) | 113 (9.7e-01) | (reference) |
|  | Positive, no. (%) | 272 (1.0e-02) | 4 (3.4e-02) | <0.001*** |
| Low socioeconomic status | Negative, no. (%) | 12,901 (4.9e-01) | 72 (6.2e-01) | (reference) |
|  | Positive, no. (%) | 13,558 (5.1e-01) | 45 (3.8e-01) | 0.05* |

### Association tests

To select latent candidate predictors in the prediction models, their associations with the outcome were verified by multivariate analyses using inverse probability weighting (see Table C8 for comparison to those verified by logistic regression). We adjusted associations using confounders (Table 2; Figures B1-B9). Significant associations persisted after adjustment, in which the effect sizes only slightly changed. However, since the effect sizes were small, the selected latent candidate predictors might be weak predictors for the prediction models.

**Table 2.** Association between each latent candidate predictor and fetal growth restriction (FGR)/small for gestational age (SGA) by inverse probability weighting.

| Variable of interest | Unadjusted OR (95% CI; *p* value) | Adjusted OR (95% CI; *p* value) | Adjustment |
| --- | --- | --- | --- |
| Pregnancy-induced hypertension | 1.012 (1.011 to 1.013; *p*<0.001***) | 1.007 (1.007 to 1.008; *p*<0.001***) | Multiple pregnancies + Risk of adverse pregnancy by maternal age |
| Multiple pregnancies | 1.051 (1.047 to 1.054; *p*<0.001***) | 1.048 (1.044 to 1.052; *p*<0.001***) | Risk of adverse pregnancy by maternal age |
| Malaria | 0.993 (0.993 to 0.993; *p*<0.001***) | 0.993 (0.993 to 0.993; *p*<0.001***) | Low socioeconomic status |
| Varicella | 0.993 (0.993 to 0.993; *p*<0.001***) | 0.993 (0.993 to 0.993; *p*<0.001***) | (no adjustment) |
| Risk of adverse pregnancy by maternal age | 0.996 (0.996 to 0.996; *p*<0.001***) | 0.996 (0.996 to 0.996; *p*<0.001***) | (no adjustment) |
| Urinary tract infection | 1.068 (1.064 to 1.073; *p*<0.001***) | 1.137 (1.128 to 1.146; *p*<0.001***) | Risk of adverse pregnancy by maternal age |
| Placenta previa | 1.028 (1.026 to 1.031; *p*<0.001***) | 1.022 (1.02 to 1.024; *p*<0.001***) | Risk of adverse pregnancy by maternal age |
| Low socioeconomic status | 0.999 (0.999 to 1; *p*=0.05*) | 0.999 (0.999 to 1; *p*=0.05*) | (no adjustment) |

\* *p*≤.05; \*\* *p*≤.01; \*\*\* *P* ≤.001; CI, confidence interval; OR, odds ratio.

### The best prediction model

Only three of the five models were approximately well-calibrated (Figure 2a): the PC-ENR, PC-GBM, and DI-VNN. Among these models, the PC-GBM was considerably the best-calibrated (intercept −0.00098, 95% CI −0.13098 to 0.12902; slope 0.95, 95% CI 0.46 to 1.44; Brier score 0.0063). Nevertheless, the downstream analyses evaluated all of the well-calibrated models.

**Figure 2.**
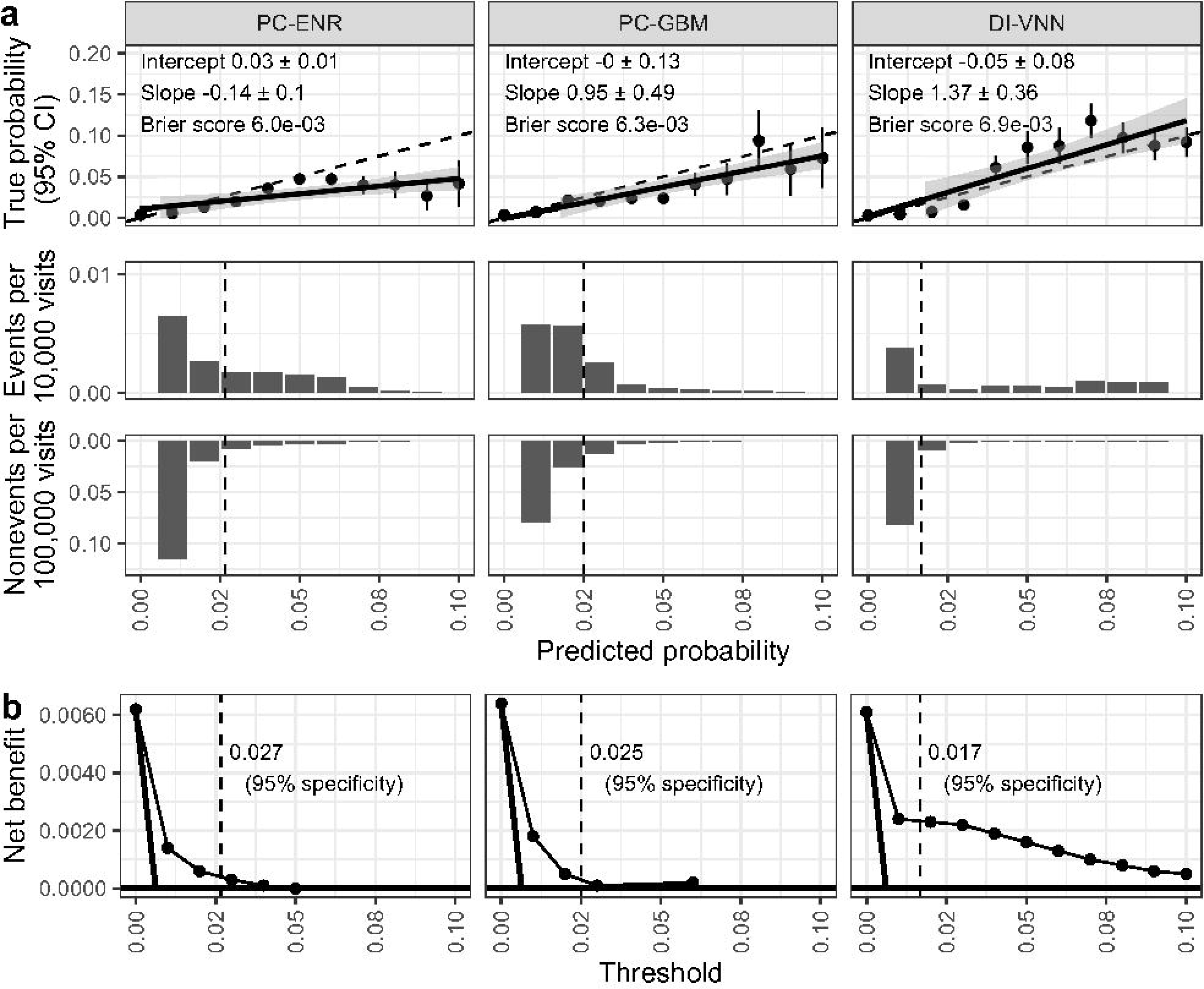
Model calibration (a) and clinical utility (b). We evaluated both using a calibration split (i.e., ∼20% of internal validation set) within the optimal range of predicted probabilities (equivalent to thresholds) across all of the models. This figure shows only the well-calibrated models. Solid lines with gray shading show the regression line and standard errors over point estimates of true probabilities. Dotted lines show a threshold of 95% specificity. CI, confidence interval; DI-VNN, deep-insight visible neural network; ENR, elastic net regression; GBM, gradient boosting machine; PC, principal component.

The net benefits of these models were higher than those of either the treat-all or treat-none prediction (Figure 2b). It also applied to those using a threshold of 95% specificity. With this threshold, we found the DI-VNN to be the best model in terms of clinical utility with a net benefit of 0.0023 (95% CI 0.0022 to 0.0024).

Regarding model explainability, both clinicians chose the DI-VNN among the well-calibrated models. They considered the plausibility of the top-five predictors according to the counterfactual probabilities (Table 3). One of the top predictors in the DI-VNN, i.e., severe preeclampsia, could change most of the predicted events into nonevents (PN 98.57%, 95% CI 98.5% to 98.63%) if the predictors were changed from positive to negative. Most of the nonevents were also changed into events (PS 2.08%, 95% CI 2.07% to 2.09%) if the predictors were changed from negative to positive. In addition, we also show the models’ parameters (Tables C9-C14) and all counterfactual probabilities (Tables C15-C17).

**Table 3.**
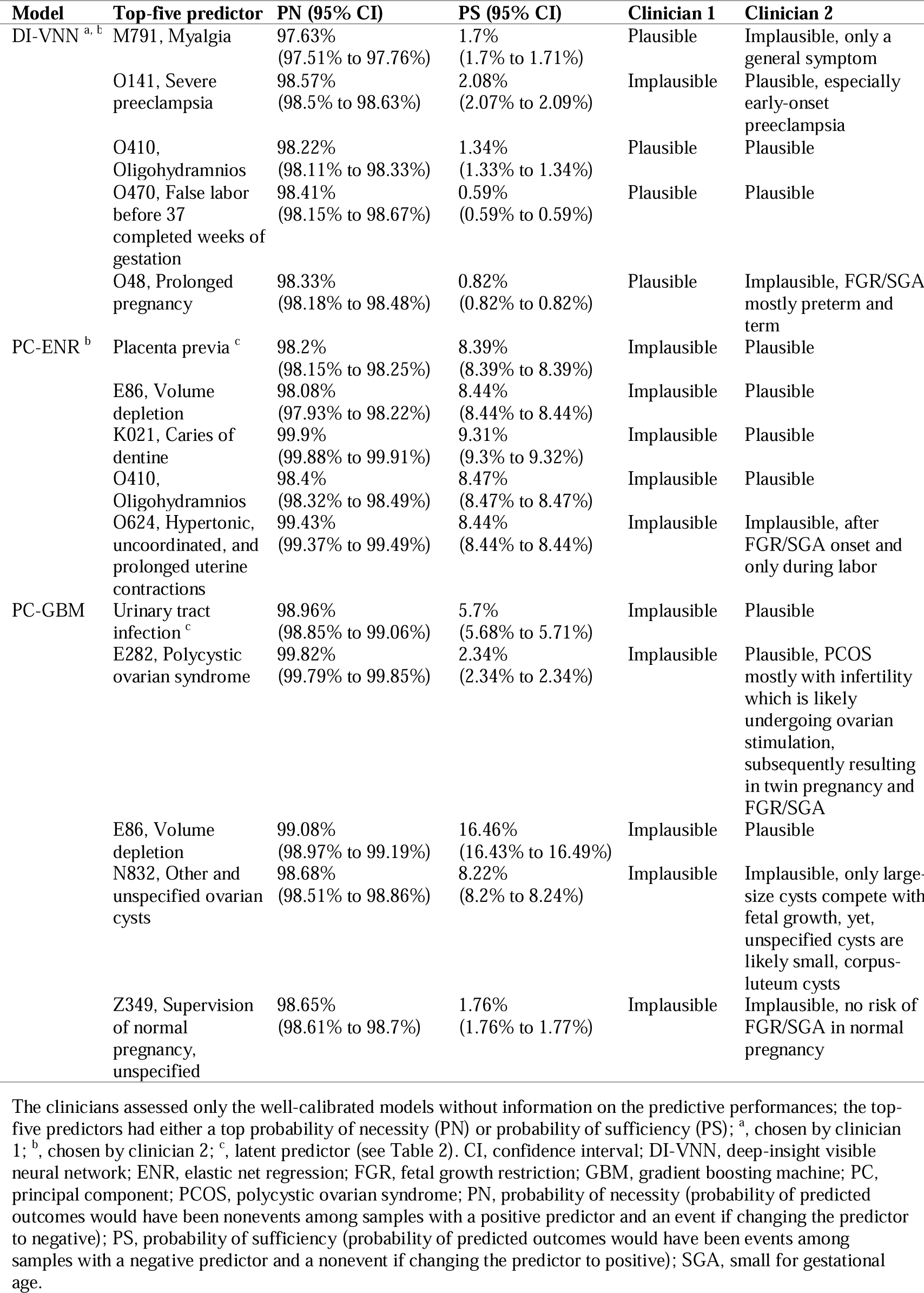
Model explainability by clinical assessments based on counterfactual probabilities.

The discrimination ability differed among the well-calibrated models according to the ROC curves (Figure 3) and AUROCs (Figure 4). Based on the internal calibration split, we identified that the best model was also the DI-VNN (AUROC 0.742, 95% CI 0.734 to 0.750; sensitivity 49.09%, 95% CI 47.60% to 50.58%). Using external validation, the AUROC of the DI-VNN (0.561, 95% CI 0.558 to 0.564) was considerably robust (i.e., the 95% CI >0.5).

**Figure 3.**
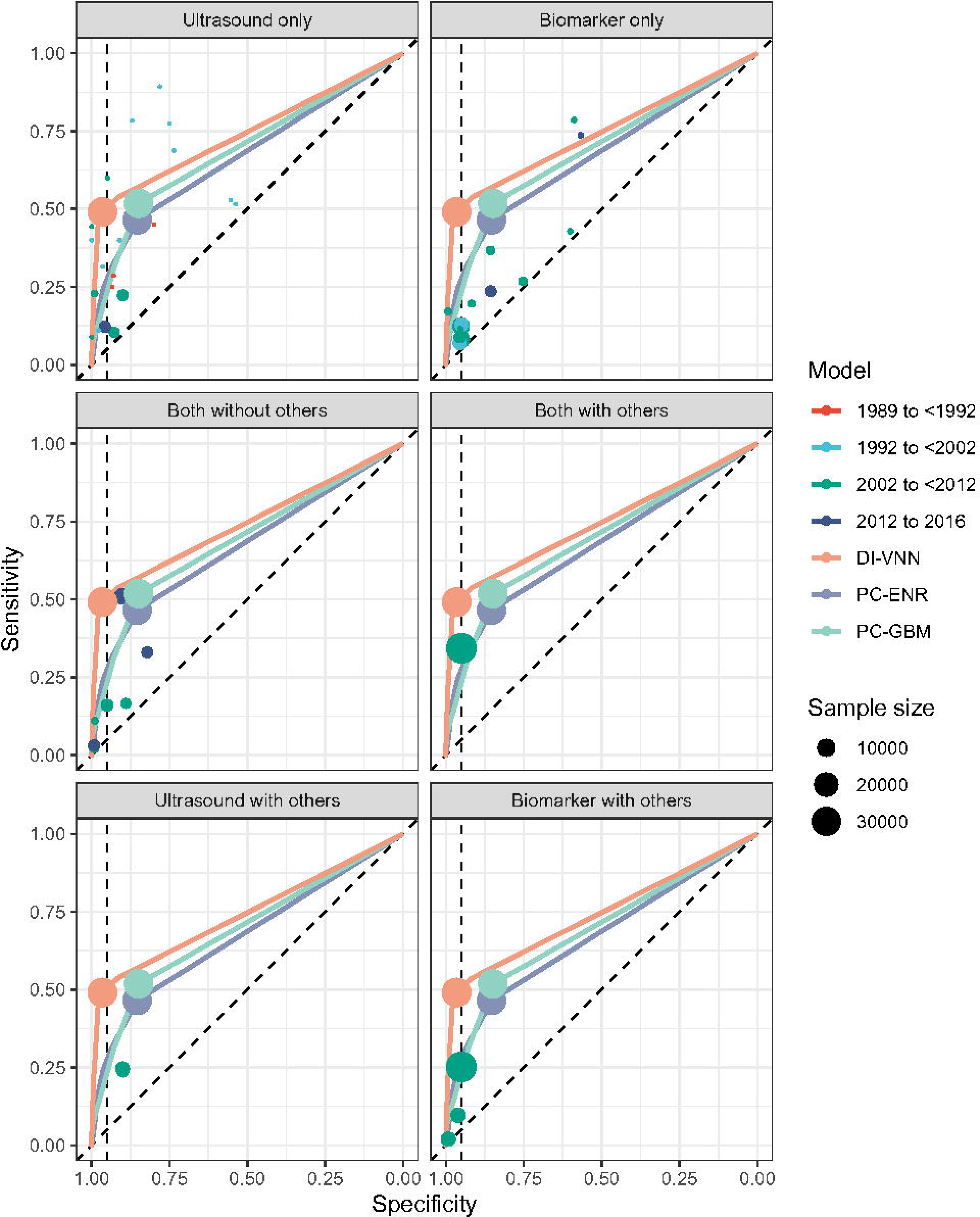
Model discrimination by receiver operating characteristics (ROC) curves. The evaluation used a calibration split (i.e., ∼20% of the internal validation set) for only the well-calibrated models. The vertical dotted lines show 95% specificity, while the diagonal dotted lines show the area under the ROC curve (AUROC) of 0.5 as a reference. DI-VNN, deep-insight visible neural network; ENR, elastic net regression; GBM, gradient boosting machine; PC, principal components.

**Figure 4.**
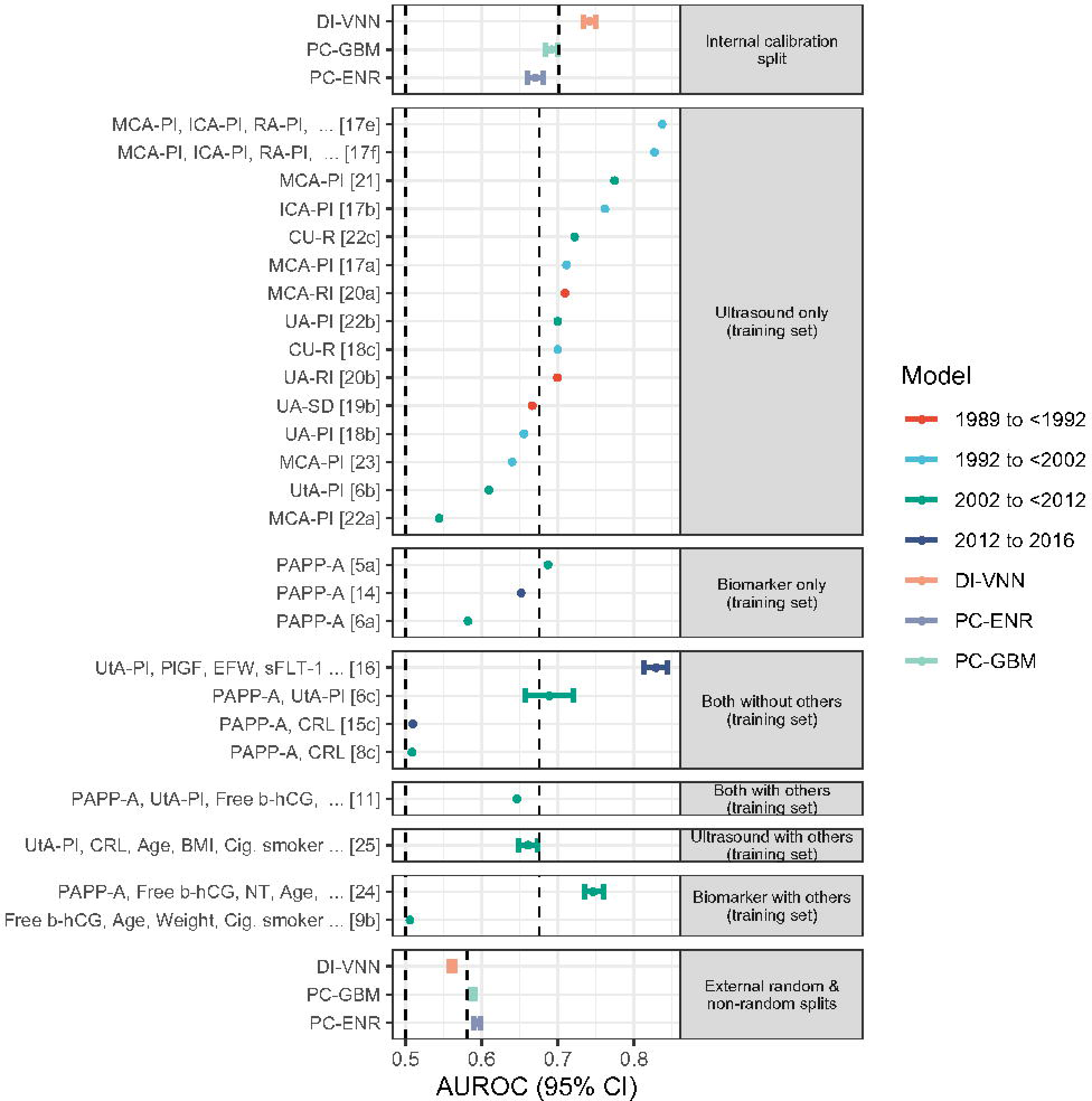
Model discrimination by the area under the receiver operating characteristics curves (AUROCs). This figure shows only the well-calibrated models. The vertical dotted lines show AUROCs of 0.5 and the averages using internal calibration split, training set, and external random and non-random splits. See Appendix D for details of eligible models from previous studies. If any predictor list of these models is too long, then it is truncated by “…”. βhCG, β-subunit human choriogonadotropin; BMI, body-mass index; Cig., cigarette; CRL, crown-rump length (fetus); CU-R, cerebral-umbilical ratio; DI-VNN, deep-insight visible neural network; EFW, estimated fetal weight; ENR, elastic net regression; GBM, gradient boosting machine; ICA, internal carotid artery; MCA, middle cerebral artery; NT, nuchal translucency thickness (fetus); PAPP-A, pregnancy-associated plasma protein-A; PC, principal component; PlGF, placental growth factor; PI, pulsatility index; RA, renal artery; RI, resistance index; ROC, receiver operating characteristics; SD, systolic-diastolic ratio; sFLT-1, soluble fms-like tyrosinase-1; UA, umbilical artery; UtA, uterine artery.

Furthermore, we compared the best model with those from previous studies. Only three studies fulfilled the eligibility criteria from three literature databases within the last 5 years. All of the studies were systematic reviews. Thus, we also searched eligible articles in the systematic reviews, including those published more than 5 years earlier. This step resulted in 381 records, including the three systematic reviews (Figure B10). We included 27 studies (Tables D1, D2) of these records for the meta-analysis. These studies used only a training set; thus, the evaluation metrics were extracted only from the training set. We categorized these studies based on the publication year such that the trend of the predictor modalities could be differentiated. By estimation, the DI-VNN was outperformed by those using ultrasound only from previous studies published from 1992 to <2002 (Figures 3, 4, Table C18). This finding was according to the sensitivity. However, those models were developed using smaller sample sizes (Figure 3) and were only evaluated using training sets (Figure 4). We also identified the latter issue for the previous model, which used both ultrasound and biomarkers but without other predictors, from a previous study published in a later year. Meanwhile, based on the AUROC using external validation splits, the DI-VNN was also estimated to outperform the previous models, which used either ultrasound or biomarkers without or with other predictors, from previous studies published from 2002 to 2016 (i.e., the two latest groups of publication years).

Eventually, we chose the DI-VNN to predict FGR/SGA in advance among 12- to-15-year-old females that visited primary, secondary, or tertiary care. Similar to the development pipeline of the prediction model, only a pregnant woman was eligible for the use of the DI-VNN to compute a predicted probability of FGR/SGA. We deployed the DI-VNN as a web application (https://predme.app/fgr_sga/). It can be used for future use or independent validation of the DI-VNN because it is open access.

## Discussion

We developed, validated, and deployed a web application to predict FGR/SGA in advance using the medical history of diagnoses and procedures. The prediction model for the web application was the DI-VNN, chosen among five prediction models in this study, using only an internal validation set. However, external validation also demonstrated the robustness of the DI-VNN’s predictive performance. It was also comparable to those developed in the previous studies, which used ultrasound and biomarkers without or with other predictors.

For predicting FGR/SGA, the previous models, as systematically reviewed in this study (Table D2), mainly required either ultrasound or biomarker measurements and a specific range of gestational ages. The models included those which were competitive with the DI-VNN based on the AUROC by internal validation (Figure 4). The models were by Shlossman, et al ^27^ (nos. 17e, 17f, and 17b), Bednarek, et al ^28^ (no. 21), Valiño, et al ^29^ (no. 16), and Poon, et al ^30^ (no. 24). Conversely, external validation estimated that the DI-VNN would outperform the other previous models with the similar requirements. The models were by Bano, et al ^31^ (no. 22a), Carbone, et al ^32^ (no. 15c), Leung, et al ^33^ (no. 8c), and Krantz, et al ^34^ (no. 9b). Furthermore, evaluation of the previous models used training sets only, in which the predictive performances might have been overoptimistic.

The DI-VNN required neither ultrasound nor biomarkers without or with other predictors. We would expect wider access for FGR/SGA predictions as either (1) a prediction model for use in resource-limited settings or (2) a preliminary prediction model before ordering advanced predictor measurements. However, the DI-VNN needs an impact study to evaluate its effect on patient outcomes in various settings.

An effective prevention for FGR was given by ≤16 weeks’ gestation.^3^ To widen prevention time window, more clinical trials are needed. These studies are more efficient if they are conducted among pregnant women with higher risk, as predicted by the DI-VNN. Since it did not require a specific range of gestational ages, the DI-VNN opens more opportunities to conduct such trials.

One of the strengths of this study were no requirements from our models, including the DI-VNN, for either ultrasound or biomarker measurements to predict FGR/SGA in advance. We could apply our models to a general population of pregnant women. Furthermore, our model did not require a specific gestational age range for computing the predicted probability. Unlike previous studies, we also conducted external validation to estimate the future predictive performance of the DI-VNN.

However, we also identified several limitations of this study. The predictive performance of the best model, i.e., the DI-VNN, was considerably moderate according to the AUROC as was the sensitivity at 95% specificity using an internal validation set. However, previous models also achieved similar predictive performances. Another limitation was that medical histories from electronic health records might take time to execute; yet, this is considerably more achievable in many settings. It still needs to be determined if the DI-VNN can improve patient outcomes. Nevertheless, this problem is not exclusive to this study because many previous studies in medicine have yet to evaluate the impacts of their prediction models.^35^

## Data Availability

The social security administrator provided the data for health or badan penyelenggara jaminan sosial (BPJS) kesehatan in Indonesia, with restrictions (access approval no.: 510/PPID/1223). Data are available from the authors upon reasonable request and with permission of the BPJS Kesehatan. The latter needs a request to the BPJS Kesehatan for their sample dataset published in August 2019.

https://e-ppid.bpjs-kesehatan.go.id/

https://github.com/herdiantrisufriyana/fgr_sga

## Abbreviations

AUROC: area under the receiver operating characteristics curve
CI: confidence interval
DI-VNN: deep-insight visible neural network
ENR: elastic net regression
FGR: fetal growth restriction
GBM: gradient boosting machine
ICD-10: International Classification of Disease version 10
OR: odds ratio
PC: principal component
PN: probability of necessity
PS: probability of sufficiency
RF: random forest
ROC: receiver operating characteristics
RR: ridge regression
SGA: small for gestational age

## Appendices

## Appendix A

Transparent reporting of a multivariable prediction model for individual prognosis or diagnosis (TRIPOD) checklist.

## Appendix B

Table B1. Guidelines for developing and reporting machine learning predictive models in biomedical research.

Table B2. Prediction model risk of bias assessment tools (PROBAST).

Table B3. Clinical checklists for assessing the suitability of machine learning applications in healthcare.

Table B4. Preferred reporting items for systematic reviews and meta-analyses (PRISMA) 2020 expanded checklist.

Figure B1. Association diagram of pregnancy-induced hypertension and fetal growth restriction (FGR)/small for gestational age (SGA).

Figure B2. Association diagram of multiple pregnancies and fetal growth restriction (FGR)/small for gestational age (SGA).

Figure B3. Association diagram of malaria and fetal growth restriction (FGR)/small for gestational age (SGA).

Figure B4. Association diagram of varicella and fetal growth restriction (FGR)/small for gestational age (SGA).

Figure B5. Association diagram of risk of an adverse pregnancy by maternal age and fetal growth restriction (FGR)/small for gestational age (SGA).

Figure B6. Association diagram of urinary tract infections and fetal growth restriction (FGR)/small for gestational age (SGA).

Figure B7. Association diagram of placenta previa and fetal growth restriction (FGR)/small for gestational age (SGA).

Figure B8. Association diagram of a low socioeconomic status and fetal growth restriction (FGR)/small for gestational age (SGA).

Figure B9. Unified association diagram and fetal growth restriction (FGR)/small for gestational age (SGA).

Figure B10. Flow diagram to find comparable models from previous studies.

## Appendix C

Table C1. R package versions.

Table C2. Codes for determining delivery or immediate after-delivery care.

Table C3. Candidate predictors with non-zero variances.

Table C4. Excluded codes in the training set that may leak outcome information.

Table C5. Pair-wise Pearson correlations to identify redundant candidate predictors.

Table C6. Results of systematic human learning and data availability for latent candidate predictors.

Table C7. International classification of disease (ICD)-10 codes or demographical variables for latent candidate predictors.

Table C8. Association tests by logistic regression (LR) and inverse probability weighting (IPW).

Table C9. Principal component (PC) weights.

Table C10. Principal-component elastic net regression (PC-ENR) weights.

Table C11. Principal-component gradient boosting machine (PC-GBM) variable importance.

Table C12. Selected predictors based on a differential analysis with a false discovery rate (FDR) of <0.05.

Table C13. Deep-insight visible neural network (DI-VNN) intermediate outputs.

Table C14. Connections between ontologies under the root of a deep-insight visible neural network (DI-VNN).

Table C15. Probabilities of necessity (PN) and sufficiency (PSs) of predictors in a principal-component elastic net regression (PC-ENR).

Table C16. Probabilities of necessity (PN) and sufficiency (PS) of predictors in a principal-component gradient boosting machine (PC-GBM).

Table C17. Probabilities of necessity (PN) and sufficiency (PS) of predictors in a deep-insight visible neural network (DI-VNN).

Table C18. Extracted data of available evaluation metrics for comparable models.

## Appendix D

Table D1. Search and filter results of previous studies.

Table D2. Comparable models to evaluate the success criteria.

## Author contributions

**HS:** Conceptualization, Methodology, Software, Validation, Formal Analysis, Investigation, Data Curation, Writing—Original Draft, Visualization, Project Administration, Funding Acquisition. **FZA:** Validation, Formal Analysis, Data Curation, Writing—Review & Editing. **AZZAH:** Validation, Formal Analysis, Data Curation, Writing—Review & Editing. **YWW:** Conceptualization, Methodology, Writing— Review & Editing, Supervision. **ECYS:** Conceptualization, Methodology, Resources, Writing—Review & Editing, Supervision, Funding acquisition. All authors have read and approved the manuscript and agreed to be accountable for all aspects of the work in ensuring that questions related to the accuracy or integrity of any part of the work are appropriately investigated and resolved.

## Acknowledgments

The BPJS Kesehatan in Indonesia permitted access to the sample dataset in this study. This study was funded by: (1) the Postdoctoral Accompanies Research Project from the National Science and Technology Council (NSTC) of Taiwan (grant no.: NSTC111-2811-E-038-003-MY2), and the Lembaga Penelitian dan Pengabdian kepada Masyarakat (LPPM) Universitas Nahdlatul Ulama Surabaya of Indonesia (grant no.: 161.4/UNUSA/Adm-LPPM/III/2021) to Herdiantri Sufriyana; and (2) the Ministry of Science and Technology (MOST) of Taiwan (grant nos.: MOST110-2628-E-038-001 and MOST111-2628-E-038-001-MY2), and the Higher Education Sprout Project from the Ministry of Education (MOE) of Taiwan (grant no.: DP2-111-21121-01-A-05) to Emily Chia-Yu Su. These funding bodies had no role in the study design; in the collection, analysis, and interpretation of data; in the writing of the report; or in the decision to submit the article for publication.

## Data Statements

The social security administrator provided the data for health or *badan penyelenggara jaminan sosial (BPJS) kesehatan* in Indonesia, with restrictions (access approval no.: 510/PPID/1223). Data are available from the authors upon reasonable request and with permission of the BPJS Kesehatan. The latter needs a request to the BPJS Kesehatan for their sample dataset published in August 2019 via https://e-ppid.bpjs-kesehatan.go.id/. The analytical codes are available at https://github.com/herdiantrisufriyana/fgr_sga.

## References

[1] Fetal growth restriction: Acog practice bulletin, number 227. Obstet Gynecol 2021;137:e16–e28. doi: 10.1097/aog.0000000000004251.

[2] Nardozza LM, Caetano AC, Zamarian AC, et al. Fetal growth restriction: Current knowledge. Arch Gynecol Obstet 2017;295:1061–77. doi: 10.1007/s00404-017-4341-9.

[3] Roberge S, Nicolaides K, Demers S, Hyett J, Chaillet N, Bujold E. The role of aspirin dose on the prevention of preeclampsia and fetal growth restriction: Systematic review and meta-analysis. Am J Obstet Gynecol 2017;216:110–20.e6. doi: 10.1016/j.ajog.2016.09.076.

[4] Pedroso MA, Palmer KR, Hodges RJ, Costa FDS, Rolnik DL. Uterine artery doppler in screening for preeclampsia and fetal growth restriction. Rev Bras Ginecol Obstet 2018;40:287–93. doi: 10.1055/s-0038-1660777.

[5] Mallia T, Grech A, Hili A, Calleja-Agius J, Pace NP. Genetic determinants of low birth weight. Minerva Ginecol 2017;69:631–43. doi: 10.23736/s0026-4784.17.04050-3.

[6] Lawn JE, Cousens S, Zupan J. 4 million neonatal deaths: When? Where? Why? Lancet 2005;365:891–900. doi: 10.1016/s0140-6736(05)71048-5.

[7] Ausbeck EB, Allman PH, Szychowski JM, Subramaniam A, Katheria A. Neonatal outcomes at extreme prematurity by gestational age versus birth weight in a contemporary cohort. Am J Perinatol 2021;38:880–88. doi: 10.1055/s-0040-1722606.

[8] Tabet M, Flick LH, Xian H, Jen Jen C. Smallness at birth and neonatal death: Reexamining the current indicator using sibling data. Am J Perinatol 2021;38:76–81. doi: 10.1055/s-0039-1694761.

[9] Lehtonen L, Gimeno A, Parra-Llorca A, Vento M. Early neonatal death: A challenge worldwide. Semin Fetal Neonatal Med 2017;22:153–60. doi: 10.1016/j.siny.2017.02.006.

[10] Colella M, Frérot A, Novais ARB, Baud O. Neonatal and long-term consequences of fetal growth restriction. Curr Pediatr Rev 2018;14:212–18. doi: 10.2174/1573396314666180712114531.

[11] Umran RM, Al-Jammali A. Neonatal outcomes in a level ii regional neonatal intensive care unit. Pediatr Int 2017;59:557–63. doi: 10.1111/ped.13200.

[12] Horbar JD, Edwards EM, Greenberg LT, et al. Racial segregation and inequality in the neonatal intensive care unit for very low-birth-weight and very preterm infants. JAMA Pediatr 2019;173:455–61. doi: 10.1001/jamapediatrics.2019.0241.

[13] Ho T, Zupancic JAF, Pursley DM, Dukhovny D. Improving value in neonatal intensive care. Clin Perinatol 2017;44:617–25. doi: 10.1016/j.clp.2017.05.009.

[14] Bettiol A, Avagliano L, Lombardi N, et al. Pharmacological interventions for the prevention of fetal growth restriction: A systematic review and network meta-analysis. Clin Pharmacol Ther 2021;110:189–99. doi: 10.1002/cpt.2164.

[15] Audette MC, Kingdom JC. Screening for fetal growth restriction and placental insufficiency. Semin Fetal Neonatal Med 2018;23:119–25. doi: 10.1016/j.siny.2017.11.004.

[16] Luntsi G, Ugwu AC, Nkubli FB, Emmanuel R, Ochie K, Nwobi CI. Achieving universal access to obstetric ultrasound in resource constrained settings: A narrative review. Radiography (Lond) 2021;27:709–15. doi: 10.1016/j.radi.2020.10.010.

[17] Selvaratnam RJ, Wallace EM, Flenady V, Davey MA. Risk factor assessment for fetal growth restriction, are we providing best care? Aust N Z J Obstet Gynaecol 2020;60:470–73. doi: 10.1111/ajo.13147.

[18] Wagstaff A, Neelsen S. A comprehensive assessment of universal health coverage in 111 countries: A retrospective observational study. Lancet Glob Health 2020;8:e39–e49. doi: 10.1016/s2214-109x(19)30463-2.

[19] Andreasen LA, Tabor A, Nørgaard LN, et al. Why we succeed and fail in detecting fetal growth restriction: A population-based study. Acta Obstet Gynecol Scand 2021;100:893–99. doi: 10.1111/aogs.14048.

[20] Sufriyana H, Husnayain A, Chen YL, et al. Comparison of multivariable logistic regression and other machine learning algorithms for prognostic prediction studies in pregnancy care: Systematic review and meta-analysis. JMIR Med Inform 2020;8:e16503. doi: 10.2196/16503.

[21] Sufriyana H, Wu YW, Su EC. Artificial intelligence-assisted prediction of preeclampsia: Development and external validation of a nationwide health insurance dataset of the bpjs kesehatan in indonesia. EBioMedicine 2020;54:102710. doi: 10.1016/j.ebiom.2020.102710.

[22] Moons KG, Altman DG, Reitsma JB, et al. Transparent reporting of a multivariable prediction model for individual prognosis or diagnosis (tripod): Explanation and elaboration. Ann Intern Med 2015;162:W1–73. doi: 10.7326/m14-0698.

[23] Sufriyana H, Wu YW, Su EC-Y. Human and machine learning pipelines for responsible clinical prediction using high-dimensional data. Protocol Exchange 2021. doi: 10.21203/rs.3.pex-1655/v1.

[24] Ariawan I, Sartono B, Jaya C. Sample dataset of the bpjs kesehatan 2015-2016. Jakarta BPJS Kesehatan; 2019.

[25] Moraffah R, Karami M, Guo R, Raglin A, Liu H. Causal interpretability for machine learning-problems, methods and evaluation. ACM SIGKDD Explorations Newsletter 2020;22:18–33. doi, PMID.

[26] Page MJ, McKenzie JE, Bossuyt PM, et al. The prisma 2020 statement: An updated guideline for reporting systematic reviews. Bmj 2021;372:n71. doi: 10.1136/bmj.n71.

[27] Shlossman P, Scisione A, Manley J, Colmorgen G, Weiner S. Doppler assessment of the intrafetal vasculature in the identification of intrauterine growth retardation. Which vessel is ‘best’ or is a combination better? American Journal of Obstetrics & Gynecology 1998;178:S88. doi: 10.1016/j.ajog.2012.01.022.

[28] Bednarek M, Dubiel M, Brêborowicz GH. P05.18: Doppler velocimetry in m1 and m2 segments of middle cerebral artery in pregnancies complicated by intrauterine growth restriction. Ultrasound in Obstetrics & Gynecology 2004;24:300–01. doi: 10.1002/uog.1423.

[29] Valiño N, Giunta G, Gallo DM, Akolekar R, Nicolaides KH. Biophysical and biochemical markers at 30-34 weeks’ gestation in the prediction of adverse perinatal outcome. Ultrasound Obstet Gynecol 2016;47:194–202. doi: 10.1002/uog.14928.

[30] Poon LC, Karagiannis G, Staboulidou I, Shafiei A, Nicolaides KH. Reference range of birth weight with gestation and first-trimester prediction of small-for-gestation neonates. Prenat Diagn 2011;31:58–65. doi: 10.1002/pd.2520.

[31] Bano S, Chaudhary V, Pande S, Mehta V, Sharma A. Color doppler evaluation of cerebral-umbilical pulsatility ratio and its usefulness in the diagnosis of intrauterine growth retardation and prediction of adverse perinatal outcome. Indian J Radiol Imaging 2010;20:20–5. doi: 10.4103/0971-3026.59747.

[32] Carbone JF, Tuuli MG, Bradshaw R, Liebsch J, Odibo AO. Efficiency of first-trimester growth restriction and low pregnancy-associated plasma protein-a in predicting small for gestational age at delivery. Prenat Diagn 2012;32:724–9. doi: 10.1002/pd.3891.

[33] Leung TY, Sahota DS, Chan LW, et al. Prediction of birth weight by fetal crown-rump length and maternal serum levels of pregnancy-associated plasma protein-a in the first trimester. Ultrasound Obstet Gynecol 2008;31:10–4. doi: 10.1002/uog.5206.

[34] Krantz D, Goetzl L, Simpson JL, et al. Association of extreme first-trimester free human chorionic gonadotropin-beta, pregnancy-associated plasma protein a, and nuchal translucency with intrauterine growth restriction and other adverse pregnancy outcomes. Am J Obstet Gynecol 2004;191:1452–8. doi: 10.1016/j.ajog.2004.05.068.

[35] Zhou Q, Chen ZH, Cao YH, Peng S. Clinical impact and quality of randomized controlled trials involving interventions evaluating artificial intelligence prediction tools: A systematic review. NPJ Digit Med 2021;4:154. doi: 10.1038/s41746-021-00524-2.

