## Appendix B for "Widely accessible prognostication using medical history for fetal growth restriction and small for gestational age in nationwide insured women"

Table B1. Guidelines for developing and reporting machine learning predictive models in biomedical research.

| Item no. | Section | Topic | Checklist item with the description and/or the corresponding text | | |
| --- | --- | --- | --- | --- | --- |
| 1 | Title | Nature of study | ✓ |  | Identify the report as introducing a predictive model |
|  |  |  |  | • | Widely accessible prognostication using medical history for fetal growth restriction and small for gestational age in nationwide insured women |
| 2 | Abstract | Structured summary | ✓ |  | Background |
|  |  |  |  | • | Prevention of fetal growth restriction (FGR)/small for gestational age (SGA) is adequate if screening is accurate. Ultrasound and biomarkers can achieve this goal; however, both are often inaccessible. |
|  |  |  | ✓ |  | Objectives |
|  |  |  |  | • | This study aimed to develop, validate, and deploy a prognostic prediction model for screening FGR/SGA using only medical history. |
|  |  |  | ✓ |  | Data sources |
|  |  |  |  | • | From a nationwide health insurance database (n=1,697,452), we retrospectively selected visits of 12-to-55-year-old females to 22,024 healthcare providers of primary, secondary, and tertiary care. |
|  |  |  | ✓ |  | Performance metrics of the predictive model or models, in both point estimates and confidence intervals |
|  |  |  |  | • | The best prediction model was a deep-insight visible neural network (DI-VNN). It had an AUROC of 0.742 (95% CI 0.734 to 0.750) and a sensitivity of 49.09% (95% CI 47.60% to 50.58% using a threshold with 95% specificity). |
|  |  |  | ✓ |  | Conclusion including the practical value of the developed predictive model or models |
|  |  |  |  | • | Our model used only medical history to improve accessibility for FGR/SGA screening. |
| 3 | Introduction | Rationale | ✓ |  | Identify the clinical goal |
|  |  |  |  | • | The prevention method depends on FGR/SGA predictions with a clinically acceptable predictive performance. However, most settings lack accessibility to predictors in existing prediction models. |
|  |  |  | ✓ |  | Review the current practice and prediction accuracy of any existing models |
|  |  |  |  | • | Since a low-cost method such as symphysis fundal height was not recommended by a Cochrane review, mainly due to low sensitivity (~17%), there is a trend to employ either ultrasound or biomarker measurements for FGR/SGA screening. |
|  |  |  |  | • | Screening by medical history is also independent of the number of pregnancy consultations on which FGR detection depends (hazard ratio 1.15, 95% confidence interval [CI] 1.05 to 1.26). However, studies have yet to develop a screening method for FGR/SGA using only medical history. |
| 4 | Introduction | Objectives | ✓ |  | State the nature of study being predictive modelling, defining the target of prediction |
|  |  |  |  | • | We aimed to develop, validate, and deploy a prognostic prediction model for screening FGR/SGA using only medical history in nationwide insured women. |
|  |  |  | ✓ |  | Identify how the prediction problem may benefit the clinical goal |
|  |  |  |  | • | Prognostic predictions of FGR/SGA using medical histories can be either a prediction model for use in resource-limited settings or a preliminary prediction model before ordering ultrasound and biomarker measurements. |
| 5 | Methods | Describe the setting | ✓ |  | Identify the clinical setting for the target predictive model |
|  |  |  |  | ❖ | Clinical setting: primary, secondary, and tertiary care |
|  |  |  |  | • | It included all affiliated healthcare providers (n=22,024) at all levels (i.e., primary, secondary, and tertiary care). |
|  |  |  | ✓ |  | Identify the modelling context in terms of facility type, size, volume, and duration of available data |
|  |  |  |  | ❖ | Facility type: healthcare providers affiliated to a nationwide health insurance |
|  |  |  |  | ❖ | Size: 22,024 |
|  |  |  |  | ❖ | Volume: 1,697,452 |
|  |  |  |  | ❖ | Duration: 2 years |
|  |  |  |  | • | The dataset was a cross-sectional, random sampling of ~1% of insurance holders within 2 years up to 2016. This sampling applied stratification by a family category and healthcare provider. It included all affiliated healthcare providers (n=22,024) at all levels (i.e., primary, secondary, and tertiary care). |
|  |  |  |  | • | From the database (n=1,697,452), we selected 12-to-55-year-old females (n=169,746) that had visited (n=507,319) primary, secondary, or tertiary care |
| 6 | Methods | Define the prediction problem | ✓ |  | Define a measurement for the prediction goal (per patient or per hospitalization or per type of outcome) |
|  |  |  |  | ❖ | Prediction goal: per visit |
|  |  |  |  | • | This study developed a prediction model for detecting in advance a visit by a subject who would be diagnosed with either FGR or SGA. |
|  |  |  | ✓ |  | Determine that the study is retrospective or prospective |
|  |  |  |  | ❖ | Study design: |
|  |  |  |  | • | We applied a retrospective design to select subjects from a public dataset version 2 (August 2019; access approval no.: 510/PPID/1223) of a nationwide health insurance database in Indonesia. |
|  |  |  | ✓ |  | Identify the problem to be prognostic or diagnostic |
|  |  |  |  | ❖ | Prediction problem: prognostic |
|  |  |  |  | • | This study developed a prediction model for detecting in advance a visit by a subject who would be diagnosed with either FGR or SGA. |
|  |  |  | ✓ |  | Determine the form of the prediction model: (1) classification if the target variable is categorical, (2) regression if the target variable is continuous, (3) survival prediction if the target variable is the time to an event. |
|  |  |  |  | ❖ | Prediction model form: classification |
|  |  |  |  | • | This study developed a prediction model for detecting in advance a visit by a subject who would be diagnosed with either FGR or SGA. |
|  |  |  | – |  | Translate survival prediction into a regression problem, with the target measured over a temporal window following the time of prediction. |
|  |  |  |  | ❖ | Not applicable because we did not use survival rate for target variable. |
|  |  |  | ✓ |  | Explain practical costs of prediction errors (e.g., implications of under-diagnosis or over-diagnosis) |
|  |  |  |  | ❖ | Under-prognosis: neonatal morbidity and mortality, higher healthcare costs |
|  |  |  |  | ❖ | Over-prognosis: higher healthcare costs |
|  |  |  |  | • | Under-prognosis could cause a subject to lose the chance for FGR/SGA prevention, which may lead to neonatal morbidity and mortality with higher healthcare costs. Meanwhile, over-prognosis may lead to higher healthcare costs by ordering more tests to confirm the risk of FGR/SGA. |
|  |  |  | ✓ |  | Defining quality metrics for prediction models |
|  |  |  |  | ❖ | Quality metrics: sensitivity at 95% specificity |
|  |  |  |  | • | Thus, for the best tradeoff, we considered resolving under-prognosis. It could be indicated by an acceptable sensitivity at 95% specificity but using more-accessible predictors. |
|  |  |  | ✓ |  | Define the success criteria for prediction (e.g., based on metrics in internal validation or external validation in the context of the clinical problem) |
|  |  |  |  | ❖ | Success criteria: acceptable sensitivity at 95% specificity but using more-accessible predictors |
|  |  |  |  | • | It could be indicated by an acceptable sensitivity at 95% specificity but using more-accessible predictors. Nevertheless, we compared our prediction models with those from previous studies selected by systematic review methods to evaluate if our predictive modeling was successful. |
| 7 | Methods | Prepare data for model building | ✓ |  | Identify relevant data sources and quote the ethics approval number for data access |
|  |  |  |  | ❖ | Data source: a cross-sectional sample dataset of a nationwide health insurance |
|  |  |  |  | ❖ | Ethic waiver number: N202106025 |
|  |  |  |  | ❖ | Data access approval number: 510/PPID/1223 |
|  |  |  |  | • | The Taipei Medical University Joint Institutional Review Board exempted this project from the ethical review (TMU-JIRB no.: N202106025). |
|  |  |  |  | • | We applied a retrospective design to select subjects from a public dataset version 2 (August 2019; access approval no.: 510/PPID/1223) of a nationwide health insurance database in Indonesia. The dataset was a cross-sectional, random sampling of ~1% of insurance holders within 2 years up to 2016. |
|  |  |  |  | • | This sampling applied stratification by a family category and healthcare provider. It included all affiliated healthcare providers (n=22,024) at all levels (i.e., primary, secondary, and tertiary care). |
|  |  |  | ✓ |  | State the inclusion and exclusion criteria for data |
|  |  |  |  | ❖ | Inclusion criteria: females aged 12 to 55 years who had visited primary, secondary, or tertiary care facilities |
|  |  |  |  | ❖ | Exclusion criteria: all visits afterward if a woman was pregnant and had a delivery |
|  |  |  |  | • | The inclusion criteria were females aged 12 to 55 years who had visited primary, secondary, or tertiary care facilities. All visits afterward were excluded if a woman was pregnant and had a delivery. If a woman became pregnant twice within the dataset period, then different identifiers were assigned to differentiate the pregnancy periods of that woman. |
|  |  |  | ✓ |  | Describe the time span of data and the sample or cohort size |
|  |  |  |  | ❖ | Time span of data: visits up to the end of pregnancy |
|  |  |  |  | ❖ | Sample size: 507,319 |
|  |  |  |  | • | The inclusion criteria were females aged 12 to 55 years who had visited primary, secondary, or tertiary care facilities. All visits afterward were excluded if a woman was pregnant and had a delivery. If a woman became pregnant twice within the dataset period, then different identifiers were assigned to differentiate the pregnancy periods of that woman. |
|  |  |  |  | • | From the database (n=1,697,452), we selected 12-to-55-year-old females (n=169,746) that had visited (n=507,319) primary, secondary, or tertiary care |
|  |  |  | ✓ |  | Define the observational units on which the response variable and predictor variables are defined |
|  |  |  |  | ❖ | Response variable for classification task: O365 (mothers) or P05 (fetuses) |
|  |  |  |  | ❖ | Predictors: medical histories of diagnoses and procedures (mothers) |
|  |  |  |  | • | The event outcome definition in this study utilized the International Classification of Disease version 10 (ICD-10) codes. These were codes preceded by either O365 (maternal care for known or suspected fetal growth) or P05 (disorders of newborns related to slow fetal growth and fetal malnutrition). Both codes indicating FGR and SGA were assigned with those respectively for mothers and fetuses/newborns. A nonevent outcome was assigned if the end of pregnancy was identified within the dataset period by the codes for determining delivery. Otherwise, we assigned an outcome to a censored one. |
|  |  |  |  | • | Candidate predictors were only medical histories of diagnoses and procedures. These were either single or multiple ICD-10 codes. As extensively described in the protocol, the preprocessing of candidate predictors consisted of (1) preventing zero variance, perfect separation and leakage of the outcome, and redundant predictors; (2) simulating real-world data; and (3) systematically determining the multiple ICD-10 codes for defining latent candidate predictors based on prior knowledge. |
|  |  |  | ✓ |  | Define the predictor variables. Extra caution is needed to prevent information leakage from the response variable to predictor variables. |
|  |  |  |  | ❖ | Predictors: medical histories of diagnoses and procedures |
|  |  |  |  | • | Candidate predictors were only medical histories of diagnoses and procedures. These were either single or multiple ICD-10 codes. As extensively described in the protocol, the preprocessing of candidate predictors consisted of (1) preventing zero variance, perfect separation and leakage of the outcome, and redundant predictors; (2) simulating real-world data; and (3) systematically determining the multiple ICD-10 codes for defining latent candidate predictors based on prior knowledge. |
|  |  |  | ✓ |  | Describe the data pre-processing performed, including data cleaning and transformation. Remove outliers with impossible or extreme responses; state any criteria used for outlier removal. |
|  |  |  |  | ❖ | Data cleaning: preventing zero variance, perfect separation and leakage of the outcome, and redundant predictors |
|  |  |  |  | ❖ | Data transformation: historical rates (modified Kaplan-Meier estimators) |
|  |  |  |  | ❖ | Outlier removal: none |
|  |  |  |  | • | As extensively described in the protocol, the preprocessing of candidate predictors consisted of (1) preventing zero variance, perfect separation and leakage of the outcome, and redundant predictors; (2) simulating real-world data; and (3) systematically determining the multiple ICD-10 codes for defining latent candidate predictors based on prior knowledge. |
|  |  |  | ✓ |  | State how missing values were handled |
|  |  |  |  | ❖ | Missing value handling: historical rates (modified Kaplan-Meier estimators) |
|  |  |  |  | • | As extensively described in the protocol, the preprocessing of candidate predictors consisted of (1) preventing zero variance, perfect separation and leakage of the outcome, and redundant predictors; (2) simulating real-world data; and (3) systematically determining the multiple ICD-10 codes for defining latent candidate predictors based on prior knowledge. |
|  |  |  | ✓ |  | Describe the basic statistics of the dataset, particularly of the response variable. These include the ratio of positive to negative classes for a classification problem and the distribution of the response variable for regression problem. |
|  |  |  |  | ❖ | Classification outcome ratio: 117:26,459 = 1:226 = 0.4% |
|  |  |  |  | • | To characterize subjects in the internal validation set, we also included subjects with uncensored outcomes (n=26,576). |
|  |  |  | ✓ |  | Define the model validation strategies. Internal validation is the minimum requirement; external validation should also be performed whenever possible. |
|  |  |  |  | ❖ | Internal validation: five-fold cross-validation, bootstrapping |
|  |  |  |  | ❖ | External validation: random, geographical, and temporal splits |
|  |  |  |  | • | This study split the dataset into internal and external validation sets. The latter was by random and non-random sampling. We applied geographical and temporal splitting, i.e., respectively, by cities and days for stratification in non-random sampling, as extensively described in the protocol. These two splitting methods constituted ~20% of the dataset. Subsequently, we applied random sampling to split out ~20% of the remainder of the dataset. This sampling constructed another subset for external validation. For simplicity, external validation aggregately used all subsets by these three splitting methods. Only ~64% of the dataset was for internal validation. For recalibration, we used ~20% of the internal validation set. Before recalibration, we applied two resampling methods to evaluate the predictive performance. These were (1) five-fold cross-validation for hyperparameter tuning and (2) bootstrapping for the final training. |
|  |  |  | ✓ |  | Specify the internal validation strategy. Common methods include random split, time-based split, and patient-based split. |
|  |  |  |  | ❖ | Internal validation: five-fold cross-validation, bootstrapping |
|  |  |  |  | • | Before recalibration, we applied two resampling methods to evaluate the predictive performance. These were (1) five-fold cross-validation for hyperparameter tuning and (2) bootstrapping for the final training. |
|  |  |  | ✓ |  | Define the validation metrics. For regression problems, the normalized root-mean-square error should be used. For classification problems, the metrics should include sensitivity, specificity, positive predictive value, negative predictive value, area under the ROC curve, and calibration plot. |
|  |  |  |  | ❖ | Validation metrics: calibration (plot, regression intercept and slope, Brier score), utility (decision curve analysis), explainability (clinical assessments based on counterfactual probabilities), and discrimination (the receiver operating characteristics, the area under curve, sensitivity at 95% specificity) |
|  |  |  |  | • | The evaluation metrics were those for assessing the models' calibration, utility, explainability, and discrimination. |
|  |  |  |  | • | To evaluate the model calibration, we assessed (1) a calibration plot with a regression line and histograms of either event or nonevent distribution of the predicted probabilities; (2) the intercept and slope of the linear regression; and (3) the Brier score. |
|  |  |  |  | • | We measured the clinical utility using a decision curve analysis by comparing the net benefits of a model with those if we treated all predictions as either positive (i.e., treat all) or negative (i.e., treat none). |
|  |  |  |  | • | Clinicians (i.e., FZA and AZZAH) assessed the explainability. They were given counterfactual quantities for each predictor in a model. |
|  |  |  |  | • | Eventually, we evaluated the discrimination ability of well-calibrated models by the ROC curve and sensitivity at 95% specificity. |
|  |  |  | ✓ |  | For retrospective studies, split the data into a derivation set and a validation set. For prospective studies, define the starting time for validation data collection. |
|  |  |  |  | ❖ | Data partition: pre-calibration (51.2%), calibration (12.8%), random split (16%), geographical and temporal splits (20%) |
|  |  |  |  | • | This study split the dataset into internal and external validation sets. The latter was by random and non-random sampling. We applied geographical and temporal splitting, i.e., respectively, by cities and days for stratification in non-random sampling, as extensively described in the protocol. These two splitting methods constituted ~20% of the dataset. Subsequently, we applied random sampling to split out ~20% of the remainder of the dataset. This sampling constructed another subset for external validation. For simplicity, external validation aggregately used all subsets by these three splitting methods. Only ~64% of the dataset was for internal validation. For recalibration, we used ~20% of the internal validation set. |
| 8 | Methods | Build the predictive model | ✓ |  | Identify independent variables that predominantly take a single value (e.g., being zero 99% of the time) |
|  |  |  |  | ❖ | Number of independent variables with zero variance: 2 of 102 |
|  |  |  |  | • | Supplemental spreadsheet: Candidate predictors with non-zero variances. |
|  |  |  |  | • |  |
|  |  |  | ✓ |  | Identify and remove redundant independent variables |
|  |  |  |  | ❖ | Number of redundant independent variables: 2 of 100 |
|  |  |  |  | • | Supplemental spreadsheet: Pair-wise Pearson correlations to identify redundant candidate predictors. |
|  |  |  | ✓ |  | Identify the independent variables that may suffer from the perfect separation problem |
|  |  |  |  | ❖ | Perfect-separation variables: 33 of 98 |
|  |  |  |  | • | Supplemental spreadsheet: Candidate predictors with non-zero variances. |
|  |  |  | ✓ |  | Report the number of independent variables, the number of positive examples, and the number of negative examples |
|  |  |  |  | ❖ | Number of independent variables: 54 of 65 (those which leak outcome were removed), in which were used in RR (4 of 54), PC-ENR (14 of 54), PC-RF (5 of 54), PC-GBM (5 of 54), DI-VNN (54 of 54) |
|  |  |  |  | • | Supplemental spreadsheet: Excluded codes in the training set that may leak outcome information. |
|  |  |  | ✓ |  | Assess whether sufficient data are available for a good fit of the model. In particular, for classification, there should be a sufficient number of observations in both positive and negative classes. |
|  |  |  |  | ❖ | Classification problem: 371 events and 58,894 nonevents (EPVs: 92 [RR], 26 [PC-ENR], 74 [PC-RF], 74 [PC-GBM], 6 [DI-VNN]) |
|  |  |  |  | • | Main figure: Subject selection by applying a retrospective design and data partitioning for internal and external validations. |
|  |  |  | ✓ |  | Determine a set of candidate modelling techniques (e.g., logistic regression, random forest, or deep learning). If only one type of model was used, justify the decision for using that model. |
|  |  |  |  | ❖ | Candidate modelling techniques: ridge regression using latent candidate predictors, elastic net regression (ENR) using principal components (PCs), random forest and gradient boosting machine using PCs selected by ENR, and deep-insight visible neural network using all candidate predictors |
|  |  |  |  | • | The first was a statistical machine learning model using pre-selected latent candidate predictors and applying ridge regression (RR). The pre-selection of latent candidate predictors was by multivariate analyses. The second to fourth models were computational machine learning models using 54 candidate predictors transformed into principal components (PCs). It was by 10-fold cross-validation and used no outcome information, as described in the protocol. For candidate predictors, we only used PCs with top proportions of variance explained such that the model training used at least ~20 events per candidate predictor. We applied three algorithms using these PCs: (1) elastic net regression (PC-ENR); (2) random forest (PC-RF); and (3) gradient boosting machine (PC-GBM). The fifth model was a deep-insight visible neural network (DI-VNN). However, unlike the protocol, we did not limit this model to only 22 of 54 candidate predictors, which had a false discovery rate of ≤5% based on differential analyses with Benjamini-Hochberg multiple testing corrections. Instead, we used all 54 candidate predictors considering the feasibility of constructing the data-driven network architecture. In addition, all model recalibration was by either a logistic regression or a general additive model using locally weighted scatterplot smoothing. The recalibration procedure also differed from the protocol. This is because the models only sometimes resulted in a wide range of predicted probabilities, as required for recalibration. |
|  |  |  | ✓ |  | Define the performance metrics to select the best model |
|  |  |  |  | ❖ | Performance metrics for model selection: calibration (plot, regression intercept and slope, Brier score), utility (decision curve analysis), explainability (clinical assessments based on counterfactual probabilities), and discrimination (the receiver operating characteristics, the area under curve, sensitivity at 95% specificity) |
|  |  |  |  | • | The evaluation metrics were those for assessing the models' calibration, utility, explainability, and discrimination. |
|  |  |  |  | • | To evaluate the model calibration, we assessed (1) a calibration plot with a regression line and histograms of either event or nonevent distribution of the predicted probabilities; (2) the intercept and slope of the linear regression; and (3) the Brier score. |
|  |  |  |  | • | We measured the clinical utility using a decision curve analysis by comparing the net benefits of a model with those if we treated all predictions as either positive (i.e., treat all) or negative (i.e., treat none). |
|  |  |  |  | • | Clinicians (i.e., FZA and AZZAH) assessed the explainability. They were given counterfactual quantities for each predictor in a model. |
|  |  |  |  | • | Eventually, we evaluated the discrimination ability of well-calibrated models by the ROC curve and sensitivity at 95% specificity. |
|  |  |  | ✓ |  | Specify the model selection strategy. Common methods include K-fold validation or bootstrap to estimate the lost function on a grid of candidate parameter values. For K-fold validation, proper stratification by the response variable is needed. |
|  |  |  |  | ❖ | Model selection strategy: five-fold cross-validation, bootstrapping |
|  |  |  |  | • | Before recalibration, we applied two resampling methods to evaluate the predictive performance. These were (1) five-fold cross-validation for hyperparameter tuning and (2) bootstrapping for the final training. |
|  |  |  | ✓ |  | (A desirable but not mandatory item) For model selection, include discussion on (1) balance between model accuracy and model simplicity or interpretability, and (2) the familiarity with the modelling techniques of the end user |
|  |  |  |  | ❖ | Modelling balance between accuracy and interpretability: only well-calibrated models were assessed for utility, explainability, and discrimination |
|  |  |  |  | ❖ | Familiarity with the modelling techniques: regression models were included due their familiarity |
|  |  |  |  | • | The evaluation metrics were those for assessing the models' calibration, utility, explainability, and discrimination. |
| 9 | Results | Report the final model and performance | ✓ |  | Report the predictive performance of the final model in terms of the validation metrics specified in the methods section |
|  |  |  |  | ❖ | Predictive performance report: as reported in the figures |
|  |  |  |  | • | Main figure: Model calibration (a) and clinical utility (b). |
|  |  |  |  | • | Main figure: Model discrimination by receiver operating characteristics (ROC) curves. |
|  |  |  |  | • | Main figure: Model discrimination by the area under the receiver operating characteristics curves (AUROCs). |
|  |  |  | ✓ |  | If possible, report the parameter estimates in the model and their confidence intervals. When the direct calculation of confidence intervals is not possible, report nonparametric estimates from bootstrap samples. |
|  |  |  |  | ❖ | Parameter estimates: counterfactual probabilities |
|  |  |  |  | • | In addition, we also show the models' parameters and all counterfactual probabilities |
|  |  |  | ✓ |  | Comparison with other models in the literature should be based on confidence intervals |
|  |  |  |  | ❖ | Other models in the literature: 52 models from 27 previous studies |
|  |  |  |  | • | Main figure: Model discrimination by the area under the receiver operating characteristics curves (AUROCs). |
|  |  |  |  | • | Supplemental spreadsheet: Extracted data of available evaluation metrics for comparable models. |
|  |  |  | ✓ |  | Interpretation of the final model. If possible, report what variables were shown to be predictive of the response variable. State which subpopulation has the best prediction and which subpopulation is most difficult to predict. |
|  |  |  |  | ❖ | Predictive variables: as described in the table |
|  |  |  |  | ❖ | Subpopulation with the best prediction: - |
|  |  |  |  | ❖ | Subpopulation that was most difficult to predict: - |
|  |  |  |  | • | Main table: Model explainability by clinical assessments based on counterfactual probabilities. |
| 10 | Discussion | Clinical implications | ✓ |  | Report the clinical implications derived from the obtained predictive performance. For example, report the dollar amount that could be saved with better prediction. How many patients could benefit from a care model leveraging the model prediction? And to what extent? |
|  |  |  |  | ❖ | Potential cost efficiency: achievable in resource-limited settings |
|  |  |  |  | ❖ | Potential healthcare impact: wider coverage of target population |
|  |  |  |  | • | One of the strengths of this study were no requirements from our models, including the DI-VNN, for either ultrasound or biomarker measurements to predict FGR/SGA in advance. We could apply our models to a general population of pregnant women. Furthermore, our model did not require a specific gestational age range for computing the predicted probability. |
| 11 | Discussion | Limitations of the model | ✓ |  | Discuss the following potential limitations: • Assumed input and output data format; • (Desirable but not mandatory items) Potential pitfalls in interpreting the model; • Potential bias of the data used in modelling; • Generalizability of the data |
|  |  |  |  | ❖ | Assumed input and output data format: require electronic health records |
|  |  |  |  | ❖ | Potential pitfalls in interpreting the model: require an impact study |
|  |  |  |  | ❖ | Potential bias of the data used in modelling: require an impact study |
|  |  |  |  | ❖ | Generalizability of the data: require an impact study |
|  |  |  |  | • | The predictive performance of the best model, i.e., the DI-VNN, was considerably moderate according to the AUROC as was the sensitivity at 95% specificity using an internal validation set. However, previous models also achieved similar predictive performances. Another limitation was that medical histories from electronic health records might take time to execute; yet, this is considerably more achievable in many settings. It still needs to be determined if the DI-VNN can improve patient outcomes. |
| 12 | Discussion |  | ✓ |  | (Desirable but not mandatory items) Report unexpected signs of coefficients, indicating collinearity or complex interaction between predictor variables |
|  |  |  |  | ❖ | Unexpected signs of coefficients: as described in the table |
|  |  |  |  | • | Main table: Model explainability by clinical assessments based on counterfactual probabilities. |

Table B2. Prediction model risk of bias assessment tools (PROBAST).

| DOMAIN 1: Participants | | | | | |
| --- | --- | --- | --- | --- | --- |
| Risk of Bias | | | | | |
| *Describe the sources of data and criteria for participant selection:* | | | | | |
|  | • | We applied a retrospective design to select subjects from a public dataset version 2 (August 2019; access approval no.: 510/PPID/1223) of a nationwide health insurance database in Indonesia. | | | |
|  | • | The inclusion criteria were females aged 12 to 55 years who had visited primary, secondary, or tertiary care facilities. All visits afterward were excluded if a woman was pregnant and had a delivery. | | | |
|  | • | This study split the dataset into internal and external validation sets. The latter was by random and non-random sampling. We applied geographical and temporal splitting, i.e., respectively, by cities and days for stratification in non-random sampling, as extensively described in the protocol. These two splitting methods constituted ~20% of the dataset. Subsequently, we applied random sampling to split out ~20% of the remainder of the dataset. This sampling constructed another subset for external validation. For simplicity, external validation aggregately used all subsets by these three splitting methods. Only ~64% of the dataset was for internal validation. For recalibration, we used ~20% of the internal validation set. | | | |
|  | |  | | Dev | Val |
| 1.1  Were appropriate data sources used, e.g. cohort, RCT or nested case-control study data? | | | | Y | Y |
| 1.2  Were all inclusions and exclusions of participants appropriate? | | | | Y | Y |
| Risk of bias introduced by selection of participants | | | RISK: | Low | Low |
|  |  |  | *(low/ high/ unclear)* |  |  |
| *Rationale of bias rating:* | | | | | |
|  | ❖ | Retrospective cohort | | | |
|  | ❖ | As intended to nationwide insured women | | | |

| DOMAIN 2: Predictors | | | | | |
| --- | --- | --- | --- | --- | --- |
| Risk of Bias | | | | | |
| *List and describe predictors included in the final model, e.g. definition and timing of assessment:* | | | | | |
|  | • | Candidate predictors were only medical histories of diagnoses and procedures. These were either single or multiple ICD-10 codes. As extensively described in the protocol, the preprocessing of candidate predictors consisted of (1) preventing zero variance, perfect separation and leakage of the outcome, and redundant predictors; (2) simulating real-world data; and (3) systematically determining the multiple ICD-10 codes for defining latent candidate predictors based on prior knowledge. | | | |
|  | |  | | Dev | Val |
| 2.1  Were predictors defined and assessed in a similar way for all participants? | | | | PY | PY |
| 2.2 Were predictor assessments made without knowledge of outcome data? | | | | Y | Y |
| 2.3  Are all predictors available at the time the model is intended to be used? | | | | Y | Y |
| Risk of bias introduced by predictors or their assessment | | | RISK: | Low | Low |
|  |  |  | *(low/ high/ unclear)* |  |  |
| *Rationale of bias rating:* | | | | | |
|  | ❖ | International standard, as used in real-world application by clinicians and professional coders. | | | |
|  | ❖ | All the codes which leak the outcome were removed. | | | |
|  | ❖ | Feature extraction only took code encounters up to the prediction date. | | | |

| DOMAIN 3: Outcome | | | | | |
| --- | --- | --- | --- | --- | --- |
| Risk of Bias | | | | | |
| *Describe the outcome, how it was defined and determined, and the time interval between predictor assessment and outcome determination:* | | | | | |
|  | • | The event outcome definition in this study utilized the International Classification of Disease version 10 (ICD-10) codes. These were codes preceded by either O365 (maternal care for known or suspected fetal growth) or P05 (disorders of newborns related to slow fetal growth and fetal malnutrition). Both codes indicating FGR and SGA were assigned with those respectively for mothers and fetuses/newborns. A nonevent outcome was assigned if the end of pregnancy was identified within the dataset period by the codes for determining delivery. Otherwise, we assigned an outcome to a censored one. | | | |
|  | • | Candidate predictors were only medical histories of diagnoses and procedures. These were either single or multiple ICD-10 codes. As extensively described in the protocol, the preprocessing of candidate predictors consisted of (1) preventing zero variance, perfect separation and leakage of the outcome, and redundant predictors; (2) simulating real-world data; and (3) systematically determining the multiple ICD-10 codes for defining latent candidate predictors based on prior knowledge. | | | |
|  | • |  | | | |
|  | • |  | | | |
|  | • |  | | | |
|  | • |  | | | |
|  | • |  | | | |
|  | |  | | Dev | Val |
| 3.1  Was the outcome determined appropriately? | | | | Y | Y |
| 3.2  Was a pre-specified or standard outcome definition used? | | | | Y | Y |
| 3.3  Were predictors excluded from the outcome definition? | | | | Y | Y |
| 3.4  Was the outcome defined and determined in a similar way for all participants? | | | | PY | PY |
| 3.5  Was the outcome determined without knowledge of predictor information? | | | | Y | Y |
| 3.6  Was the time interval between predictor assessment and outcome determination appropriate? | | | | Y | Y |
| Risk of bias introduced by the outcome or its determination | | | RISK: | Low | Low |
|  |  |  | *(low/ high/ unclear)* |  |  |
| *Rationale of bias rating:* | | | | | |
|  | ❖ | International standard, as used in real-world application by clinicians and professional coders. | | | |
|  | ❖ | All the codes which leak the outcome were removed. | | | |
|  | ❖ | Feature extraction only took code encounters up to the prediction date, as we developed prognostic prediction models. | | | |

| DOMAIN 4: Analysis | | | | | |
| --- | --- | --- | --- | --- | --- |
| Risk of Bias | | | | | |
| *Describe numbers of participants, number of candidate predictors, outcome events and events per candidate predictor:* | | | | | |
|  | • | Main figure: Subject selection by applying a retrospective design and data partitioning for internal and external validations. | | | |
|  | • | Supplemental spreadsheet: Candidate predictors with non-zero variances. | | | |
| *Describe how the model was developed (for example in regards to modelling technique (e.g. survival or logistic modelling), predictor selection, and risk group definition):* | | | | | |
|  | • | The event outcome definition in this study utilized the International Classification of Disease version 10 (ICD-10) codes. These were codes preceded by either O365 (maternal care for known or suspected fetal growth) or P05 (disorders of newborns related to slow fetal growth and fetal malnutrition). Both codes indicating FGR and SGA were assigned with those respectively for mothers and fetuses/newborns. A nonevent outcome was assigned if the end of pregnancy was identified within the dataset period by the codes for determining delivery. Otherwise, we assigned an outcome to a censored one. | | | |
|  | • | The first was a statistical machine learning model using pre-selected latent candidate predictors and applying ridge regression (RR). The pre-selection of latent candidate predictors was by multivariate analyses. The second to fourth models were computational machine learning models using 54 candidate predictors transformed into principal components (PCs). It was by 10-fold cross-validation and used no outcome information, as described in the protocol. For candidate predictors, we only used PCs with top proportions of variance explained such that the model training used at least ~20 events per candidate predictor. We applied three algorithms using these PCs: (1) elastic net regression (PC-ENR); (2) random forest (PC-RF); and (3) gradient boosting machine (PC-GBM). The fifth model was a deep-insight visible neural network (DI-VNN). However, unlike the protocol, we did not limit this model to only 22 of 54 candidate predictors, which had a false discovery rate of ≤5% based on differential analyses with Benjamini-Hochberg multiple testing corrections. Instead, we used all 54 candidate predictors considering the feasibility of constructing the data-driven network architecture. In addition, all model recalibration was by either a logistic regression or a general additive model using locally weighted scatterplot smoothing. The recalibration procedure also differed from the protocol. This is because the models only sometimes resulted in a wide range of predicted probabilities, as required for recalibration. | | | |
| *Describe whether and how the model was validated, either internally (e.g. bootstrapping, cross validation, random split sample) or externally (e.g. temporal validation, geographical validation, different setting, different type of participants):* | | | | | |
|  | • | This study split the dataset into internal and external validation sets. The latter was by random and non-random sampling. We applied geographical and temporal splitting, i.e., respectively, by cities and days for stratification in non-random sampling, as extensively described in the protocol. These two splitting methods constituted ~20% of the dataset. Subsequently, we applied random sampling to split out ~20% of the remainder of the dataset. This sampling constructed another subset for external validation. For simplicity, external validation aggregately used all subsets by these three splitting methods. Only ~64% of the dataset was for internal validation. For recalibration, we used ~20% of the internal validation set. Before recalibration, we applied two resampling methods to evaluate the predictive performance. These were (1) five-fold cross-validation for hyperparameter tuning and (2) bootstrapping for the final training. | | | |
| *Describe the performance measures of the model, e.g. (re)calibration, discrimination, (re)classification, net benefit, and whether they were adjusted for optimism:* | | | | | |
|  | • | The evaluation metrics were those for assessing the models' calibration, utility, explainability, and discrimination. | | | |
|  | • | To evaluate the model calibration, we assessed (1) a calibration plot with a regression line and histograms of either event or nonevent distribution of the predicted probabilities; (2) the intercept and slope of the linear regression; and (3) the Brier score. | | | |
|  | • | We measured the clinical utility using a decision curve analysis by comparing the net benefits of a model with those if we treated all predictions as either positive (i.e., treat all) or negative (i.e., treat none). | | | |
|  | • | Clinicians (i.e., FZA and AZZAH) assessed the explainability. They were given counterfactual quantities for each predictor in a model. | | | |
|  | • | Eventually, we evaluated the discrimination ability of well-calibrated models by the ROC curve and sensitivity at 95% specificity. | | | |
| *Describe any participants who were excluded from the analysis:* | | | | | |
|  | • | The event outcome definition in this study utilized the International Classification of Disease version 10 (ICD-10) codes. These were codes preceded by either O365 (maternal care for known or suspected fetal growth) or P05 (disorders of newborns related to slow fetal growth and fetal malnutrition). Both codes indicating FGR and SGA were assigned with those respectively for mothers and fetuses/newborns. A nonevent outcome was assigned if the end of pregnancy was identified within the dataset period by the codes for determining delivery. Otherwise, we assigned an outcome to a censored one. | | | |
| *Describe missing data on predictors and outcomes as well as methods used for missing data:* | | | | | |
|  | • | As extensively described in the protocol, the preprocessing of candidate predictors consisted of (1) preventing zero variance, perfect separation and leakage of the outcome, and redundant predictors; (2) simulating real-world data; and (3) systematically determining the multiple ICD-10 codes for defining latent candidate predictors based on prior knowledge. | | | |
|  | |  | | Dev | Val |
| 4.1  Were there a reasonable number of participants with the outcome? | | | | Y | Y |
| 4.2  Were continuous and categorical predictors handled appropriately? | | | | Y | Y |
| 4.3  Were all enrolled participants included in the analysis? | | | | PY | PY |
| 4.4  Were participants with missing data handled appropriately? | | | | PY | PY |
| 4.5 Was selection of predictors based on univariable analysis avoided? | | | | Y |  |
| 4.6  Were complexities in the data (e.g. censoring, competing risks, sampling of controls) accounted for appropriately? | | | | PY | PY |
| 4.7  Were relevant model performance measures evaluated appropriately? | | | | Y | Y |
| 4.8  Were model overfitting and optimism in model performance accounted for? | | | | Y |  |
| 4.9 Do predictors and their assigned weights in the final model correspond to the results from multivariable analysis? | | | | PY |  |
| Risk of bias introduced by the analysis | | | RISK: | Low | Low |
|  |  |  | *(low/ high/ unclear)* |  |  |
| *Rationale of bias rating:* | | | | | |
|  | ❖ | Internal validation: 371 events and 58,894 nonevents. Number of candidate predictors: RR (4 of 54), PC-ENR (14 of 54), PC-RF (5 of 54), PC-GBM (5 of 54), DI-VNN (54 of 54). Events per variable: 92 [RR], 26 [PC-ENR], 74 [PC-RF], 74 [PC-GBM], 6 [DI-VNN]. External validation: 472 events and 56,012 nonevents. | | | |
|  | ❖ | No continuous predictors | | | |
|  | ❖ | Excluded instances due missing outcome were taken into account by inverse probability weighting. | | | |
|  | ❖ | Univariate analysis was avoided. Instead, we applied multivariate analysis and dimensional reduction (excluding outcome). | | | |
|  | ❖ | Censoring was handled by inverse probability weighting. Competing risks were included in the predictors. There was no sampling of controls since we applied retrospective cohort paradigm. | | | |
|  | ❖ | Calibration, net benefit, and discriminative performances were evaluated, including calibration plot and measures, and the area under curve of the receiver operating characteristics. | | | |
|  | ❖ | Resampling techniques and geographical/temporal splits were applied to avoid overoptimistic evaluation. | | | |
|  | ❖ | We only included latent candidate predictors which were selected by multivariate analysis. | | | |
|  | ❖ | Missing value handling by historical rates (modified Kaplan-Meier estimators) for predictors and inverse probability weighting including missing values for outcome | | | |

| Overall judgement about risk of bias of the prediction model evaluation | | |
| --- | --- | --- |
| Overall judgement of risk of bias | RISK: | Low |
|  | *(low/ high/ unclear)* |  |
| *Summary of sources of potential bias:* | | |
| - | | |

Table B3. Clinical checklists for assessing the suitability of machine learning applications in healthcare.

| Item | | Response | |
| --- | --- | --- | --- |
| Q1. | What is the purpose and context of the algorithm? |  | A prediction model for use in resource-limited settings |
|  |  |  | A preliminary prediction model before ordering advanced predictor measurements |
| Q2. | How good were the data used to train the algorithm? |  |  |
| Q2a. | To what extent were the data accurate and free of bias? |  | This algorithm is accurate for 12-to-55-year-old females that visit primary, secondary, or tertiary care. |
| Q2b. | Were data labelled correctly? |  | Data were labelled as for real-world use. |
| Q2c. | Were the data standardized and interoperable? |  | All predictors used the International Classification of Disease (ICD)-10. |
| Q3. | Were there sufficient data to train the algorithm? |  | This algorithm had less data for its training (i.e., 6 events per predictor), compared to those for the comparator training, but DI-VNN was more accepted by clinical perspectives and externally validated. |
| Q4. | How well does the algorithm perform? |  | DI-VNN had comparable predictive performance by external validation and systematic review and meta-analysis of 52 models from 27 previous studies. |
| Q5. | Is the algorithm transferable to new clinical settings? |  | This algorithm is transferable to settings in which ICD-10 codes can be obtained. |
| Q6. | Are the outputs of the algorithm clinically intelligible? |  | In DI-VNN, clinicians could explore how the outputs were obtained for an individual. |
| Q7. | How will this algorithm fit into and complement current workflows? |  | This algorithm should be a part of automatic screening in electronic health record system. |
|  |  |  | A prediction is made each time a patient have a visit and been encountered the codes of diagnoses and procedures for that visit. |
| Q8. | Has use of the algorithm been shown to improve patient care and outcomes? |  | Unclear yet, but we provide a publicly-accessed, free web application for future studies to investigate the impact of this algorithm. |
| Q9. | Could the algorithm cause patient harm? |  | Under-prognosis could cause a subject to lose the chance for FGR/SGA prevention, which may lead to neonatal morbidity and mortality with higher healthcare costs. Meanwhile, over-prognosis may lead to higher healthcare costs by ordering more tests to confirm the risk of FGR/SGA. |
| Q10. | Does the algorithm raise ethical, legal or social concerns? |  | None of the predictors used socioeconomic variables and all the categories were well represented in the data used to train this algorithm; thus, we can expect a fair prediction. |
|  |  |  | This algorithm does not need to access medical histories from other healthcare facilities; thus, there is no privacy issue. |

Table B4. Preferred reporting items for systematic reviews and meta-analyses (PRISMA) 2020 expanded checklist.

| Section and topic | Item # | Elements recommended for reporting | |
| --- | --- | --- | --- |
| Eligibility criteria | 5 | ❖ | Population: either pregnant or non-pregnant women without specifying medical conditions |
|  |  | ❖ | Index: prediction models from this study |
|  |  | ❖ | Comparator: prediction models or rules from previous studies |
|  |  | ❖ | Outcome: either FGR or SGA as a binary variable (i.e., event vs. nonevent) with ≥20 events per variable/predictor (EPVs) |
|  |  | ❖ | Time: from days to weeks before the outcome (i.e., long prognostication) |
|  |  | ❖ | Setting: either primary care or hospital |
| Information sources | 6 | ❖ | PubMed |
|  |  | ❖ | Scopus |
|  |  | ❖ | Web of Science |
| Search strategy | 7 | ❖ | The search filter included all original articles and systematic reviews and excluded other article types, e.g., conference abstracts, but we included conference papers. We searched for studies published up to April 3, 2021, in three literature databases within the last 5 years. Keywords were *fetal growth restriction* or *intrauterine growth retardation* and *small for gestational age*, combined with prognostic prediction. |
|  |  | ❖ | Only three studies fulfilled the eligibility criteria from three literature databases within the last 5 years. All of the studies were systematic reviews. Thus, we also searched eligible articles in the systematic reviews, including those published more than 5 years earlier. |
| Selection process | 8 | ❖ | Articles were searched and loosely filtered by author HS using titles and abstracts. HS subsequently assessed the eligibility of the filtered articles by examining full text. This step could find articles with ambiguous eligibility, which were also independently assessed by author YWW. If HS and YWW disagreed on an article, author ECYS made the final decision. |
| Data collection process | 9 | ❖ | HS extracted data of the best model from each eligible article, for the outcome with the most similar definition to the one in this study. If an article did not report the area under curve of the receiver operatic characteristics (AUROC), we calculated it by the trapezoidal rule utilizing the sensitivity and specificity. If a study used more than one validation technique, the extracted metrics were those estimated by the most recommended technique. External validation was considerably better than internal validation. For the latter, bootstrapping was considerably better than cross-validation, yet, it was still better than test split. |
| Data items (outcomes) | 10a | ❖ | Sensitivity |
|  |  | ❖ | Specificity |
|  |  | ❖ | AUROC |
| Data items (other variables) | 10b | ❖ | Other extracted information included (1) study design; (2) population; (3) setting; (4) outcome definition; (5) sample size; (6) details on events and nonevents; (7) number of candidate predictors; (8) EPVs; (9) predictors in the final model; and (10) validation technique. |
| Effect measures | 12 | ❖ | Plotting sensitivities and specificities on the receiver operating characteristics |
|  |  | ❖ | AUROCs with 95% confidence intervals |

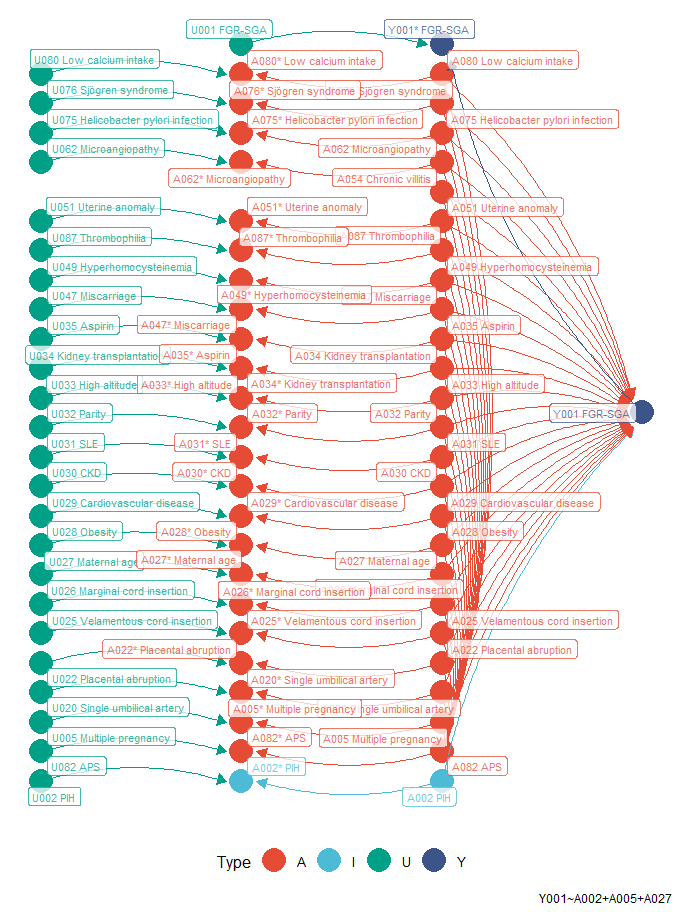

Figure B1. Association diagram of pregnancy-induced hypertension and FGR/SGA. Type A is confounder. Type I is variable of interest. Asterisk (*) symbol indicates measured variable. Type U is any unmeasured variables which affect measurement of variable. APS, anti-phospholipid syndrome; CKD, chronic kidney disease; FGR, fetal growth restriction; PIH, pregnancy-induced hypertension; SGA, small for gestational age; SLE, systemic lupus erythematosus.

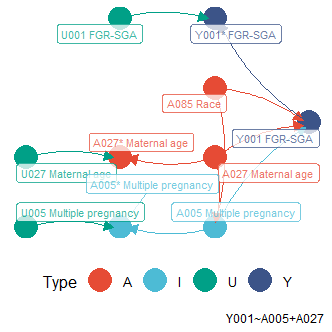

Figure B2. Association diagram of multiple pregnancies and FGR/SGA. Type A is confounder. Type I is variable of interest. Asterisk (*) symbol indicates measured variable. Type U is any unmeasured variables which affect measurement of variable. FGR, fetal growth restriction; SGA, small for gestational age.

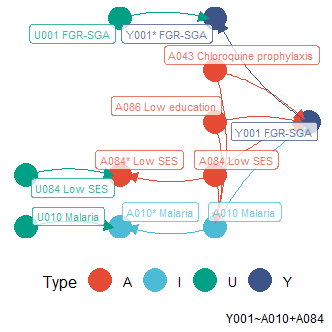

Figure B3. Association diagram of malaria and FGR/SGA. Type A is confounder. Type I is variable of interest. Asterisk (*) symbol indicates measured variable. Type U is any unmeasured variables which affect measurement of variable. FGR, fetal growth restriction; SES, socioeconomic status; SGA, small for gestational age.

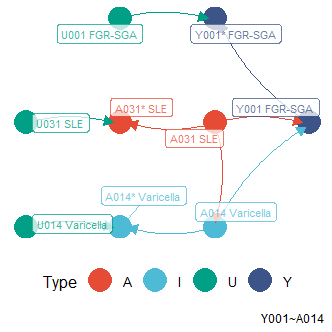

Figure B4. Association diagram of varicella and FGR/SGA. Type A is confounder. Type I is variable of interest. Asterisk (*) symbol indicates measured variable. Type U is any unmeasured variables which affect measurement of variable. FGR, fetal growth restriction; SGA, small for gestational age; SLE, systemic lupus erythematosus.

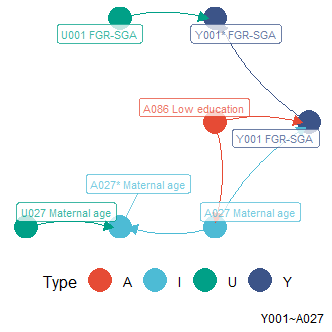

Figure B5. Association diagram of risk of an adverse pregnancy by maternal age and FGR/SGA. Type A is confounder. Type I is variable of interest. Asterisk (*) symbol indicates measured variable. Type U is any unmeasured variables which affect measurement of variable. FGR, fetal growth restriction; SGA, small for gestational age.

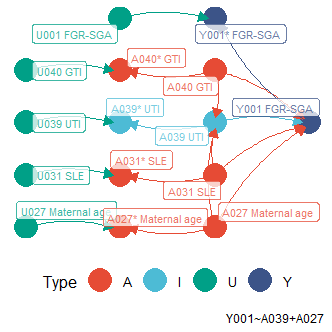

Figure B6. Association diagram of UTI and FGR/SGA. Type A is confounder. Type I is variable of interest. Asterisk (*) symbol indicates measured variable. Type U is any unmeasured variables which affect measurement of variable. FGR, fetal growth restriction; GTI, genital tract infections; SGA, small for gestational age; SLE, systemic lupus erythematosus; UTI, urinary tract infections.

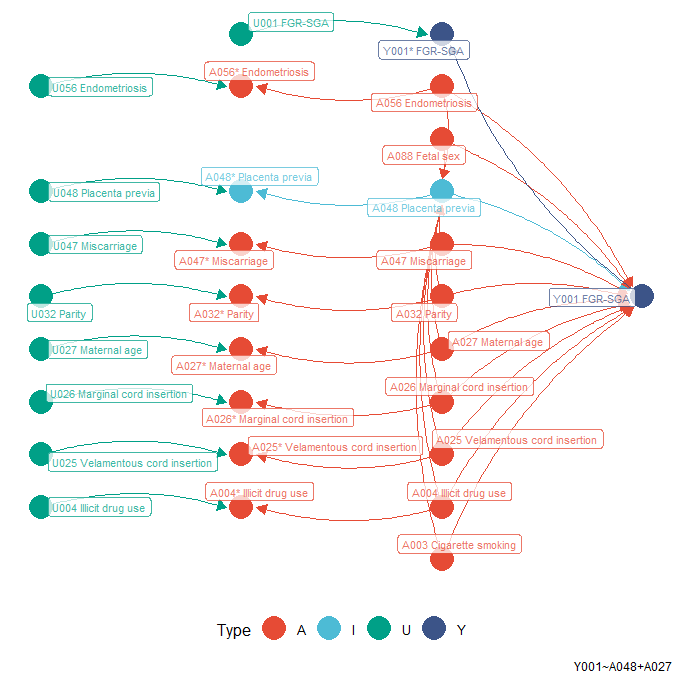

Figure B7. Association diagram of placenta previa and FGR/SGA. Type A is confounder. Type I is variable of interest. Asterisk (*) symbol indicates measured variable. Type U is any unmeasured variables which affect measurement of variable. FGR, fetal growth restriction; SGA, small for gestational age.

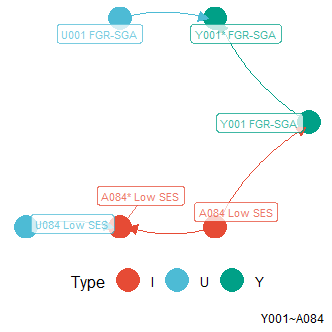

Figure B8. Association diagram of a low SES and FGR/SGA. Type A is confounder. Type I is variable of interest. Asterisk (*) symbol indicates measured variable. Type U is any unmeasured variables which affect measurement of variable. FGR, fetal growth restriction; SES, socioeconomic status; SGA, small for gestational age.

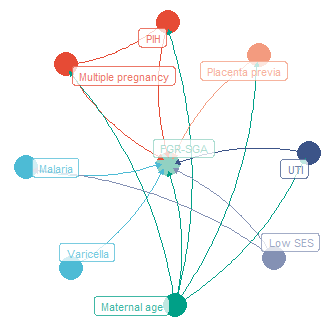

Figure B9. Unified association diagram between latent candidate predictors and FGR/SGA, which had significant associations. Yet, those, as visualized by the inter-predictor edges, were not tested. Colors are intended to easily identify nodes and edges. FGR, fetal growth restriction; PIH, pregnancy-induced hypertension; SES, socioeconomic status; SGA, small for gestational age; UTI, urinary tract infection.

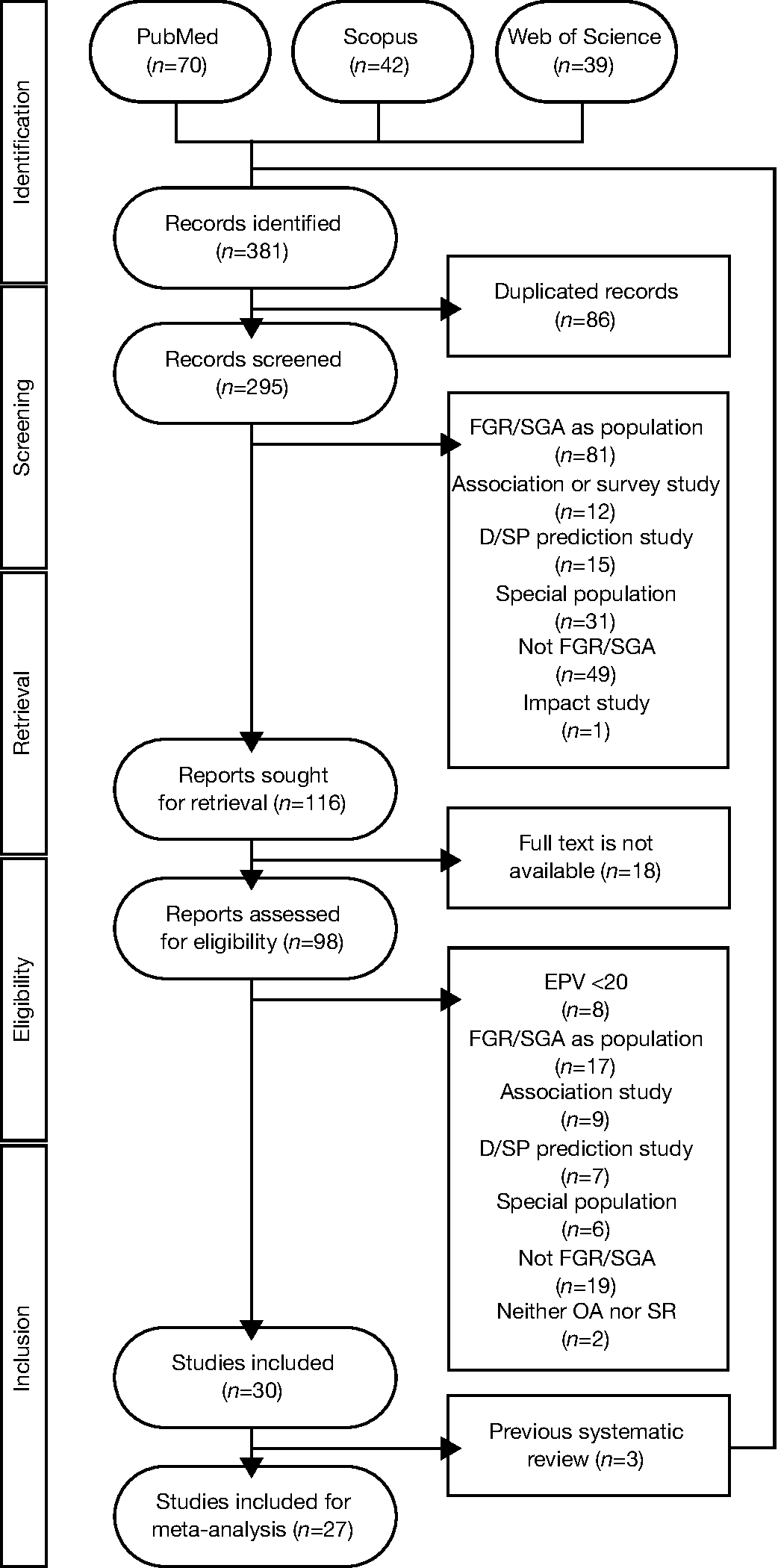

Figure B10. Flow diagram to find comparable models from previous studies. Records indirectly identified from previous systematic reviews were added after those directly identified from literature databases. The numbers indicate the overall results. D/SP, diagnostic or short prognostic; FGR, fetal growth restriction; *n*, number of records/reports/studies; OA, original article; SGA, small for gestational age; SR, systematic review.
