## Appendix D for "Widely accessible prognostication using medical history for fetal growth restriction and small for gestational age in nationwide insured women"

Table D1. Search and filter results of previous studies.

| Section | Inclusion |  | Exclusion |  |
| --- | --- | --- | --- | --- |
|  | Step | Studies ^a^ | Step | Studies ^a^ |
| Identification | Records identified | [1-295] |  |  |
| Screening | Records screened | [1-295] | Duplicated records | (not applicable) ^a^ |
| Retrieval | Reports sought for retrieval ^b^ | [1-116] | FGR/SGA as population | [117-197] |
|  |  |  | Association or survey study | [198-209] |
|  |  |  | Diagnostic or short prognostic study | [210-214] |
|  |  |  | Special population | [215-245] |
|  |  |  | Not FGR/SGA | [246-294] |
|  |  |  | Impact study | [295] |
| Eligibility | Reports assessed for eligibility | [1-98] | Full text is not available | [99-116] |
| Inclusion | Studies included | [1-30] | EPV <20 | [31-38] |
|  |  |  | FGR/SGA as population | [39-55] |
|  |  |  | Association study | [56-64] |
|  |  |  | Diagnostic or short prognostic study | [65-71] |
|  |  |  | Special population | [72-77] |
|  |  |  | Not FGR/SGA | [78-96] |
|  |  |  | Neither original article nor systematic review | [97,98] |
|  | Studies included for meta-analysis | [1-27] | Previous systematic review | [28], ^c^ [29], ^d^ [30], ^e^ |

^a^, duplicates removed; ^b^, no abstract [12,102,103,105,106,110,112-115]; ^c^, source for [1-15,38,92-94,197,202-209,291-294] ; ^d^, source for [16-23,34-37,41-55,63,64,66-71,76,77,84-91,102-116,123-196,199-201,211-214,229-245,262-290]; ^e^, source for [24-27,31-33,75,101]; FGR, fetal growth restriction; SGA, small-for-gestational age.

Table D2. Comparable models to evaluate the success criteria.

| Study/model | Metrics (95% CI) | Other variables | |
| --- | --- | --- | --- |
| [1] Yaron Y *et al.* (2002)  PAPP-A ≤0.5 MoM | Sensitivity: |  | Study design: retrospective cohort |
|  | 0.37 |  | Population: pregnant women at 11–13 weeks’ gestation, singleton, and underwent combined first trimester screening for trisomy 21 |
|  | Specificity: |  | Setting: hospital |
|  | 0.86 |  | Outcome definition: event (FGR/SGA, birth weight <5^th^ percentile) and non-event (not FGR/SGA with or without other complications) |
|  | AUROC:* |  | Sample size: event (*n*=49) and non-event (*n*=1573) |
|  | 0.612 |  | Predictors in the best final model: PAPP-A |
|  |  |  | The most recommended validation technique: no internal validation |
| [2] Tul N *et al.* (2003)  PAPP-A ≤0.5 MoM | Sensitivity: |  | Study design: prospective cohort |
|  | 0.2 |  | Population: pregnant women, singleton, and underwent combined first trimester screening for trisomy 21 |
|  | Specificity: |  | Setting: hospital |
|  | 0.92 |  | Outcome definition: event (FGR/SGA, birth weight <10^th^ percentile) and non-event (not FGR/SGA without other complications) |
|  | AUROC:* |  | Sample size: event (*n*=51) and non-event (*n*=953) |
|  | 0.557 |  | Predictors in the best final model: PAPP-A |
|  |  |  | The most recommended validation technique: no internal validation |
| [3] Spencer K *et al.* (2005)  Log_10_ PAPP-A MoM and log10 UtA-PI MoM | Sensitivity: |  | Study design: prospective cohort |
|  | 0.16 |  | Population: pregnant women from 11–13 weeks’ gestation, singleton, and underwent combined first trimester screening for trisomy 21 |
|  | Specificity: |  | Setting: hospital |
|  | 0.95 |  | Outcome definition: event (FGR/SGA, birth weight <5^th^ percentile) and non-event (not FGR/SGA with or without other complications) |
|  | AUROC:* |  | Sample size: event (*n*=172) and non-event (*n*=3827) |
|  | 0.555 |  | Predictors in the best final model: PAPP-A at 11–13 weeks’ gestation and UtA-PI at 22–24 weeks’ gestation |
|  |  |  | The most recommended validation technique: no internal validation |
| [4] Smith GC *et al.* (2002)  (a) Lowest 5% of PAPP-A MoM  (b) Lowest 5% of FβhCG MoM | Sensitivity: |  | Study design: prospective cohort |
|  | (a) 0.12; (b) 0.08 |  | Population: pregnant women from 8–14 weeks’ gestation, singleton, underwent combined first trimester screening for trisomy 21, delivered within 24–43 weeks’ gestation, and normal karyotype |
|  | Specificity: |  | Setting: hospitals |
|  | (a) 0.95; (b) 0.95 |  | Outcome definition: event (FGR/SGA, birth weight <5^th^ percentile) and non-event (not FGR/SGA with or without other complications) |
|  | AUROC:* |  | Sample size: event (*n*=370) and non-event (*n*=8469) |
|  | (a) 0.538; (b) 0.515 |  | Predictors in the best final model: (a) PAPP-A; (b) FβhCG |
|  |  |  | The most recommended validation technique: no internal validation |
| [5] Radoi VM and Bohiltea LC (2009)  (a) Lowest 5% of PAPP-A MoM  (b) Lowest 5% of FβhCG MoM | Sensitivity: |  | Study design: prospective cohort |
|  | (a) 0.79; (b) 0.43 |  | Population: pregnant women from 10–13 weeks’ gestation, singleton, underwent combined first trimester screening for trisomy 21, delivered within 24–43 weeks’ gestation, and normal karyotype |
|  | Specificity: |  | Setting: hospital |
|  | (a) 0.59; (b) 0.6 |  | Outcome definition: event (FGR/SGA, birth weight <5^th^ percentile) and non-event (not FGR/SGA with or without other complications) |
|  | AUROC:* |  | Sample size: event (*n*=370) and non-event (*n*=8469) |
|  | (a) 0.687; (b) 0.51 |  | Predictors in the best final model: (a) PAPP-A; (b) FβhCG |
|  |  |  | The most recommended validation technique: no internal validation |
| [6] Pilalis A *et al.* (2007)  (a) PAPP-A ≤ 5^th^ percentile  (b) UtA-PI ≥ 95^th^ percentile  (c) PAPP-A and UtA-PI | Sensitivity: |  | Study design: prospective cohort |
|  | (a) 0.17; (b) 0.23; (c) 0.11 |  | Population: pregnant women from 11–14 weeks’ gestation, singleton, underwent combined first trimester screening for trisomy 21, fetal  crown–rump length of 45–84 mm, and known pregnancy outcome |
|  | Specificity: |  | Setting: hospital |
|  | (a) 0.99; (b) 0.99; (c) 0.99* |  | Outcome definition: event (FGR/SGA, birth weight <5^th^ percentile) and non-event (not FGR/SGA with or without other complications) |
|  | AUROC: |  | Sample size: event (*n*=35) and non-event (*n*=843) |
|  | (a) 0.582*; (b) 0.061*;  (c) 0.689 (0.658 to 0.72)** |  | Predictors in the best final model: (a) PAPP-A; (b) UtA-PI; (c) PAPP-A and UtA-PI |
|  |  |  | The most recommended validation technique: no internal validation |
| [7] Ong CY *et al.* (2000)  (a) PAPP-A ≤ 5^th^ percentile  (b) FβhCG ≤ 5^th^ percentile | Sensitivity: |  | Study design: retrospective cohort |
|  | (a) 0.13; (b) 0.07 |  | Population: pregnant women from 10–14 weeks’ gestation, singleton, and underwent combined first trimester screening for trisomy 21 |
|  | Specificity: |  | Setting: hospital |
|  | (a) 0.95; (b) 0.96 |  | Outcome definition: event (FGR/SGA, birth weight <5^th^ percentile) and non-event (not FGR/SGA with or without other complications) |
|  | AUROC:* |  | Sample size: event (*n*=171) and non-event (*n*=5126) |
|  | (a) 0.539; (b) 0.513 |  | Predictors in the best final model: (a) PAPP-A; (b) FβhCG |
|  |  |  | The most recommended validation technique: no internal validation |
| [8] Leung TY *et al.* (2008)  (a) Log_10_ PAPP-A ≤ 5^th^ percentile  (b) CRL < expected by 3–6 days  (c) Both (a) and (b)  (d) Either (a) or (b) | Sensitivity: |  | Study design: prospective cohort |
|  | (a) 0.09; (b) 0.10;  (c) 0.02; (d) 0.17 |  | Population: pregnant women from 11–14 weeks’ gestation, singleton, underwent combined first trimester screening for trisomy 21, no chromosomal and structural abnormalities, no intrauterine death, and known pregnancy outcome |
|  | Specificity: |  | Setting: hospital |
|  | (a) 0.96; (b) 0.93;  (c) 0.99; (d) 0.89 |  | Outcome definition: event (FGR/SGA, birth weight <10^th^ percentile) and non-event (not FGR/SGA with or without other complications) |
|  | AUROC: |  | Sample size: event (*n*=241) and non-event (*n*=2519) |
|  | (a) 0.608 (0.57 to 0.646) **;  (b) 0.608 (0.57 to 0.646) **; |  | Predictors in the best final model: (a) PAPP-A; (b) CRL; (c) PAPP-A and CRL; (d) PAPP-A and CRL |
|  | (c) 0.509*; (d) 0.528* |  | The most recommended validation technique: no internal validation |
| [9] Krantz D *et al.* (2004)  (a) PAPP-A MoM ≤ 5^th^ percentile and maternal factors  (b) FβhCG MoM ≤ 1^th^ percentile and maternal factors | Sensitivity: |  | Study design: prospective cohort |
|  | (a) 0.1; (b) 0.02 |  | Population: pregnant women from 10–14 weeks’ gestation, singleton, underwent combined first trimester screening for trisomy 21, no trisomy 21 or 18 in previous and observed pregnancy, no early termination/fetal loss, and known pregnancy outcome and birth weight |
|  | Specificity: |  | Setting: hospitals |
|  | (a) 0.96; (b) 0.99 |  | Outcome definition: event (FGR/SGA, birth weight <10^th^ percentile) and non-event (not FGR/SGA with or without other complications) |
|  | AUROC:* |  | Sample size: event (*n*=384) and non-event (*n*=5892) |
|  | (a) 0.529; (b) 0.506 |  | Predictors in the best final model: (a) PAPP-A and maternal factors; (b) FβhCG and maternal factors. Maternal factors: age, weight, race, and cigarrette smoker. |
|  |  |  | The most recommended validation technique: no internal validation |
| [10] Kavak ZN *et al.* (2006)  PAPP-A MoM ≤ 5^th^ percentile | Sensitivity: |  | Study design: prospective cohort |
|  | 0.11 |  | Population: pregnant women, singleton, underwent combined first trimester screening for trisomy 21, no chromosomal and structural abnormalities, no chronic hypertension and previously diagnosed diabetes, and no previous preeclampsia, gestational hypertension, gestational diabetes, and FGR |
|  | Specificity: |  | Setting: hospital |
|  | 0.95 |  | Outcome definition: event (FGR/SGA, birth weight <10^th^ percentile) and non-event (not FGR/SGA with or without other complications) |
|  | AUROC: |  | Sample size: event (*n*=35) and non-event (*n*=441) |
|  | 0.61 (0.5 to 0.71)** |  | Predictors in the best final model: PAPP-A |
|  |  |  | The most recommended validation technique: no internal validation |
| [11] Karagiannis G *et al.* (2011)  Biophysical and biochemical markers and maternal factors | Sensitivity: |  | Study design: retrospective cohort |
|  | 0.34 |  | Population: pregnant women from 11–14 weeks’ gestation, singleton, underwent combined first trimester screening for trisomy 21, no fetal abnormalities, no termination, miscarriage, or fetal death before 24 weeks’ gestation, no preeclampsia, and known pregnancy outcome |
|  | Specificity: |  | Setting: hospital |
|  | 0.95 |  | Outcome definition: event (FGR/SGA, birth weight <5^th^ percentile) and non-event (not FGR/SGA with or without other complications) |
|  | AUROC: *  0.647 |  | Sample size: event (*n*=1536) and non-event (*n*=31,314) |
|  |  |  | Predictors in the best final model: UtA-PI, MAP, NT, PAPP-A, FβhCG, PLGF, PP13, ADAM12, CRL, and maternal factors (age, weight, cigarrette smoker, parity, race, and method of conception) |
|  |  |  | The most recommended validation technique: no internal validation |
| [12] Conserva V *et al.* (2010)  PAPP-A ≤0.4 MoM | Sensitivity: |  | Study design: case-control |
|  | 0.27 |  | Population: pregnant women, singleton, underwent combined first trimester screening for trisomy 21, no chromosomal or fetal abnormalities, no miscarriage, and no gestational diabetes |
|  | Specificity: |  | Setting: biotech lab |
|  | 0.75 |  | Outcome definition: event (FGR/SGA, birth weight <10^th^ percentile) and non-event (not FGR/SGA with or without other complications) |
|  | AUROC: * |  | Sample size: event (*n*=179) and non-event (*n*=1508) |
|  | 0.51 |  | Predictors in the best final model: PAPP-A |
|  |  |  | The most recommended validation technique: no internal validation |
| [13] Cheong M-L *et al.* (2005)  PAPP-A MoM ≤ 5^th^ percentile | Sensitivity: |  | Study design: retrospective cohort |
|  | 0.09 |  | Population: pregnant women from 10–13 weeks’ gestation, singleton, underwent combined first trimester screening for trisomy 21, with complete and correct chart record, no chromosomal or major structural abnormalities, no miscarriage before 24 weeks’ gestation, and no major maternal disease |
|  | Specificity: |  | Setting: hospital |
|  | 0.96 |  | Outcome definition: event (FGR/SGA, birth weight <5^th^ percentile) and non-event (not FGR/SGA with or without other complications) |
|  | AUROC: |  | Sample size: event (*n*=155) and non-event (*n*=2934) |
|  | 0.59 |  | Predictors in the best final model: PAPP-A |
|  |  |  | The most recommended validation technique: no internal validation |
| [14] Cervino-Gomez E *et al.* (2014)  PAPP-A ≤0.4 MoM | Sensitivity:  0.74 |  | Study design: case-control (pairing on PAPP-A group) |
|  |  |  | Population: pregnant women from 11–14 weeks’ gestation, singleton, underwent combined first trimester screening for trisomy 21, maternal age of ≤38 years old, no maternal illness, no chromosomal or fetal abnormalities, and no *in vitro fertilization* |
|  | Specificity: |  | Setting: hospital |
|  | 0.58 |  | Outcome definition: event (FGR/SGA, birth weight <10^th^ percentile) and non-event (not FGR/SGA with or without other complications) |
|  | AUROC: *  0.652 |  | Sample size: event (*n*=95) and non-event (*n*=358) |
|  |  |  | Predictors in the best final model: PAPP-A |
|  |  |  | The most recommended validation technique: no internal validation |
| [15] Carbone JF *et al.* (2012)  (a) PAPP-A ≤ 5^th^ percentile  (b) CRL Z-score < -1  (c) Both (a) and (b)  (d) Either (a) or (b) | Sensitivity: |  | Study design: prospective cohort |
|  | (a) 0.24; (b) 0.12;  (c) 0.03; (d) 0.01 |  | Population: pregnant women from 10–14 weeks’ gestation, singleton, underwent combined first trimester screening for trisomy 21, and no chromosomal and structural abnormalities |
|  | Specificity: |  | Setting: hospital |
|  | (a) 0.86; (b) 0.96;  (c) 0.99; (d) 0.82 |  | Outcome definition: event (FGR/SGA, birth weight <10^th^ percentile) and non-event (not FGR/SGA with or without other complications) |
|  | AUROC: |  | Sample size: event (*n*=185) and non-event (*n*=3144) |
|  | (a) 0.55; (b) 0.54;  (c) 0.51; (d) 0.58 |  | Predictors in the best final model: (a) PAPP-A; (b) CRL; (c) PAPP-A and CRL; (d) PAPP-A and CRL |
|  |  |  | The most recommended validation technique: no internal validation |
| [16] Valiño N *et al.* (2016)  EFW, PlGF, sFlt-1, UtA-PI, UA-PI, MCA-PI | Sensitivity: |  | Study design: prospective cohort |
|  | 0.51 |  | Population: pregnant women from 30–35 weeks’ gestation delivering ≥37 weeks |
|  | Specificity: |  | Setting: hospitals |
|  | 0.9 |  | Outcome definition: event (FGR/SGA, birth weight <10^th^ percentile) and non-event (not FGR/SGA with or without other complications) |
|  | AUROC: |  | Sample size: event (*n*=762) and non-event (*n*=7506) |
|  | 0.829 (0.813 to 0.844) ** |  | Predictors in the best final model: EFW, PlGF, sFlt-1, UtA-PI, UA-PI, and MCA-PI |
|  |  |  | The most recommended validation technique: no internal validation |
| [17] Shlossman P *et al.* (1998)  (a) MCA-PI  (b) ICA-PI  (c) RA-PI  (d) AO-PI  (e) Index  (f) DF | Sensitivity: |  | Study design: NA |
|  | (a) 0.69; (b) 0.77; (c) 0.53;  (d) 0.52; (e) 0.89; (c) 0.78 |  | Population: pregnant women from 25–40 weeks’ gestation |
|  |  |  | Setting: NA |
|  | Specificity:  (a) 0.74; (b) 0.75; (c) 0.55;  (d) 0.54; (e) 0.78; (c) 0.87 |  | Outcome definition: event (FGR/SGA, birth weight <10^th^ percentile) and non-event (not FGR/SGA with or without other complications) |
|  | AUROC: * |  | Sample size: event (*n*=NA) and non-event (*n*=NA) with total *n*=70 |
|  | (a) 0.712; (b) 0.762;  (c) 0.541; (d) 0.527;  (e) 0.837; (f) 0.827 |  | Predictors in the best final model: (a) MCA-PI; (b) ICA-PI; (c) RA-PI; (d) AO-PI; (e) Index=(MCA+ICA)I(RA+AO); and (f) Discriminant function of (a) to (d) |
|  |  |  | The most recommended validation technique: no internal validation |
| [18] Gramellini D *et al.* (1992)  (a) MCA-PI  (b) UA-PI  (c) Cerebral-umbilical ratio | Sensitivity: |  | Study design: case-control |
|  | (a) 0.11; (b) 0.4; (c) 0.4 |  | Population: pregnant women from 30–41 weeks’ gestation |
|  |  |  | Setting: hospital |
|  | Specificity:  (a) 0.98; (b) 0.91; (c) 1.0 |  | Outcome definition: event (FGR/SGA, birth weight <10^th^ percentile) and non-event (uncomplicated pregnancies delivering spontaneously 37–41 weeks) |
|  | AUROC: * |  | Sample size: event (*n*=45) and non-event (*n*=45) |
|  | (a) 0.544; (b) 0.656; (c) 0.7 |  | Predictors in the best final model: (a) MCA-PI; (b) UA-PI; (c) Cerebral-umbilical ratio |
|  |  |  | The most recommended validation technique: no internal validation |
| [19] Gramellini D *et al.* (1990)  (a) MCA S/D  (b) UA S/D  (c) UpA S/D | Sensitivity: |  | Study design: prospective cohort |
|  | (a) 0.45; (b) 0.5; (c) 0.25 |  | Population: pregnant women from 30–40 weeks’ gestation |
|  |  |  | Setting: hospital |
|  | Specificity:  (a) 0.8; (b) 0.83; (c) 0.93 |  | Outcome definition: event (FGR/SGA, birth weight NA percentile) and non-event (not FGR/SGA with or without other complications) |
|  | AUROC: * |  | Sample size: event (*n*=20) and non-event (*n*=30) |
|  | (a) 0.625; (b) 0.667;  (c) 0.592 |  | Predictors in the best final model: (a) MCA S/D; (b) UA S/D; (c) UpA S/D |
|  |  |  | The most recommended validation technique: no internal validation |
| [20] Echizenya N *et al.* (1989)  (a) MCA-RI  (b) UA-RI  (c) UtA-RI | Sensitivity: |  | Study design: case-control |
|  | (a) 0.48; (b) 0.47; (c) 0.29 |  | Population: pregnant women from >17 and >30 weeks’ gestation respectively for cases and controls |
|  |  |  | Setting: hospital |
|  | Specificity:  (a) 0.94; (b) 0.93; (c) 0.93 |  | Outcome definition: event (FGR/SGA, birth weight 10^th^ percentile) and non-event (uncomplicated pregnancies) |
|  | AUROC: * |  | Sample size: event (*n*=30) and non-event (*n*=59) |
|  | (a) 0.71; (b) 0.7; (c) 0.61 |  | Predictors in the best final model: (a) MCA-RI; (b) UA-RI; (c) UtA-RI |
|  |  |  | The most recommended validation technique: no internal validation |
| [21] Bednarek M *et al.* (2004)  Subcortical MCA-PI | Sensitivity: |  | Study design: case-control |
|  | 0.6 |  | Population: pregnant women from 26–42 weeks’ gestation |
|  |  |  | Setting: hospital |
|  | Specificity:  0.95 |  | Outcome definition: event (FGR/SGA, birth weight 10^th^ percentile) and non-event (uncomplicated pregnancies) |
|  | AUROC: * |  | Sample size: event (*n*=64) and non-event (*n*=98) |
|  | 0.77 |  | Predictors in the best final model: Subcortical MCA-PI |
|  |  |  | The most recommended validation technique: no internal validation |
| [22] Bano S *et al.* (2010)  (a) MCA-PI  (b) UA-PI  (c) Cerebral-umbilical ratio | Sensitivity: |  | Study design: case-control |
|  | (a) 0.09; (b) 0.47; (c) 0.44 |  | Population: pregnant women from 30–41 weeks’ gestation |
|  |  |  | Setting: hospital |
|  | Specificity:  (a) 1.0; (b) 0.93; (c) 1.0 |  | Outcome definition: event (FGR/SGA, birth weight <10^th^ percentile) and non-event (not FGR/SGA with or without other complications) |
|  | AUROC: * |  | Sample size: event (*n*=45) and non-event (*n*=45) |
|  | (a) 0.54; (b) 0.7; (c) 0.72 |  | Predictors in the best final model: (a) MCA-PI; (b) UA-PI; (c) Cerebral-umbilical ratio |
|  |  |  | The most recommended validation technique: no internal validation |
| [23] Alataş C *et al.* (1996)  MCA-PI | Sensitivity: |  | Study design: case-control |
|  | 0.32 |  | Population: pregnant women from 24 weeks’ gestation with high-risk pregnancies |
|  |  |  | Setting: hospital |
|  | Specificity:  0.96 |  | Outcome definition: event (FGR/SGA, birth weight <10^th^ percentile) and non-event (not FGR/SGA with other complications) |
|  | AUROC: *  0.64 |  | Sample size: event (*n*=NA) and non-event (*n*=NA) with total *n*=70 |
|  |  |  | Predictors in the best final model: MCA-PI |
|  |  |  | The most recommended validation technique: no internal validation |
| [24] Poon LC *et al.* (2011)  Biophysical and biochemical markers and maternal factors | Sensitivity: |  | Study design: prospective cohort |
|  | 0.252 |  | Population: pregnant women from 11–14 weeks’ gestation, singleton, underwent combined first trimester screening for trisomy 21, with complete outcome data, no chromosomal or major structural abnormalities, no miscarriage or fetal death before 24 weeks’ gestation, no termination for social reasons, and no preeclampsia |
|  |  |  | Setting: hospital |
|  | Specificity:  0.95 |  | Outcome definition: event (FGR/SGA, birth weight <5^th^ percentile) and non-event (uncomplicated pregnancies) |
|  | AUROC:  0.747 (0.735 to 0.76)** |  | Sample size: event (*n*=1536) and non-event (*n*=31,314) |
|  |  |  | Predictors in the best final model: NT, PAPP-A, FβhCG, and maternal factors (age, weight, height, cigarette smoker, parity, race, method of conception, medical history) |
|  |  |  | The most recommended validation technique: no internal validation |
| [25] Plasencia W *et al.* (2007)  Log10 UtA-PI MoM and maternal factors | Sensitivity: |  | Study design: prospective cohort |
|  | 0.246 |  | Population: pregnant women from 11–14 weeks’ gestation, singleton, underwent combined first trimester screening for trisomy 21, with complete data, no chromosomal or major structural abnormalities, no miscarriage or fetal death before 24 weeks’ gestation, and no termination for social reasons |
|  |  |  | Setting: hospital |
|  | Specificity:  0.9 |  | Outcome definition: event (FGR/SGA, birth weight <10^th^ percentile) and non-event (not FGR/SGA with other complications) |
|  | AUROC: |  | Sample size: event (*n*=760) and non-event (*n*=5255) |
|  | 0.661 (0.648 to 0.673)** |  | Predictors in the best final model: UtA-PI and maternal factors (age, BMI, CRL, cigarrette smoker, alcohol drinker, drug abuse, parity, ethnicity, family history of preeclampsia,method of conception, medical history, medication during pregnancy) |
|  |  |  | The most recommended validation technique: no internal validation |
| [26] Onwudiwe N *et al.* (2008)  Log10 UtA-PI MoM | Sensitivity: |  | Study design: prospective cohort |
|  | 0.223 (0.181 to 0.269) ** |  | Population: pregnant women from 22–24 weeks’ gestation, singleton, underwent combined first trimester screening for trisomy 21, with complete outcome data, no fetal abnormalities, and no miscarriage or fetal death before 24 weeks’ gestation |
|  |  |  | Setting: hospital |
|  | Specificity:  0.9 |  | Outcome definition: event (FGR/SGA, birth weight <10^th^ percentile) and non-event (not FGR/SGA with other complications) |
|  | AUROC: |  | Sample size: event (*n*=366) and non-event (*n*=2981) |
|  | 0.609 (0.591 to 0.626)** |  | Predictors in the best final model: UtA-PI |
|  |  |  | The most recommended validation technique: no internal validation |
| [27] De Paco C *et al.* (2008)  Log10 maternal cardiac output MoM and maternal factors | Sensitivity: |  | Study design: prospective cohort |
|  | 0.239 (0.203 to 0.277) ** |  | Population: pregnant women from 11–14 weeks’ gestation, singleton, underwent combined first trimester screening for trisomy 21, with complete outcome data, no fetal abnormalities, no major defects, no miscarriage or fetal death before 24 weeks’ gestation, and no termination for social reasons |
|  |  |  | Setting: hospital |
|  | Specificity:  0.9 |  | Outcome definition: event (FGR/SGA, birth weight <10^th^ percentile) and non-event (not FGR/SGA with or without other complications) |
|  | AUROC: |  | Sample size: event (*n*=532) and non-event (*n*=3761) |
|  | 0.629 (0.602 to 0.655)** |  | Predictors in the best final model: maternal cardiac output and maternal factors (age, weight, height, CRL, cigarrette smoker, alcohol drinker, drug abuse, parity, ethnicity, family history of preeclampsia,method of conception, medical history, medication during pregnancy) |
|  |  |  | The most recommended validation technique: no internal validation |
| * Inferred from sensitivity and specificity for AUROC; or ROC for sensitivity and specificity  ** 95% confidence interval (CI)  ADAM12, A-disintegrin and metalloprotease-12; AO, intra-abdominal aorta; AUROC, area under receiver operating characteristics curve; BMI, body mass index; CRL, crown-rump length (fetus); DF, discriminant function; EFW, estimated fetal weight; FGR, fetal growth restriction; FβhCG, free β-subunit human choriogonadotropin; ICA, internal carotid artery; MAP, mean arterial pressure; MCA, middle cerebral artery; PI, pulsatility index; NA, not available; NT, nuchal translucency thickness (fetus); PAPP-A, pregnancy associated plasma protein-A; PlGF, placental growth factor; PP13, placental protein 13; RA, renal artery; RI, resistance index; S/D, systolic/diastolic ratio; sFLT-1, soluble fms-like tyrosinase-1; SGA, small-for-gestational age; UA, umbilical artery; UpA, utero-placental artery; UtA, uterine artery. | | | |

**References**

1.	Yaron Y, Heifetz S, Ochshorn Y, Lehavi O, Orr-Urtreger A. Decreased first trimester PAPP-A is a predictor of adverse pregnancy outcome. Prenat Diagn 2002 Sep;22(9):778-782. DOI: 10.1002/pd.407

2.	Tul N, Pusenjak S, Osredkar J, Spencer K, Novak-Antolic Z. Predicting complications of pregnancy with first-trimester maternal serum free-betahCG, PAPP-A and inhibin-A. Prenat Diagn 2003 Dec 15;23(12):990-996. DOI: 10.1002/pd.735

3.	Spencer K, Yu CK, Cowans NJ, Otigbah C, Nicolaides KH. Prediction of pregnancy complications by first-trimester maternal serum PAPP-A and free beta-hCG and with second-trimester uterine artery Doppler. Prenat Diagn 2005 Oct;25(10):949-953. DOI: 10.1002/pd.1251

4.	Smith GC, Stenhouse EJ, Crossley JA, Aitken DA, Cameron AD, Connor JM. Early pregnancy levels of pregnancy-associated plasma protein a and the risk of intrauterine growth restriction, premature birth, preeclampsia, and stillbirth. J Clin Endocrinol Metab 2002 Apr;87(4):1762-1767. DOI: 10.1210/jcem.87.4.8430

5.	Radoi VM, Bohiltea LC. Pregnancy-Associated Plasma Protein A and Pregnancy Outcomes. gineco ro maternal fetal medicine 2009;5(1). DOI:

6.	Pilalis A, Souka AP, Antsaklis P, Daskalakis G, Papantoniou N, Mesogitis S, Antsaklis A. Screening for pre-eclampsia and fetal growth restriction by uterine artery Doppler and PAPP-A at 11-14 weeks' gestation. Ultrasound Obstet Gynecol 2007 Feb;29(2):135-140. DOI: 10.1002/uog.3881

7.	Ong CY, Liao AW, Spencer K, Munim S, Nicolaides KH. First trimester maternal serum free beta human chorionic gonadotrophin and pregnancy associated plasma protein A as predictors of pregnancy complications. Bjog 2000 Oct;107(10):1265-1270. DOI: 10.1111/j.1471-0528.2000.tb11618.x

8.	Leung TY, Sahota DS, Chan LW, Law LW, Fung TY, Leung TN, Lau TK. Prediction of birth weight by fetal crown-rump length and maternal serum levels of pregnancy-associated plasma protein-A in the first trimester. Ultrasound Obstet Gynecol 2008 Jan;31(1):10-14. DOI: 10.1002/uog.5206

9.	Krantz D, Goetzl L, Simpson JL, Thom E, Zachary J, Hallahan TW, Silver R, Pergament E, Platt LD, Filkins K, Johnson A, Mahoney M, Hogge WA, Wilson RD, Mohide P, Hershey D, Wapner R. Association of extreme first-trimester free human chorionic gonadotropin-beta, pregnancy-associated plasma protein A, and nuchal translucency with intrauterine growth restriction and other adverse pregnancy outcomes. Am J Obstet Gynecol 2004 Oct;191(4):1452-1458. DOI: 10.1016/j.ajog.2004.05.068

10.	Kavak ZN, Basgul A, Elter K, Uygur M, Gokaslan H. The efficacy of first-trimester PAPP-A and free beta hCG levels for predicting adverse pregnancy outcome. J Perinat Med 2006;34(2):145-148. DOI: 10.1515/jpm.2006.026

11.	Karagiannis G, Akolekar R, Sarquis R, Wright D, Nicolaides KH. Prediction of small-for-gestation neonates from biophysical and biochemical markers at 11-13 weeks. Fetal Diagn Ther 2011;29(2):148-154. DOI: 10.1159/000321694

12.	Conserva V, Signaroldi M, Mastroianni C, Stampalija T, Ghisoni L, Ferrazzi E. Distinction between fetal growth restriction and small for gestational age newborn weight enhances the prognostic value of low PAPP-A in the first trimester. Prenat Diagn 2010 Oct;30(10):1007-1009. DOI: 10.1002/pd.2579

13.	Cheong M-L, She B-Q, Tsai M-S, Chen S-C, Lee F-K. Can First-Trimester Maternal Serum Level Of Pregnancy-associated Plasma Protein-A Predict Subsequent Fetal Growth Restriction? Taiwanese Journal of Obstetrics and Gynecology 2005 2005/06/01/;44(2):148-152. DOI: <https://doi.org/10.1016/S1028-4559(09)60127-3>

14.	Cervino-Gomez E, Gonzalez-Rodriguez L, Cernadas-Pires S, Gonzalez-Boubeta R, Lopez Ramon y Cajal CN. Association of first trimester PAPP-A with small for gestational age infant and other adverse pregnancy outcomes. J Matern Fetal Neonatal Med 2014;27(S1):138. DOI:

15.	Carbone JF, Tuuli MG, Bradshaw R, Liebsch J, Odibo AO. Efficiency of first-trimester growth restriction and low pregnancy-associated plasma protein-A in predicting small for gestational age at delivery. Prenat Diagn 2012 Aug;32(8):724-729. DOI: 10.1002/pd.3891

16.	Valiño N, Giunta G, Gallo DM, Akolekar R, Nicolaides KH. Biophysical and biochemical markers at 30-34 weeks' gestation in the prediction of adverse perinatal outcome. Ultrasound Obstet Gynecol 2016 Feb;47(2):194-202. DOI: 10.1002/uog.14928

17.	Shlossman P, Scisione A, Manley J, Colmorgen G, Weiner S. Doppler assessment of the intrafetal vasculature in the identification of intrauterine growth retardation. Which vessel is ’best’ or is a combination better? American Journal of Obstetrics & Gynecology 1998;178(S1):S88. DOI:

18.	Gramellini D, Folli MC, Raboni S, Vadora E, Merialdi A. Cerebral-umbilical Doppler ratio as a predictor of adverse perinatal outcome. Obstet Gynecol 1992 Mar;79(3):416-420. DOI: 10.1097/00006250-199203000-00018

19.	Gramellini D, Folli M, Sacchini C, Sterbini M, Lombardo A, Merialdi A. Fetal and maternal velocimetry in high risk pregnancies for the assessment of adverse perinatal outcome. Echocardiography 1990;7(5):597-601. DOI:

20.	Echizenya N, Kagiya A, Tachizaki T, Saito Y. Significance of velocimetry as a monitor of fetal assessment and management. Fetal Ther 1989;4(4):188-194. DOI: 10.1159/000263450

21.	Bednarek M, Dubiel M, Brêborowicz GH. P05.18: Doppler velocimetry in M1 and M2 segments of middle cerebral artery in pregnancies complicated by intrauterine growth restriction. Ultrasound in Obstetrics & Gynecology 2004;24(3):300-301. DOI: <https://doi.org/10.1002/uog.1423>

22.	Bano S, Chaudhary V, Pande S, Mehta V, Sharma A. Color doppler evaluation of cerebral-umbilical pulsatility ratio and its usefulness in the diagnosis of intrauterine growth retardation and prediction of adverse perinatal outcome. Indian J Radiol Imaging 2010 Feb;20(1):20-25. PMID: PMC2844742

23.	Alataş C, Aksoy E, Akarsu C, Yakin K, Bahçeci M. Prediction of perinatal outcome by middle cerebral artery Doppler velocimetry. Arch Gynecol Obstet 1996;258(3):141-146. DOI: 10.1007/s004040050115

24.	Poon LC, Karagiannis G, Staboulidou I, Shafiei A, Nicolaides KH. Reference range of birth weight with gestation and first-trimester prediction of small-for-gestation neonates. Prenat Diagn 2011 Jan;31(1):58-65. DOI: 10.1002/pd.2520

25.	Plasencia W, Maiz N, Bonino S, Kaihura C, Nicolaides KH. Uterine artery Doppler at 11 + 0 to 13 + 6 weeks in the prediction of pre-eclampsia. Ultrasound Obstet Gynecol 2007 Oct;30(5):742-749. DOI: 10.1002/uog.5157

26.	Onwudiwe N, Yu CK, Poon LC, Spiliopoulos I, Nicolaides KH. Prediction of pre-eclampsia by a combination of maternal history, uterine artery Doppler and mean arterial pressure. Ultrasound Obstet Gynecol 2008 Dec;32(7):877-883. DOI: 10.1002/uog.6124

27.	De Paco C, Kametas N, Rencoret G, Strobl I, Nicolaides KH. Maternal cardiac output between 11 and 13 weeks of gestation in the prediction of preeclampsia and small for gestational age. Obstet Gynecol 2008 Feb;111(2 Pt 1):292-300. DOI: 10.1097/01.AOG.0000298622.22494.0c

28.	Morris RK, Bilagi A, Devani P, Kilby MD. Association of serum PAPP-A levels in first trimester with small for gestational age and adverse pregnancy outcomes: systematic review and meta-analysis. Prenat Diagn 2017 Mar;37(3):253-265. DOI: 10.1002/pd.5001

29.	Vollgraff Heidweiller-Schreurs CA, Korevaar DA, Mol BWJ, Bax CJ, de Groot CJM, de Boer MA, Bossuyt PMM. Publication bias may exist among prognostic accuracy studies of middle cerebral artery Doppler ultrasound. J Clin Epidemiol 2019 Dec;116:1-8. DOI: 10.1016/j.jclinepi.2019.07.016

30.	Kleinrouweler CE, Cheong-See FM, Collins GS, Kwee A, Thangaratinam S, Khan KS, Mol BWJ, Pajkrt E, Moons KGM, Schuit E. Prognostic models in obstetrics: Available, but far from applicable. American Journal of Obstetrics and Gynecology 2016;214(1):79-90.e36. DOI: 10.1016/j.ajog.2015.06.013

31.	Bachmann LM, Khan KS, Ogah J, Owen P. Multivariable analysis of tests for the diagnosis of intrauterine growth restriction. Ultrasound Obstet Gynecol 2003 Apr;21(4):370-374. DOI: 10.1002/uog.77

32.	de Caunes F, Alexander GR, Berchel C, Guengant JP, Papiernik E. Anamnestic pregnancy risk assessment. Int J Gynaecol Obstet 1990 Nov;33(3):221-227. DOI: 10.1016/0020-7292(90)90005-6

33.	Doherty DA, James IR, Newnham JP. Estimation of the Doppler ultrasound umbilical maximal waveform envelope: II. Prediction of fetal distress. Ultrasound Med Biol 2002 Oct;28(10):1261-1270. DOI: 10.1016/s0301-5629(02)00574-4

34.	Dzikova E, Adamova G, Dzikov Z, et al. The Doppler changes of fetal circulation in patients with preeclampsia and intrauterine growth retardation. Journal of Maternal-Fetal and Neonatal Medicine 2010;23:434. DOI:

35.	Ropacka-Lesiak M, Korbelak T, Breborowicz G. [Doppler blood flow velocimetry in the middle cerebral artery in uncomplicated pregnancy]. Ginekol Pol 2011 Mar;82(3):185-190. DOI:

36.	Ropacka-Lesiak M, Korbelak T, Świder-Musielak J, Breborowicz G. Cerebroplacental ratio in prediction of adverse perinatal outcome and fetal heart rate disturbances in uncomplicated pregnancy at 40 weeks and beyond. Arch Med Sci 2015 Mar 16;11(1):142-148. PMID: PMC4379368

37.	Subramanian V, Venkat J, Dhanapal M. Which is Superior, Doppler Velocimetry or Non-stress Test or Both in Predicting the Perinatal Outcome of High-Risk Pregnancies. J Obstet Gynaecol India 2016 Oct;66(Suppl 1):149-156. PMID: PMC5016437

38.	Goetzinger KR, Singla A, Gerkowicz S, Dicke JM, Gray DL, Odibo AO. The efficiency of first-trimester serum analytes and maternal characteristics in predicting fetal growth disorders. Am J Obstet Gynecol 2009 Oct;201(4):412.e411-416. DOI: 10.1016/j.ajog.2009.07.016

39.	Jouannic JM, Blondiaux E, Senat MV, Friszer S, Adamsbaum C, Rousseau J, Hornoy P, Letourneau A, de Laveaucoupet J, Lecarpentier E, Rosenblatt J, Quibel T, Mollot M, Ancel PY, Alison M, Goffinet F. Prognostic value of diffusion-weighted magnetic resonance imaging of brain in fetal growth restriction: results of prospective multicenter study. Ultrasound Obstet Gynecol 2020 Dec;56(6):893-900. DOI: 10.1002/uog.21926

40.	Villalaín C, Herraiz I, Quezada MS, Gómez-Arriaga PI, Simón E, Gómez-Montes E, Galindo A. Prognostic value of the aortic isthmus Doppler assessment on late onset fetal growth restriction. J Perinat Med 2019 Feb 25;47(2):212-217. DOI: 10.1515/jpm-2018-0185

41.	Akalin-Sel T, Campbell S. Understanding the pathophysiology of intra-uterine growth retardation: the role of the 'lower limb reflex' in redistribution of blood flow. Eur J Obstet Gynecol Reprod Biol 1992 Sep 23;46(2-3):79-86. DOI: 10.1016/0028-2243(92)90250-3

42.	Bartha JL, Moya EM, Hervías-Vivancos B. Three-dimensional power Doppler analysis of cerebral circulation in normal and growth-restricted fetuses. J Cereb Blood Flow Metab 2009 Sep;29(9):1609-1618. DOI: 10.1038/jcbfm.2009.70

43.	Bellido-González M, Díaz-López M, López-Criado S, Maldonado-Lozano J. Cognitive Functioning and Academic Achievement in Children Aged 6-8 Years, Born at Term After Intrauterine Growth Restriction and Fetal Cerebral Redistribution. J Pediatr Psychol 2017 Apr 1;42(3):345-354. DOI: 10.1093/jpepsy/jsw060

44.	Chang TC, Robson SC, Spencer JA, Gallivan S. Prediction of perinatal morbidity at term in small fetuses: comparison of fetal growth and Doppler ultrasound. Br J Obstet Gynaecol 1994 May;101(5):422-427. DOI: 10.1111/j.1471-0528.1994.tb11916.x

45.	Cruz-Martínez R, Figueras F, Hernandez-Andrade E, Oros D, Gratacos E. Fetal brain Doppler to predict cesarean delivery for nonreassuring fetal status in term small-for-gestational-age fetuses. Obstet Gynecol 2011 Mar;117(3):618-626. DOI: 10.1097/AOG.0b013e31820b0884

46.	Cruz-Martinez R, Tenorio V, Padilla N, Crispi F, Figueras F, Gratacos E. Risk of ultrasound-detected neonatal brain abnormalities in intrauterine growth-restricted fetuses born between 28 and 34 weeks' gestation: relationship with gestational age at birth and fetal Doppler parameters. Ultrasound Obstet Gynecol 2015 Oct;46(4):452-459. DOI: 10.1002/uog.14920

47.	Herskovitz R, Kingdom J, Rodeck C. Cerebral blood flow redistribution in small term fetuses with normal umbilical artery blood flow. American journal of obstetrics and gynecology 1999;180(S1). DOI:

48.	Illa M, Iraola A, Meler E, Eixarch E, Figueras F, Gratacós E. OP14.09: Perinatal and neurological outcomes in low birth fetuses with pathologic doppler study in middle cerebral artery. Ultrasound in Obstetrics & Gynecology 2007 10/01;30:503-504. DOI: 10.1002/uog.4552

49.	Khalil A, Morales Rosello J, Nath M, Bhide A, Papageorghiou AT, Thilaganathan B. OP10.10: Longitudinal change in the cerebroplacental ratio in pregnancies with small‐for‐gestational‐age fetuses complicated by stillbirth and perinatal death. Ultrasound in Obstetrics & Gynecology 2017 09/01;50:80-80. DOI: 10.1002/uog.17788

50.	Murata S, Nakata M, Sumie M, Sugino N. OC29.03: The Doppler cerebroplacental ratio predicts risk of non-reassuring fetal status for fetal growth restriction in term pregnancy. Ultrasound in Obstetrics & Gynecology 2009;34(S1):56-56. DOI: <https://doi.org/10.1002/uog.6629>

51.	Oros D, Figueras F, Cruz-Martinez R, Padilla N, Meler E, Hernandez-Andrade E, Gratacos E. Middle versus anterior cerebral artery Doppler for the prediction of perinatal outcome and neonatal neurobehavior in term small-for-gestational-age fetuses with normal umbilical artery Doppler. Ultrasound Obstet Gynecol 2010 Apr;35(4):456-461. DOI: 10.1002/uog.7588

52.	Sekizuka N. Combined examination of middle cerebral artery and umbilical artery flow velocity waveforms in growth-retarded fetuses. Asia Oceania J Obstet Gynaecol 1993 Mar;19(1):13-19. DOI: 10.1111/j.1447-0756.1993.tb00341.x

53.	Somprasit C, Chanthasenanont A, Nanthakomon T, Pongrojpaw D. P01.05: The value of middle cerebral artery-umbilical artery pulsatility index ratio in prediction of severe fetal growth restriction. Ultrasound in obstetrics & gynecology : the official journal of the International Society of Ultrasound in Obstetrics and Gynecology 2010 10/01;36:169. DOI: 10.1002/uog.8307

54.	Vázquez-Sarandeses A, Gómez-Montes E, Herraiz I, Gómez-Arriaga P, Quezada M, Galindo A. Validation of a predictive risk model for adverse perinatal outcome in late-onset small for gestational age fetuses. J Matern Fetal Neonatal Med 2016;29(S1):17. DOI:

55.	Zohav E, Elasbach A, Sofer H, Hadad A, Segal O, Anteby E. P29.07: The role of third trimester cerebroplacental ratio in the prediction of the mode of delivery in singleton AGA and SGA fetuses. Ultrasound in Obstetrics & Gynecology 2016;48(S1):265-. DOI:

56.	Alici Davutoglu E, Ozel A, Oztunc F, Madazli R. Modified myocardial performance index and its prognostic significance for adverse perinatal outcome in early and late onset fetal growth restriction. J Matern Fetal Neonatal Med 2020 Jan;33(2):277-282. DOI: 10.1080/14767058.2018.1489534

57.	Bilagi A, Burke DL, Riley RD, Mills I, Kilby MD, Morris RK. Association of maternal serum PAPP-A levels, nuchal translucency and crown-rump length in first trimester with adverse pregnancy outcomes: retrospective cohort study. PRENATAL DIAGNOSIS 2017 JUL;37(7):705-711. DOI: 10.1002/pd.5069

58.	Gong S, Gaccioli F, Dopierala J, Sovio U, Cook E, Volders PJ, Martens L, Kirk PDW, Richardson S, Smith GCS, Charnock-Jones DS. The RNA landscape of the human placenta in health and disease. Nat Commun 2021 May 11;12(1):2639. PMID: PMC8113443 outside the submitted work and non-financial support from Roche Diagnostics Ltd, outside the submitted work; G.C.S.S. reports grants and personal fees from GlaxoSmithKline Research and Development Limited, personal fees and non-financial support from Roche Diagnostics Ltd, outside the submitted work; D.S.C-J. and G.C.S.S. report grants from Sera Prognostics Inc, non-financial support from Illumina Inc, outside the submitted work. J.D. reports being an employee of GlaxoSmithKline Research and Development Limited, outside the submitted work. S.G., F.G., U.S., E.C., P-J.V., L.M., P.D.W.K., and S.R. have nothing to disclose.

59.	Kim SY, Lee SM, Sung SJ, Han SJ, Kim BJ, Park CW, Park JS, Jun JK. Red cell distribution width as a potential prognostic biomarker in fetal growth restriction. J Matern Fetal Neonatal Med 2021 Mar;34(6):883-888. DOI: 10.1080/14767058.2019.1622665

60.	Perry H, Gutierrez J, Binder J, Thilaganathan B, Khalil A. Maternal arterial stiffness in hypertensive pregnancies with and without small-for-gestational-age neonate. ULTRASOUND IN OBSTETRICS & GYNECOLOGY 2020 JUL;56(1):44-50. DOI: 10.1002/uog.21893

61.	Tramontana A, Dieplinger B, Stangl G, Hafner E, Dieplinger H. First trimester serum afamin concentrations are associated with the development of pre-eclampsia and gestational diabetes mellitus in pregnant women. Clinica Chimica Acta 2018;476:160-166. DOI: 10.1016/j.cca.2017.11.031

62.	Tramontana A, Pablik E, Stangl G, Hartmann B, Dieplinger H, Hafner E. Combination of first trimester serum afamin levels and three-dimensional placental bed vascularization as a possible screening method to detect women at-risk for adverse pregnancy complications like pre-eclampsia and gestational diabetes mellitus in low-risk pregnancies. Placenta 2018 Feb;62:9-15. DOI: 10.1016/j.placenta.2017.12.014

63.	Figueras F, Cruz-Martinez R, Sanz-Cortes M, Arranz A, Illa M, Botet F, Costas-Moragas C, Gratacos E. Neurobehavioral outcomes in preterm, growth-restricted infants with and without prenatal advanced signs of brain-sparing. Ultrasound Obstet Gynecol 2011 Sep;38(3):288-294. DOI: 10.1002/uog.9041

64.	Mari G, Abuhamad AZ, Keller M, Verpairojkit B, Ment L, Copel JA. Is the fetal brain-sparing effect a risk factor for the development of intraventricular hemorrhage in the preterm infant? Ultrasound Obstet Gynecol 1996 Nov;8(5):329-332. DOI: 10.1046/j.1469-0705.1996.08050329.x

65.	Akhavan S, Lak P, Rahimi-Sharbaf F, Mohammadi SR, Shirazi M. Admission Test and Pregnancy Outcome. Iran J Med Sci 2017 Jul;42(4):362-368. PMID: PMC5523043

66.	Albaiges Baiget G, Meler E, Caner N, Rodríguez I, Echevarria M, Serra B. EP16.09: Cerebroplacental ratio at term and risk of adverse perinatal outcome. Ultrasound in Obstetrics & Gynecology 2017;50(S1):334-334. DOI: <https://doi.org/10.1002/uog.18583>

67.	Cameán M, Antolin E, Mateos A, J B. EP15.07: Influence of umbilical artery resistance and fetal weight on fetal vascular circulatory redistibution. Ultrasound in Obstetrics & Gynecology 2016;48(S1):327-. DOI:

68.	Cheema R, Bayoumi MZ, Gudmundsson S. Multivascular Doppler surveillance in high risk pregnancies. J Matern Fetal Neonatal Med 2012 Jul;25(7):970-974. DOI: 10.3109/14767058.2011.602141

69.	Korszun P, Dubiel M, Breborowicz G, Danska A, Gudmundsson S. Fetal superior mesenteric artery blood flow velocimetry in normal and high-risk pregnancy. J Perinat Med 2002;30(3):235-241. DOI: 10.1515/jpm.2002.033

70.	Strigini FA, De Luca G, Lencioni G, Scida P, Giusti G, Genazzani AR. Middle cerebral artery velocimetry: different clinical relevance depending on umbilical velocimetry. Obstet Gynecol 1997 Dec;90(6):953-957. DOI: 10.1016/s0029-7844(97)00482-1

71.	To W, Mok KM. P215: Comparison of two different placental-cerebral ratios in presence of normal umbilical arterial waveform in assessment of fetal growth restriction near term. Ultrasound in Obstetrics & Gynecology 2003;22(S1):128-128. DOI: <https://doi.org/10.1002/uog.681>

72.	Alanwar A, El Nour AA, El Mandooh M, Abdelazim IA, Abbas L, Abbas AM, Abdallah A, Nossair WS, Svetlana S. Prognostic accuracy of cerebroplacental ratio for adverse perinatal outcomes in pregnancies complicated with severe pre-eclampsia; a prospective cohort study. Pregnancy Hypertens 2018 Oct;14:86-89. DOI: 10.1016/j.preghy.2018.08.446

73.	Lim S, Li W, Kemper J, Nguyen A, Mol BW, Reddy M. Biomarkers and the Prediction of Adverse Outcomes in Preeclampsia: A Systematic Review and Meta-analysis. Obstet Gynecol 2021 Jan 1;137(1):72-81. DOI: 10.1097/aog.0000000000004149

74.	Wnuk A, Stangret A, Wątroba M, Płatek AE, Skoda M, Cendrowski K, Sawicki W, Szukiewicz D. Can adipokine visfatin be a novel marker of pregnancy-related disorders in women with obesity? Obes Rev 2020 Jul;21(7):e13022. DOI: 10.1111/obr.13022

75.	Seed PT, Chappell LC, Black MA, Poppe KK, Hwang YC, Kasabov N, McCowan L, Shennan AH, Wu SH, Poston L, North RA. Prediction of preeclampsia and delivery of small for gestational age babies based on a combination of clinical risk factors in high-risk women. Hypertens Pregnancy 2011;30(1):58-73. DOI: 10.3109/10641955.2010.486460

76.	Ozeren M, Dinç H, Ekmen U, Senekayli C, Aydemir V. Umbilical and middle cerebral artery Doppler indices in patients with preeclampsia. Eur J Obstet Gynecol Reprod Biol 1999 Jan;82(1):11-16. DOI: 10.1016/s0301-2115(98)00167-5

77.	Simanaviciute D, Gudmundsson S. Fetal middle cerebral to uterine artery pulsatility index ratios in normal and pre-eclamptic pregnancies. Ultrasound Obstet Gynecol 2006 Nov;28(6):794-801. DOI: 10.1002/uog.3805

78.	Atallah A, Guibaud L, Gaucherand P, Massardier J, des Portes V, Massoud M. Fetal and perinatal outcome associated with small cerebellar diameter based on second- or third-trimester ultrasonography. PRENATAL DIAGNOSIS 2019 JUN;39(7):536-543. DOI: 10.1002/pd.5465

79.	Bhorat I, Pillay M, Reddy T. The clinical prognostic significance of myocardial performance index (MPI) in stable placental-mediated disease. Cardiovasc J Afr 2018 Sep/Oct 23;29(5):310-316. DOI: 10.5830/cvja-2018-036

80.	Hamilton EF, Dyachenko A, Ciampi A, Maurel K, Warrick PA, Garite TJ. Estimating risk of severe neonatal morbidity in preterm births under 32 weeks of gestation. J Matern Fetal Neonatal Med 2020 Jan;33(1):73-80. DOI: 10.1080/14767058.2018.1487395

81.	Ortiz JU, Graupner O, Karge A, Flechsenhar S, Haller B, Ostermayer E, Abel K, Kuschel B, Lobmaier SM. Does gestational age at term play a role in the association between cerebroplacental ratio and operative delivery for intrapartum fetal compromise? ACTA OBSTETRICIA ET GYNECOLOGICA SCANDINAVICA 2021. DOI: 10.1111/aogs.14222

82.	Vollgraff Heidweiller-Schreurs CA, De Boer MA, Heymans MW, Schoonmade LJ, Bossuyt PMM, Mol BWJ, De Groot CJM, Bax CJ. Prognostic accuracy of cerebroplacental ratio and middle cerebral artery Doppler for adverse perinatal outcome: systematic review and meta-analysis. Ultrasound Obstet Gynecol 2018 Mar;51(3):313-322. PMID: PMC5873403

83.	Vollgraff Heidweiller-Schreurs CA, van Osch IR, Heymans MW, Ganzevoort W, Schoonmade LJ, Bax CJ, Mol B, de Groot C, Bossuyt P, de Boer MA. Cerebroplacental ratio in predicting adverse perinatal outcome: a meta-analysis of individual participant data. Bjog 2021 Jan;128(2):226-235. PMID: PMC7818434

84.	Bakalis S, Akolekar R, Gallo DM, Poon LC, Nicolaides KH. Umbilical and fetal middle cerebral artery Doppler at 30-34 weeks' gestation in the prediction of adverse perinatal outcome. Ultrasound Obstet Gynecol 2015 Apr;45(4):409-420. DOI: 10.1002/uog.14822

85.	Bligh LN, Alsolai AA, Greer RM, Kumar S. Cerebroplacental ratio thresholds measured within 2 weeks before birth and risk of Cesarean section for intrapartum fetal compromise and adverse neonatal outcome. Ultrasound Obstet Gynecol 2018 Sep;52(3):340-346. DOI: 10.1002/uog.17542

86.	Chandran R, Serra-Serra V, Sellers SM, Redman CW. Fetal cerebral Doppler in the recognition of fetal compromise. Br J Obstet Gynaecol 1993 Feb;100(2):139-144. DOI: 10.1111/j.1471-0528.1993.tb15209.x

87.	D'Antonio F, Iacovella C, Agarwal N, Thilaganathan B, Bhide A. OP16.04: The value of cerebroplacental ratio for fetal assessment in prolonged pregnancy. Ultrasound in Obstetrics & Gynecology 2011;38(S1):102-102. DOI: <https://doi.org/10.1002/uog.9408>

88.	Gaafar HM, El Wahab HA, Abd El Fatah G, Salah D, Rifai NM. OP29.03: Brain volume and Doppler velocimetry in growth-restricted, small-for- and appropriate-for-gestational age fetuses. Ultrasound in Obstetrics & Gynecology 2016;48(S1):147-147. DOI: <https://doi.org/10.1002/uog.16439>

89.	Khalil AA, Morales-Rosello J, Morlando M, Hannan H, Bhide A, Papageorghiou A, Thilaganathan B. Is fetal cerebroplacental ratio an independent predictor of intrapartum fetal compromise and neonatal unit admission? Am J Obstet Gynecol 2015 Jul;213(1):54.e51-54.e10. DOI: 10.1016/j.ajog.2014.10.024

90.	Maged AM, Abdelhafez A, Al Mostafa W, Elsherbiny W. Fetal middle cerebral and umbilical artery Doppler after 40 weeks gestational age. J Matern Fetal Neonatal Med 2014 Dec;27(18):1880-1885. DOI: 10.3109/14767058.2014.892068

91.	Monaghan C, Binder J, Thilaganathan B, Morales-Rosello J, Khalil A. The role of uteroplacental and fetal Dopplers at term as markers of perinatal death. BJOG 2017;124(S2):7. DOI:

92.	Barrett SL, Bower C, Hadlow NC. Use of the combined first-trimester screen result and low PAPP-A to predict risk of adverse fetal outcomes. Prenat Diagn 2008 Jan;28(1):28-35. DOI: 10.1002/pd.1898

93.	Dane B, Dane C, Kiray M, Cetin A, Koldas M, Erginbas M. Correlation between first-trimester maternal serum markers, second-trimester uterine artery doppler indices and pregnancy outcome. Gynecol Obstet Invest 2010;70(2):126-131. DOI: 10.1159/000303260

94.	Goetzinger KR, Cahill AG, Macones GA, Odibo AO. Association of first-trimester low PAPP-A levels with preterm birth. Prenat Diagn 2010 Apr;30(4):309-313. DOI: 10.1002/pd.2452

95.	Jasim SK, Al-Momen H, Al-Naddawi AM. Prediction of maternal diabetes and adverse neonatal outcome in normotensive pregnancy using serum uric acid. International Journal of Research in Pharmaceutical Sciences 2019;10(4):3563-3569. DOI: 10.26452/ijrps.v10i4.1736

96.	Lagendijk J, Steyerberg EW, Daalderop LA, Been JV, Steegers EAP, Posthumus AG. Validation of a prognostic model for adverse perinatal health outcomes. Sci Rep 2020 Jul 9;10(1):11243. PMID: PMC7347528

97.	Griffin M, Heazell AEP, Chappell LC, Zhao J, Lawlor DA. The ability of late pregnancy maternal tests to predict adverse pregnancy outcomes associated with placental dysfunction (specifically fetal growth restriction and pre-eclampsia): a protocol for a systematic review and meta-analysis of prognostic accuracy studies. Syst Rev 2020 Apr 8;9(1):78. PMID: PMC7140577

98.	Petersen KB. Individual fertility assessment and counselling in women of reproductive age. Dan Med J 2016 Oct;63(10). DOI:

99.	Kholin AM, Khodzhaeva ZS, Gus AI. Pathological placentation and prediction of preeclampsia and intrauterine growth restriction in the first trimester. Akusherstvo i Ginekologiya (Russian Federation) 2018 (5):12-19. DOI: 10.18565/aig.2018.5.12-19

100.	Villalain C, Galindo A, Di Mascio D, Buca D, Morales-Rosello J, Loscalzo G, Giulia Sileo F, Finarelli A, Bertucci E, Facchinetti F, Rizzo G, Brunelli R, Giancotti A, Muzii L, Maria Maruotti G, Carbone L, D'Amico A, Tinari S, Morelli R, Cerra C, Nappi L, Greco P, Liberati M, D'Antonio F, Herraiz I. Diagnostic performance of cerebroplacental and umbilicocerebral ratio in appropriate for gestational age and late growth restricted fetuses attempting vaginal delivery: a multicenter, retrospective study. J Matern Fetal Neonatal Med 2021 Jun 8:1-7. DOI: 10.1080/14767058.2021.1926977

101.	Snidvongs W, Bhongsvej S, Witoonpanich P, Thaitumyanond P, Charoenvidhya D, Wiswasukmongkol V, Tannirandorn Y, Trisukosol D. Intrauterine growth retardation: incidence, screening results, pregnancy outcome. J Med Assoc Thai 1989 Jul;72(7):387-394. DOI:

102.	Baschat A, Kramer W, Reiss I, Gortner LW, U G. Perinatal mortality and its relationship to abnormal arterial and venous flow in growth restricted fetuses. American journal of obstetrics and gynecology 1998;178(S1). DOI:

103.	Baschat A, Reiss I, Gortner L, Welner C, Harman C. Brain sparing does not cause intraventricular hemorrhage. American Journal of Obstetrics & Gynecology 2000;182(S1). DOI:

104.	Bayoumy A, Crauciuc E, Pricop F. [Cerebro-umbilical ratio--high fidelity indicator in ante-partum fetal disorder]. Rev Med Chir Soc Med Nat Iasi 2005 Jul-Sep;109(3):532-536. DOI:

105.	Cavero A ML, De Miguel JR, Mora R, Sancho I, Ceballos C. Predictive capacity of Doppler velocimetry to predict the perinatal outcome in pregnancies with antenatal suspect of intrauterine growth retardation (IUGR). Progresos en Obstetricia y Ginecologia 1996;39(6):415-422. DOI:

106.	Devine P, Bracero L, Lysikiewicz A, Tejani N. Middle cerebral-umbilical Doppler ratio predicts adverse outcome in postdate pregnancies. American Journal of Obstetrics & Gynecology 1993;166(S1). DOI:

107.	Fuchs T, Zalewski J, Zimmer M, Florjański J, Pańczak K. [Useful of cerebral placental ratio in pregnancies complicated by intrauterine growth retardation and it correlation with perinatal outcome]. Ginekol Pol 2000 Apr;71(4):304-310. DOI:

108.	Li X, Zhuang Y, Zhang J. [The evaluation of various fetal blood flow velocity waveforms in predicting perinatal outcomes]. Zhonghua Fu Chan Ke Za Zhi 1995 Jan;30(1):22-26. DOI:

109.	Meyberg R, Hendrik HJ, Ertan AK, Friedrich M, Schmidt W. The clinical significance of antenatal pathological Doppler findings in fetal middle cerebral artery compared to umbilical artery and fetal aorta. Clin Exp Obstet Gynecol 2000;27(2):92-94. DOI:

110.	Ott W. Comparison of the non-stress test and middle cerebral to umbilical artery systolic/diastolic velocity wave form ratio for the prediction of neonatal compromise. Journal of Maternal-Fetal Investigation 1996;6(2):73-76. DOI:

111.	Pál A, Ulrich G, Manfred H. [Prognostic value of the study of the blood flow in the fetal median cerebral artery]. Orv Hetil 1991 Aug 18;132(33):1815-1817. DOI:

112.	Pare E, Koll A, Fouron J. Prognostic value of fetal middle cerebral artery and umbilical artery dopplers in the setting of bilateral abnormal uterine flow. American Journal of Obstetrics & Gynecology 1999;180(S1). DOI:

113.	Sacchini C, Ludovici G, Piantelli G, Amone F, Paita Y, Gramellini D. Clinical findings in gestational hypertension categorized by doppler velocimetry. Journal of Maternal-Fetal Investigation 1995;5(4):230-235. DOI:

114.	Shpati D, Qirko R, Balla F. Prediction by doppler in iugr and preeclampsia for perinatal outcome. Journal of Perinatal Medicine 2013;41. DOI:

115.	Souza A, Scavuzzi A, Noronha-Neto C, Neto M, Amorim M. T5.2 Resistance index of uterine, umbilical and middle cerebral fetal arteries to predict small for gestational age infants in pregnant women with hypertensive syndromes. Cancer Letters - CANCER LETT 2010 10/01;1. DOI: 10.1016/S2210-7789(10)60073-6

116.	Zha C, Li J, Li X. [Pulsatility indexes of fetal middle cerebral artery and umbilical artery for predicting intrauterine fetal growth retardation]. Zhonghua Fu Chan Ke Za Zhi 1996 Jun;31(6):345-347. DOI:

117.	Linsell L, Malouf R, Johnson S, Morris J, Kurinczuk JJ, Marlow N. Prognostic factors for behavioral problems and psychiatric disorders in children born very preterm or very low birth weight: A systematic review. Journal of Developmental and Behavioral Pediatrics 2016;37(1):88-102. DOI: 10.1097/DBP.0000000000000238

118.	Lobmaier SM, Mensing van Charante N, Ferrazzi E, Giussani DA, Shaw CJ, Müller A, Ortiz JU, Ostermayer E, Haller B, Prefumo F, Frusca T, Hecher K, Arabin B, Thilaganathan B, Papageorghiou AT, Bhide A, Martinelli P, Duvekot JJ, van Eyck J, Visser GH, Schmidt G, Ganzevoort W, Lees CC, Schneider KT. Phase-rectified signal averaging method to predict perinatal outcome in infants with very preterm fetal growth restriction- a secondary analysis of TRUFFLE-trial. Am J Obstet Gynecol 2016 Nov;215(5):630.e631-630.e637. DOI: 10.1016/j.ajog.2016.06.024

119.	Monteith C, Flood K, Mullers S, Unterscheider J, Breathnach F, Daly S, Geary MP, Kennelly MM, McAuliffe FM, O'Donoghue K, Hunter A, Morrison JJ, Burke G, Dicker P, Tully EC, Malone FD. Evaluation of normalization of cerebro-placental ratio as a potential predictor for adverse outcome in SGA fetuses. Am J Obstet Gynecol 2017 Mar;216(3):285.e281-285.e286. DOI: 10.1016/j.ajog.2016.11.1008

120.	Pels A, Mensing van Charante NA, Vollgraff Heidweiller-Schreurs CA, Limpens J, Wolf H, de Boer MA, Ganzevoort W. The prognostic accuracy of short term variation of fetal heart rate in early-onset fetal growth restriction: A systematic review. Eur J Obstet Gynecol Reprod Biol 2019 Mar;234:179-184. DOI: 10.1016/j.ejogrb.2019.01.005

121.	Villalain C, Quezada MS, Gomez-Arriaga P, Simon E, Gomez-Montes E, Galindo A, Herraiz I. Prognostic Factors of Successful Cervical Ripening and Labor Induction in Late-Onset Fetal Growth Restriction. FETAL DIAGNOSIS AND THERAPY 2020 JUL;47(7):536-544. DOI: 10.1159/000503390

122.	Yoshida A, Umehara N, Sasahara J, Ozawa K, Ichizuka K, Tanaka K, Tanemoto T, Ishikawa H, Murakoshi T, Kiyoshi K, Oba MS, Ishii K, Sago H. Prenatal risk stratification of severe small-for-gestational-Age infants: A Japanese multicenter study. Journal of Maternal-Fetal and Neonatal Medicine 2016;29(8):1353-1357. DOI: 10.3109/14767058.2015.1049147

123.	Arbeille P, Perrotin F, Salihagic A, Sthale H, Lansac J, Platt LD. Fetal Doppler Hypoxic index for the prediction of abnormal fetal heart rate at delivery in chronic fetal distress. Eur J Obstet Gynecol Reprod Biol 2005 Aug 1;121(2):171-177. DOI: 10.1016/j.ejogrb.2004.11.032

124.	Arduini D, Rizzo G. Prediction of fetal outcome in small for gestational age fetuses: comparison of Doppler measurements obtained from different fetal vessels. J Perinat Med 1992;20(1):29-38. DOI: 10.1515/jpme.1992.20.1.29

125.	Babic I, Alameri S, Tulbah M, Moretti F, Kurdi W. P08.08: Cerebroplacental ratio and its association with adverse perinatal outcomes in early and late onset intrauterine growth restriction (IUGR). Ultrasound in Obstetrics & Gynecology 2015;46(S1):149-149. DOI: <https://doi.org/10.1002/uog.15394>

126.	Bahado-Singh RO, Kovanci E, Jeffres A, Oz U, Deren O, Copel J, Mari G. The Doppler cerebroplacental ratio and perinatal outcome in intrauterine growth restriction. Am J Obstet Gynecol 1999 Mar;180(3 Pt 1):750-756. DOI: 10.1016/s0002-9378(99)70283-8

127.	Baptista E, Domingues AP, Marta E, Moura P. P08.16: Perinatal outcome of lat… onset smalŒ fo’ gestationaŒ age foetuses with abnormal middle cerebral artery Doppler fluxometry: preliminary results. Ultrasound in Obstetrics & Gynecology 2012;40. DOI:

128.	Baschat AA, Cosmi E, Bilardo CM, Wolf H, Berg C, Rigano S, Germer U, Moyano D, Turan S, Hartung J, Bhide A, Müller T, Bower S, Nicolaides KH, Thilaganathan B, Gembruch U, Ferrazzi E, Hecher K, Galan HL, Harman CR. Predictors of neonatal outcome in early-onset placental dysfunction. Obstet Gynecol 2007 Feb;109(2 Pt 1):253-261. DOI: 10.1097/01.Aog.0000253215.79121.75

129.	Baschat AA, Gembruch U, Viscardi RM, Gortner L, Harman CR. Antenatal prediction of intraventricular hemorrhage in fetal growth restriction: what is the role of Doppler? Ultrasound Obstet Gynecol 2002 Apr;19(4):334-339. DOI: 10.1046/j.1469-0705.2002.00661.x

130.	Bilinski R, Guirguis G, JR A. OC15.04: The Tei index as a sonographic tool for predicting fetal acidosis in intrauterine growth restricted fetuses with placental insufficiency. Ultrasound in Obstetrics & Gynecology 2015;46(S1):32-. DOI:

131.	Bozzetti V, Paterlini G, Gazzolo D, Van Bel F, Visser GH, Roncaglia N, Tagliabue PE. Monitoring Doppler patterns and clinical parameters may predict feeding tolerance in intrauterine growth-restricted infants. Acta Paediatr 2013 Nov;102(11):e519-523. DOI: 10.1111/apa.12380

132.	Chalubinski KM, Repa A, Stammler-Safar M, Ott J. Impact of Doppler sonography on intrauterine management and neonatal outcome in preterm fetuses with intrauterine growth restriction. Ultrasound Obstet Gynecol 2012 Mar;39(3):293-298. DOI: 10.1002/uog.9039

133.	Constantinescu S, Denes M, Vladareanu R. Intrauterine growth restriction: Perinatal assessment in predicting the offspring neurologic impairment. A 2 years prospective study. Ginecoeu 2013;9(1):35-40. DOI:

134.	Crimmins S, Desai A, Block-Abraham D, Berg C, Gembruch U, Baschat AA. A comparison of Doppler and biophysical findings between liveborn and stillborn growth-restricted fetuses. Am J Obstet Gynecol 2014 Dec;211(6):669.e661-610. DOI: 10.1016/j.ajog.2014.06.022

135.	Del Río M, Martínez JM, Figueras F, Bennasar M, Olivella A, Palacio M, Coll O, Puerto B, Gratacós E. Doppler assessment of the aortic isthmus and perinatal outcome in preterm fetuses with severe intrauterine growth restriction. Ultrasound Obstet Gynecol 2008 Jan;31(1):41-47. DOI: 10.1002/uog.5237

136.	Dubiel M, Breborowicz GH, Gudmundsson S. Evaluation of fetal circulation redistribution in pregnancies with absent or reversed diastolic flow in the umbilical artery. Early Hum Dev 2003 Apr;71(2):149-156. DOI: 10.1016/s0378-3782(03)00006-9

137.	Dubiel M, Korszun P, Ropacka M, Breborowicz G. [Fetal brain blood flow velocity monitoring in high risk pregnancies]. Ginekol Pol 2003 Oct;74(10):1076-1082. DOI:

138.	Eixarch E, Meler E, Iraola A, Illa M, Crispi F, Hernandez-Andrade E, Gratacos E, Figueras F. Neurodevelopmental outcome in 2-year-old infants who were small-for-gestational age term fetuses with cerebral blood flow redistribution. Ultrasound Obstet Gynecol 2008 Dec;32(7):894-899. DOI: 10.1002/uog.6249

139.	Ekholm E, Maunu J, Palo P, Rikalainen H, Lapinleimu H, Inki P, Haataja L, Lehtonen L. P05.20: The association of antenatal blood flow with neonatal brain damage in very low birth‐weight infants. Ultrasound in Obstetrics & Gynecology 2004 08/01;24:301-301. DOI: 10.1002/uog.1425

140.	Fardiazar Z, Atashkhouei S, Yosefzad Y, Goldust M, Torab R. Comparison of fetal middle cerebral arteries, umbilical and uterin artery color Doppler ultrasound with blood gas analysis in pregnancy complicated by IUGR. Iran J Reprod Med 2013 Jan;11(1):47-51. PMID: PMC3941376

141.	Figueras F, Savchev S, Triunfo S, Crovetto F, Gratacos E. An integrated model with classification criteria to predict small-for-gestational-age fetuses at risk of adverse perinatal outcome. Ultrasound Obstet Gynecol 2015 Mar;45(3):279-285. DOI: 10.1002/uog.14714

142.	Figueras Retuerta F BM, Eixarch E, et al. Arterial, venous and intracardiac parameters in growth-restricted fetuses: Associations with adverse perinatal outcome. Ultrasound Review of Obstetrics and Gynecology 2004;4(3):179-185. DOI:

143.	Flood K, Unterscheider J, Daly S, Geary MP, Kennelly MM, McAuliffe FM, O'Donoghue K, Hunter A, Morrison JJ, Burke G, Dicker P, Tully EC, Malone FD. The role of brain sparing in the prediction of adverse outcomes in intrauterine growth restriction: results of the multicenter PORTO Study. Am J Obstet Gynecol 2014 Sep;211(3):288.e281-285. DOI: 10.1016/j.ajog.2014.05.008

144.	Fong KW, Ohlsson A, Hannah ME, Grisaru S, Kingdom J, Cohen H, Ryan M, Windrim R, Foster G, Amankwah K. Prediction of perinatal outcome in fetuses suspected to have intrauterine growth restriction: Doppler US study of fetal cerebral, renal, and umbilical arteries. Radiology 1999 Dec;213(3):681-689. DOI: 10.1148/radiology.213.3.r99dc08681

145.	Fratelli N, Orabona R, Cavalli C, et al. P29.02: Umbilical cerebral ratio for detection of poor neonatal acid–base status in small-for-gestational age late preterm fetuses. Ultrasound in Obstetrics & Gynecology 2016;48(S1):263-264. DOI:

146.	Fu J, Olofsson P. Relations between fetal brain-sparing circulation, oxytocin challenge test, mode of delivery and fetal outcome in growth-restricted term fetuses. Acta Obstet Gynecol Scand 2011 Mar;90(3):227-230. DOI: 10.1111/j.1600-0412.2010.01042.x

147.	Fuchs F, Bentaleb J, Taillefer C, Malhamé I, Wavrant S, Dubé J, Fouron J, Audibert F. OP29.09: Umbilical and middle cerebral artery Doppler to predict adverse outcome in a cohort of fetuses with intrauterine growth restriction. Ultrasound in Obstetrics & Gynecology 2014 09/01;44:134-135. DOI: 10.1002/uog.13925

148.	Garcia-Simon R, Figueras F, Savchev S, Fabre E, Gratacos E, Oros D. Cervical condition and fetal cerebral Doppler as determinants of adverse perinatal outcome after labor induction for late-onset small-for-gestational-age fetuses. Ultrasound Obstet Gynecol 2015 Dec;46(6):713-717. DOI: 10.1002/uog.14807

149.	Goetzinger K, Cahill A, Odibo L, Macones G, Odibo A. 425: Multi-vessel Doppler parameters for the prediction of adverse neonatal outcome in the preterm growth-restricted fetus: does gestational age matter? American Journal of Obstetrics & Gynecology 2013;208(1):S186. DOI: 10.1016/j.ajog.2012.10.591

150.	Guzman E, Vintzileos A, Martins M. Relationship between middle cerebral artery velocimetry, computer fetal heart rate assessment and degree of acidemia at birth in intrauterine growth restricted fetuses. American Journal of Obstetrics and Gynecology 1995;172:337. DOI:

151.	Habek D, Hodek B, Herman R, Jugović D, Cerkez Habek J, Salihagić A. Fetal biophysical profile and cerebro-umbilical ratio in assessment of perinatal outcome in growth-restricted fetuses. Fetal Diagn Ther 2003 Jan-Feb;18(1):12-16. DOI: 10.1159/000066377

152.	Habek D, Salihagić A, Jugović D, Herman R. Doppler cerebro-umbilical ratio and fetal biophysical profile in the assessment of peripartal cardiotocography in growth-retarded fetuses. Fetal Diagn Ther 2007;22(6):452-456. DOI: 10.1159/000106354

153.	Hata T, Aoki S, Manabe A, Kanenishi K, Yamashiro C, Tanaka H, Yanagihara T. Subclassification of small-for-gestational-age fetus using fetal Doppler velocimetry. Gynecol Obstet Invest 2000;49(4):236-239. DOI: 10.1159/000010266

154.	Hernandez-Andrade E, Figueroa-Diesel H, Jansson T, Rangel-Nava H, Gratacos E. Changes in regional fetal cerebral blood flow perfusion in relation to hemodynamic deterioration in severely growth-restricted fetuses. Ultrasound Obstet Gynecol 2008 Jul;32(1):71-76. DOI: 10.1002/uog.5377

155.	Hershkovitz R, Kingdom JC, Geary M, Rodeck CH. Fetal cerebral blood flow redistribution in late gestation: identification of compromise in small fetuses with normal umbilical artery Doppler. Ultrasound Obstet Gynecol 2000 Mar;15(3):209-212. DOI: 10.1046/j.1469-0705.2000.00079.x

156.	Ivanovski MJ, Lazarevski S, Popovic M. Middle cerebral artery flow velocity waveforms in prediction of adverse outcome in intrauterine growth retarded fetuses. Gynaecologia et Perinatologia 2005 07/01;14:133-139. DOI:

157.	Jugović D, Tumbri J, Medić M, Jukić MK, Kurjak A, Arbeille P, Salihagić-Kadić A. New Doppler index for prediction of perinatal brain damage in growth-restricted and hypoxic fetuses. Ultrasound Obstet Gynecol 2007 Sep;30(3):303-311. DOI: 10.1002/uog.4094

158.	Kalafat E, Ozturk E, Sivanathan J, Thilaganathan B, Khalil A. Longitudinal change in cerebroplacental ratio in small-for-gestational-age fetuses and risk of stillbirth. Ultrasound Obstet Gynecol 2019 Oct;54(4):492-499. DOI: 10.1002/uog.20193

159.	Karlsen HO, Ebbing C, Rasmussen S, Kiserud T, Johnsen SL. Use of conditional centiles of middle cerebral artery pulsatility index and cerebroplacental ratio in the prediction of adverse perinatal outcomes. Acta Obstet Gynecol Scand 2016 Jun;95(6):690-696. DOI: 10.1111/aogs.12912

160.	Lakhkar BN, Rajagopal KV, Gourisankar PT. Doppler prediction of adverse perinatal outcome in PIH and IUGR. Indian Journal of Radiology and Imaging 2006 02/01;16. DOI: 10.4103/0971-3026.29064

161.	Madazli R, Uludağ S, Ocak V. Doppler assessment of umbilical artery, thoracic aorta and middle cerebral artery in the management of pregnancies with growth restriction. Acta Obstet Gynecol Scand 2001 Aug;80(8):702-707. DOI: 10.1034/j.1600-0412.2001.080008702.x

162.	Makhseed M, Jirous J, Ahmed MA, Viswanathan DL. Middle cerebral artery to umbilical artery resistance index ratio in the prediction of neonatal outcome. Int J Gynaecol Obstet 2000 Nov;71(2):119-125. DOI: 10.1016/s0020-7292(00)00262-9

163.	Manogura AC, Turan O, Kush ML, Berg C, Bhide A, Turan S, Moyano D, Bower S, Nicolaides KH, Galan HL, Müller T, Thilaganathan B, Gembruch U, Harman CR, Baschat AA. Predictors of necrotizing enterocolitis in preterm growth-restricted neonates. Am J Obstet Gynecol 2008 Jun;198(6):638.e631-635. DOI: 10.1016/j.ajog.2007.11.048

164.	Marchi L, Pisani M, Simeone S, et al. Markers of adverse feto-neonatal outcomes in late-iugr. Journal of Maternal-Fetal and Neonatal Medicine 2014;27:124-125. DOI:

165.	Mari G, Balasubramaniam M, Nien J, Kusanovic J, Espinoza J, Goncalves L, Soto E, Santolaya-Forgas J, Treadwell M. OC20.03: Middle cerebral artery peak systolic velocity—a new Doppler parameter in the evaluation of SGA fetuses with an abnormal umbilical artery. Ultrasound in Obstetrics & Gynecology 2005 09/01;26:342-342. DOI: 10.1002/uog.2115

166.	Mari G, Deter RL. Middle cerebral artery flow velocity waveforms in normal and small-for-gestational-age fetuses. Am J Obstet Gynecol 1992 Apr;166(4):1262-1270. DOI: 10.1016/s0002-9378(11)90620-6

167.	Mari G, Hanif F, Kruger M, Cosmi E, Santolaya-Forgas J, Treadwell MC. Middle cerebral artery peak systolic velocity: a new Doppler parameter in the assessment of growth-restricted fetuses. Ultrasound Obstet Gynecol 2007 Mar;29(3):310-316. DOI: 10.1002/uog.3953

168.	Marsoosi V, Bahadori F, Esfahani F, Ghasemi-Rad M. The role of Doppler indices in predicting intra ventricular hemorrhage and perinatal mortality in fetal growth restriction. Med Ultrason 2012 Jun;14(2):125-132. DOI:

169.	Maunu J, Ekholm E, Parkkola R, Palo P, Rikalainen H, Lapinleimu H, Haataja L, Lehtonen L. Antenatal Doppler measurements and early brain injury in very low birth weight infants. J Pediatr 2007 Jan;150(1):51-56.e51. DOI: 10.1016/j.jpeds.2006.10.057

170.	Miyashita S, Chiba Y. Doppler Studies Can Predict Long-Term Outcome of Growth-Restricted Fetuses. Journal of Medical Ultrasound 2002 12/31;10:86-93. DOI: 10.1016/S0929-6441(09)60027-8

171.	Murata S, Nakata M, Sumie M, Sugino N. The Doppler cerebroplacental ratio predicts non-reassuring fetal status in intrauterine growth restricted fetuses at term. J Obstet Gynaecol Res 2011 Oct;37(10):1433-1437. DOI: 10.1111/j.1447-0756.2011.01563.x

172.	Nanthakomon T, Uerpairojkit B. Outcome of small-for-gestational-age fetuses according to umbilical artery Doppler: is there any yield from additional middle cerebral artery Doppler? J Matern Fetal Neonatal Med 2010 Aug;23(8):900-905. DOI: 10.3109/14767050903353208

173.	Odibo AO, Goetzinger KR, Cahill AG, Odibo L, Macones GA. Combined sonographic testing index and prediction of adverse outcome in preterm fetal growth restriction. Am J Perinatol 2014 Feb;31(2):139-144. PMID: PMC3932308

174.	Odibo AO, Riddick C, Pare E, Stamilio DM, Macones GA. Cerebroplacental Doppler ratio and adverse perinatal outcomes in intrauterine growth restriction: evaluating the impact of using gestational age-specific reference values. J Ultrasound Med 2005 Sep;24(9):1223-1228. DOI: 10.7863/jum.2005.24.9.1223

175.	Oros D, Garcia-Simon R, Savchev S, Ernesto F, Figueras F. P13.12: Labour induction in late onset intrauterine growth restriction according to cerebral Doppler and cervical conditions. Ultrasound in Obstetrics & Gynecology 2014;44:257-257. DOI:

176.	Ozcan T, Sbracia M, d'Ancona RL, Copel JA, Mari G. Arterial and venous Doppler velocimetry in the severely growth-restricted fetus and associations with adverse perinatal outcome. Ultrasound Obstet Gynecol 1998 Jul;12(1):39-44. DOI: 10.1046/j.1469-0705.1998.12010039.x

177.	Parra-Saavedra M, Crovetto F, Triunfo S, Savchev S, Parra G, Sanz M, Gratacos E, Figueras F. Added value of umbilical vein flow as a predictor of perinatal outcome in term small-for-gestational-age fetuses. Ultrasound Obstet Gynecol 2013 Aug;42(2):189-195. DOI: 10.1002/uog.12380

178.	Peguero A, Eixarch E, Fernández-Blanco L, Oltra M, Paules C, Miranda J, Mazarico E, Gómez Roig M, Gratacós E, Figueras F. EP13.02: Fetal growth centile and Doppler parameters for prediction of poor outcome in second trimester intrauterine growth restriction (IUGR). Ultrasound in Obstetrics & Gynecology 2016;48(S1):320-321. DOI: <https://doi.org/10.1002/uog.16966>

179.	Rhee E, Detti L, Mari G. Superior mesenteric artery flow velocity waveforms in small for gestational age fetuses. J Matern Fetal Med 1998 May-Jun;7(3):120-123. DOI: 10.1002/(sici)1520-6661(199805/06)7:3<120::Aid-mfm4>3.0.Co;2-m

180.	Roy A, Mukherjee S, Bhattacharyya SK, Banerjee P, Das B, Patra KK. Perinatal outcome in pregnancies with intra-uterine growth restriction by using umbilical and middle cerebral artery colour Doppler. J Indian Med Assoc 2012 Mar;110(3):154-157, 163. DOI:

181.	Shaheen S BI, Ahmad I, Singh A. Doppler cerebroplacental ratio and adverse perinatal outcome. Journal of SAFOG 2014;6(1):25-27. DOI:

182.	Shahinaj R, Bare T. Predictive value of abnormal doppler velocimetry in VLBW fetuses with preeclampsia. Journal of Perinatal Medicine 2015;43:65. DOI:

183.	Simeone S, Marchi L, Canarutto R, Pina Rambaldi M, Serena C, Servienti C, Mecacci F. Doppler velocimetry and adverse outcome in labor induction for late IUGR. J Matern Fetal Neonatal Med 2017 Feb;30(3):323-328. DOI: 10.3109/14767058.2016.1171839

184.	Spinillo A, Gardella B, Bariselli S, Alfei A, Silini EM, Bello BD. Cerebroplacental Doppler ratio and placental histopathological features in pregnancies complicated by fetal growth restriction. J Perinat Med 2014 May;42(3):321-328. DOI: 10.1515/jpm-2013-0128

185.	Spinillo A, Montanari L, Roccio M, Zanchi S, Tzialla C, Stronati M. Prognostic significance of the interaction between abnormal umbilical and middle cerebral artery Doppler velocimetry in pregnancies complicated by fetal growth restriction. Acta Obstet Gynecol Scand 2009;88(2):159-166. DOI: 10.1080/00016340802632358

186.	Starčević M, Predojević M, Butorac D, Tumbri J, Konjevoda P, Kadić AS. Early functional and morphological brain disturbances in late-onset intrauterine growth restriction. Early Hum Dev 2016 Feb;93:33-38. DOI: 10.1016/j.earlhumdev.2015.12.001

187.	Sterne G, Shields LE, Dubinsky TJ. Abnormal fetal cerebral and umbilical Doppler measurements in fetuses with intrauterine growth restriction predicts the severity of perinatal morbidity. J Clin Ultrasound 2001 Mar-Apr;29(3):146-151. DOI: 10.1002/1097-0096(200103/04)29:3<146::aid-jcu1014>3.0.co;2-i

188.	Thiebaugeorges O, Ancel PY, Goffinet F, Bréart G. A population-based study of 518 very preterm neonates from high-risk pregnancies: prognostic value of umbilical and cerebral artery Doppler velocimetry for mortality before discharge and severe neurological morbidity. Eur J Obstet Gynecol Reprod Biol 2006 Sep-Oct;128(1-2):69-76. DOI: 10.1016/j.ejogrb.2006.03.015

189.	To WW, Chan AM, Mok KM. Use of umbilical-cerebral Doppler ratios in predicting fetal growth restriction in near-term fetuses. Aust N Z J Obstet Gynaecol 2005 Apr;45(2):130-136. DOI: 10.1111/j.1479-828X.2005.00361.x

190.	Turan OM, Turan S, Berg C, Gembruch U, Nicolaides KH, Harman CR, Baschat AA. Duration of persistent abnormal ductus venosus flow and its impact on perinatal outcome in fetal growth restriction. Ultrasound Obstet Gynecol 2011 Sep;38(3):295-302. DOI: 10.1002/uog.9011

191.	Turan S, Turan OM, Berg C, Moyano D, Bhide A, Bower S, Thilaganathan B, Gembruch U, Nicolaides K, Harman C, Baschat AA. Computerized fetal heart rate analysis, Doppler ultrasound and biophysical profile score in the prediction of acid-base status of growth-restricted fetuses. Ultrasound Obstet Gynecol 2007 Oct;30(5):750-756. DOI: 10.1002/uog.4101

192.	Unterscheider J, Daly S, Geary MP, Kennelly MM, McAuliffe FM, O'Donoghue K, Hunter A, Morrison JJ, Burke G, Dicker P, Tully EC, Malone FD. Predictable progressive Doppler deterioration in IUGR: does it really exist? Am J Obstet Gynecol 2013 Dec;209(6):539.e531-537. DOI: 10.1016/j.ajog.2013.08.039

193.	Vergani P, Roncaglia N, Locatelli A, Andreotti C, Crippa I, Pezzullo JC, Ghidini A. Antenatal predictors of neonatal outcome in fetal growth restriction with absent end-diastolic flow in the umbilical artery. Am J Obstet Gynecol 2005 Sep;193(3 Pt 2):1213-1218. DOI: 10.1016/j.ajog.2005.07.032

194.	Warshak CR, Masters H, Regan J, DeFranco E. Doppler for growth restriction: the association between the cerebroplacental ratio and a reduced interval to delivery. J Perinatol 2015 May;35(5):332-337. DOI: 10.1038/jp.2014.211

195.	Westby Eger SH, Kessler J, Kiserud T, Markestad T, Sommerfelt K. Foetal Doppler abnormality is associated with increased risk of sepsis and necrotising enterocolitis in preterm infants. Acta Paediatr 2015 Apr;104(4):368-376. DOI: 10.1111/apa.12893

196.	Yoshimura S, Masuzaki H, Miura K, Gotoh H, Ishimaru T. Fetal blood flow redistribution in term intrauterine growth retardation (IUGR) and post-natal growth. Int J Gynaecol Obstet 1998 Jan;60(1):3-8. DOI: 10.1016/s0020-7292(97)00212-9

197.	Fox NS, Chasen ST. First trimester pregnancy associated plasma protein-A as a marker for poor pregnancy outcome in patients with early-onset fetal growth restriction. Prenat Diagn 2009 Dec;29(13):1244-1248. DOI: 10.1002/pd.2397

198.	Persico N, D'Ambrosi F, Fabietti I, Boito S, Aiello E, Bulfoni A, Ciralli F, Kustermann A, Mosca F, Fedele L. Fetal Doppler changes 1week after endoscopic equatorial laser for twin-to-twin transfusion syndrome: A longitudinal study. PRENATAL DIAGNOSIS 2018 APR;38(5):344-348. DOI: 10.1002/pd.5234

199.	Baschat A, Harman C, Robinson J, Galan HL. 621: How should the growth restricted (FGR) fetus be monitored before delivery? Results of a national survey. American Journal of Obstetrics & Gynecology 2007;197(6):S180. DOI: 10.1016/j.ajog.2007.10.648

200.	Monaghan C, Binder J, Thilaganathan B, Morales-Roselló J, Khalil A. Perinatal loss at term: role of uteroplacental and fetal Doppler assessment. Ultrasound Obstet Gynecol 2018 Jul;52(1):72-77. DOI: 10.1002/uog.17500

201.	Twomey S, Flatley C, Kumar S. The association between a low cerebro-umbilical ratio at 30-34 weeks gestation, increased intrapartum operative intervention and adverse perinatal outcomes. Eur J Obstet Gynecol Reprod Biol 2016 Aug;203:89-93. DOI: 10.1016/j.ejogrb.2016.05.036

202.	D'Antonio F, Rijo C, Thilaganathan B, Akolekar R, Khalil A, Papageourgiou A, Bhide A. Association between first-trimester maternal serum pregnancy-associated plasma protein-A and obstetric complications. Prenat Diagn 2013 Sep;33(9):839-847. DOI: 10.1002/pd.4141

203.	Kirkegaard I, Henriksen TB, Tørring N, Uldbjerg N. PAPP-A and free β-hCG measured prior to 10 weeks is associated with preterm delivery and small-for-gestational-age infants. Prenat Diagn 2011 Feb;31(2):171-175. DOI: 10.1002/pd.2671

204.	Kwik M, Morris J. Association between first trimester maternal serum pregnancy associated plasma protein-A and adverse pregnancy outcome. Aust N Z J Obstet Gynaecol 2003 Dec;43(6):438-442. DOI: 10.1046/j.0004-8666.2003.00126.x

205.	Marttala J, Peuhkurinen S, Laitinen P, Gissler M, Nieminen P, Ryynanen M. Low maternal PAPP-A is associated with small-for-gestational age newborns and stillbirths. Acta Obstet Gynecol Scand 2010 Sep;89(9):1226-1228. DOI: 10.3109/00016349.2010.493195

206.	Montanari L, Alfei A, Albonico G, Moratti R, Arossa A, Beneventi F, Spinillo A. The impact of first-trimester serum free beta-human chorionic gonadotropin and pregnancy-associated plasma protein A on the diagnosis of fetal growth restriction and small for gestational age infant. Fetal Diagn Ther 2009;25(1):130-135. DOI: 10.1159/000207554

207.	Patil M, Panchanadikar TM, Wagh G. Variation of papp-a level in the first trimester of pregnancy and its clinical outcome. J Obstet Gynaecol India 2014 Apr;64(2):116-119. PMID: PMC3984652

208.	Ranta JK, Raatikainen K, Romppanen J, Pulkki K, Heinonen S. Decreased PAPP-A is associated with preeclampsia, premature delivery and small for gestational age infants but not with placental abruption. Eur J Obstet Gynecol Reprod Biol 2011 Jul;157(1):48-52. DOI: 10.1016/j.ejogrb.2011.03.004

209.	Saruhan Z, Ozekinci M, Simsek M, Mendilcioglu I. Association of first trimester low PAPP-A levels with adverse pregnancy outcomes. Clin Exp Obstet Gynecol 2012;39(2):225-228. DOI:

210.	Sanchez-Fernandez M, Corral ME, Aceituno L, Mazheika M, Mendoza N, Mozas-Moreno J. Observer Influence with Other Variables on the Accuracy of Ultrasound Estimation of Fetal Weight at Term. MEDICINA-LITHUANIA 2021 MAR;57(3). DOI: 10.3390/medicina57030216

211.	Jörn H, Funk A, Fendel H. [Doppler ultrasound diagnosis in post-term pregnancy]. Geburtshilfe Frauenheilkd 1993 Sep;53(9):603-608. DOI: 10.1055/s-2007-1023595

212.	Latif HA, Gaafar HM, Moety GA, Mahmoud DS, El Rifai NM. Brain Volume and Doppler Velocimetry in Growth-Restricted, Small-for-Gestational-Age, and Appropriate-for-Gestational-Age Fetuses. Am J Perinatol 2017 Mar;34(4):333-339. DOI: 10.1055/s-0036-1586752

213.	Melamed N, Pittini A, Kingdom J, Barrett J. 163: Sonographic factors distinguishing late intrauterine growth restriction from late small for gestational age fetuses. American Journal of Obstetrics & Gynecology 2016;214(1):S104-S105. DOI: 10.1016/j.ajog.2015.10.199

214.	Rizzo G, Capponi A, Arduini D, Romanini C. Ductus venosus velocity waveforms in appropriate and small for gestational age fetuses. Early Hum Dev 1994 Sep 30;39(1):15-26. DOI: 10.1016/0378-3782(94)90066-3

215.	Allotey J, Fernandez-Felix BM, Zamora J, Moss N, Bagary M, Kelso A, Khan R, van der Post JAM, Mol BW, Pirie AM, McCorry D, Khan KS, Thangaratinam S. Predicting seizures in pregnant women with epilepsy: Development and external validation of a prognostic model. PLoS Med 2019 May;16(5):e1002802. PMID: PMC6513048 following competing interests: AMP has been paid to provide medico-legal reports on the standard of care of women with epilepsy in pregnancy and use of anti-epileptic drugs, and has jointly held grants from the NIHR and Epilepsy Action for research on epilepsy in pregnancy. The other authors have declared that no competing interests exist.

216.	Binder J, Kalafat E, Palmrich P, Pateisky P, Khalil A. Angiogenic markers and their longitudinal change for predicting adverse outcomes in pregnant women with chronic hypertension. Am J Obstet Gynecol 2021 Apr 1. DOI: 10.1016/j.ajog.2021.03.041

217.	Graupner O, Helfrich F, Ostermayer E, Lobmaier SM, Ortiz JU, Ewert P, Wacker-Gussmann A, Haller B, Axt-Fliedner R, Enzensberger C, Abel K, Karge A, Oberhoffer R, Kuschel B. Application of the INTERGROWTH-21st chart compared to customized growth charts in fetuses with left heart obstruction: late trimester biometry, cerebroplacental hemodynamics and perinatal outcome. Arch Gynecol Obstet 2019 Sep;300(3):601-613. DOI: 10.1007/s00404-019-05198-6

218.	Grewal J, Siu SC, D'Souza R, Lee T, Singer J, Rychel V, Kiess M, Sermer M, Silversides CK. Cardiac Risk Score to Predict Small-for-Gestational-Age Infants in Pregnant Women with Heart Disease. Can J Cardiol 2021 Apr 8. DOI: 10.1016/j.cjca.2021.03.023

219.	Henry A, Gopikrishna S, Mahajan A, Alphonse J, Meriki N, Welsh AW. Use of the Foetal Myocardial Performance Index in monochorionic, diamniotic twin pregnancy: a prospective cohort and nested case-control study. J Matern Fetal Neonatal Med 2019 Jun;32(12):2017-2029. DOI: 10.1080/14767058.2018.1424817

220.	Kushner T, Sarkar M, Tran T. Noninvasive Tests for Prognosticating Outcomes in Patients With Chronic Liver Disease in Pregnancy: Ready for Prime Time? American Journal of Gastroenterology 2019;114(2):209-211. DOI: 10.14309/ajg.0000000000000101

221.	Leavitt K, Odibo L, Nwosu O, Odibo AO. Comparing the cerebro-placental to umbilico-cerebral Doppler ratios for the prediction of adverse neonatal outcomes in pregnancies complicated by fetal growth restriction. JOURNAL OF MATERNAL-FETAL & NEONATAL MEDICINE 2021. DOI: 10.1080/14767058.2021.1901880

222.	Mackie FL, Hall MJ, Morris RK, Kilby MD. Early prognostic factors of outcomes in monochorionic twin pregnancy: systematic review and meta-analysis. Am J Obstet Gynecol 2018 Nov;219(5):436-446. DOI: 10.1016/j.ajog.2018.05.008

223.	Mackie FL, Whittle R, Morris RK, Hyett J, Riley RD, Kilby MD. First-trimester ultrasound measurements and maternal serum biomarkers as prognostic factors in monochorionic twins: a cohort study. Diagn Progn Res 2019;3:9. PMID: PMC6507122

224.	Majak GB, Reisæter AV, Zucknick M, Lorentzen B, Vangen S, Henriksen T, Michelsen TM. Preeclampsia in kidney transplanted women; Outcomes and a simple prognostic risk score system. PLoS One 2017;12(3):e0173420. PMID: PMC5358770

225.	Ozgen G, Cakmak BD, Dundar B, Tasgoz FN, Bayram F, Karadag B. Is pregnancy associated plasma protein-A (PAPP-A) a marker for adverse perinatal outcomes in preterm isolated oligohydramnios cases? TAIWANESE JOURNAL OF OBSTETRICS & GYNECOLOGY 2018 FEB;57(1):71-75. DOI: 10.1016/j.tjog.2017.12.012

226.	Robertson JA, Kimble RM, Stockton K, Sekar R. Antenatal ultrasound features in fetuses with gastroschisis and its prediction in neonatal outcome. Aust N Z J Obstet Gynaecol 2017 Feb;57(1):52-56. DOI: 10.1111/ajo.12565

227.	Swarray-Deen A, Nkyekyer K, Seffah JD, Mumuni K, Mensah-Brown SA, Tuuli MG, Oppong SA. Cerebro-placental ratio as a prognostic factor of fetal outcome in pregnancy complicated by maternal sickle cell disease. Int J Gynaecol Obstet 2020 Aug;150(2):248-253. DOI: 10.1002/ijgo.13196

228.	Walter IJ, Haneveld MJK, Lely AT, Bloemenkamp KWM, Limper M, Kooiman J. Pregnancy outcome predictors in antiphospholipid syndrome: A systematic review and meta-analysis. Autoimmun Rev 2021 Jul 16:102901. DOI: 10.1016/j.autrev.2021.102901

229.	Adiga P, Kantharaja I, Hebbar S, Rai L, Guruvare S, Mundkur A. Predictive value of middle cerebral artery to uterine artery pulsatility index ratio in hypertensive disorders of pregnancy. Int J Reprod Med 2015;2015:614747. PMID: PMC4333592

230.	Arbeille P, Maulik D, Stree JL, Fignon A, Amyel C, Deufel M. Fetal cerebral and renal Doppler in small for gestational age fetuses in hypertensive pregnancies. Eur J Obstet Gynecol Reprod Biol 1994 Aug;56(2):111-116. DOI: 10.1016/0028-2243(94)90266-6

231.	Dubiel M, Breborowicz GH, Marsal K, Gudmundsson S. Fetal adrenal and middle cerebral artery Doppler velocimetry in high-risk pregnancy. Ultrasound Obstet Gynecol 2000 Oct;16(5):414-418. DOI: 10.1046/j.1469-0705.2000.00278.x

232.	Dubiel M, Gunnarsson GO, Gudmundsson S. Blood redistribution in the fetal brain during chronic hypoxia. Ultrasound Obstet Gynecol 2002 Aug;20(2):117-121. DOI: 10.1046/j.1469-0705.2002.00758.x

233.	Flores Acosta C, Saldívar Rodríguez D, Nuñez Albar R, Treviño Martinez G, Maldonado J, Sepulveda Gonzalez G. P24.10: Cerebroplacental Doppler Index in severe preeclampsia. Ultrasound in Obstetrics & Gynecology 2010;36(S1):261-262. DOI: <https://doi.org/10.1002/uog.8638>

234.	Gibbons A, Flatley C, Kumar S. Cerebroplacental ratio in pregnancies complicated by gestational diabetes mellitus. Ultrasound Obstet Gynecol 2017 Aug;50(2):200-206. DOI: 10.1002/uog.17242

235.	Lalthantluanga C, Devi N, Singh N, Shugeta N, Khuman V, Keishing S. Study on role of obstetrical Doppler in pregnancies with hypertensive disorders of pregnancy. Journal of Medical Society 2015 May 1, 2015;29(2):79-82. DOI: 10.4103/0972-4958.163195

236.	Orabona R, Gerosa V, Gregorini ME, Pagani G, Prefumo F, Valcamonico A, Frusca T. The prognostic role of various indices and ratios of Doppler velocimetry in patients with pre-eclampsia. Clin Exp Hypertens 2015;37(1):57-62. DOI: 10.3109/10641963.2014.897723

237.	Pagani G, Gerosa V, Gregorini ME, Rovida PL, Prefumo F, Valcamonico A, Frusca T, Andrea L. PP110. The role of doppler to predict adverse pregnancy outcome in patients with pre-eclampsia. Pregnancy Hypertens 2012 Jul;2(3):298-299. DOI: 10.1016/j.preghy.2012.04.221

238.	Shahinaj R, Manoku N, Kroi E, Tasha I. The value of the middle cerebral to umbilical artery Doppler ratio in the prediction of neonatal outcome in patient with preeclampsia and gestational hypertension. J Prenat Med 2010 Apr;4(2):17-21. PMID: PMC3279170

239.	Singh M, Sharma A, Singh P. ROLE OF DOPPLER INDICES IN THE PREDICTION OF ADVERSE PERINATAL OUTCOME IN PREECLAMPSIA. The Journal of medical research 2013;3:315-318. DOI:

240.	Souza AS, Amorim MM, Vasconcelos-Neto MJ, Oliveira-Filho JR, Sousa-Júnior FA. [Factors associated with fetal brain-sparing effect in patients with hypertension in pregnancy]. Rev Bras Ginecol Obstet 2013 Jul;35(7):309-316. DOI: 10.1590/s0100-72032013000700005

241.	Souza AS, Vasconcelos Neto MJ, Cunha AS, Monteiro Ede F, Amorim MM. [Comparison of Doppler indexes to predict small infants for gestational age in pregnant women with hypertensive syndromes]. Rev Bras Ginecol Obstet 2011 Apr;33(4):157-163. DOI:

242.	Swarray-Deen A. OP21.05: The cerebroplacental ratio as a prognostic factor of fetal outcome in pregnancies complicated by maternal sickle cell disease. Ultrasound in Obstetrics & Gynecology 2017;50:11V 117. DOI:

243.	Urban R, Lemancewicz A, Urban J, Alifier M, Kretowska M. [The Doppler cerebroplacental ratios and perinatal outcome in post-term pregnancy]. Ginekol Pol 2000 Apr;71(4):317-321. DOI:

244.	Yalti S, Oral O, Gürbüz B, Ozden S, Atar F. Ratio of middle cerebral to umbilical artery blood velocity in preeclamptic & hypertensive women in the prediction of poor perinatal outcome. Indian J Med Res 2004 Jul;120(1):44-50. DOI:

245.	Zavala-Coca C, Pacora P. Abnormal cerebral-placental ratio identifies 60% of newborns with severe neonatal morbidity in pregnancies complicated by severe preeclampsia. American Journal of Obstetrics & Gynecology 2003;189(6):S181. DOI: 10.1016/j.ajog.2003.10.445

246.	Allotey J, Snell KIE, Smuk M, Hooper R, Chan CL, Ahmed A, Chappell LC, von Dadelszen P, Dodds J, Green M, Kenny L, Khalil A, Khan KS, Mol BW, Myers J, Poston L, Thilaganathan B, Staff AC, Smith GCS, Ganzevoort W, Laivuori H, Odibo AO, Ramirez JA, Kingdom J, Daskalakis G, Farrar D, Baschat AA, Seed PT, Prefumo F, Costa FD, Groen H, Audibert F, Masse J, Skrastad RB, Salvesen KA, Haavaldsen C, Nagata C, Rumbold AR, Heinonen S, Askie LM, Smits LJM, Vinter CA, Magnus PM, Eero K, Villa PM, Jenum AK, Andersen LB, Norman JE, Ohkuchi A, Eskild A, Bhattacharya S, McAuliffe FM, Galindo A, Herraiz I, Carbillon L, Klipstein-Grobusch K, Yeo S, Teede HJ, Browne JL, Moons KGM, Riley RD, Thangaratinam S, Network IC. Validation and development of models using clinical, biochemical and ultrasound markers for predicting pre-eclampsia: an individual participant data meta-analysis. HEALTH TECHNOLOGY ASSESSMENT 2020 DEC;24(72):I-+. DOI: 10.3310/hta24720

247.	Belza C, Fitzgerald K, de Silva N, Avitzur Y, Wales PW. Early Predictors of Enteral Autonomy in Pediatric Intestinal Failure Resulting From Short Bowel Syndrome: Development of a Disease Severity Scoring Tool. JPEN J Parenter Enteral Nutr 2019 Nov;43(8):961-969. DOI: 10.1002/jpen.1691

248.	Caughey AB. Prediction of Stillbirth: An Umbrella Review of Evaluation of Prognostic Variables. OBSTETRICAL & GYNECOLOGICAL SURVEY 2021 JUN;76(6):315-317. DOI: 10.1097/OGX.0000000000000937

249.	Choudhry S, Salter A, Cunningham TW, Levy PT, Hackett BP, Singh GK, Johnson MC. Risk factors and prognostic significance of altered left ventricular geometry in preterm infants. J Perinatol 2018 May;38(5):543-549. DOI: 10.1038/s41372-018-0047-5

250.	El Ayoubi M, Jarreau PH, Van Reempts P, Cuttini M, Kaminski M, Zeitlin J. Does the antenatal detection of fetal growth restriction (FGR) have a prognostic value for mortality and short-term morbidity for very preterm infants? Results from the MOSAIC cohort. Journal of Maternal-Fetal and Neonatal Medicine 2016;29(4):596-601. DOI: 10.3109/14767058.2015.1012062

251.	Eysbouts YK, Ottevanger PB, Massuger L, IntHout J, Short D, Harvey R, Kaur B, Sebire NJ, Sarwar N, Sweep F, Seckl MJ. Can the FIGO 2000 scoring system for gestational trophoblastic neoplasia be simplified? A new retrospective analysis from a nationwide dataset. Ann Oncol 2017 Aug 1;28(8):1856-1861. PMID: PMC5834141

252.	Fuchs F, Borrego P, Amouroux C, Antoine B, Ollivier M, Faure JM, Lopez C, Forgues D, Faure A, Merrot T, Boulot P, Jeandel C, Philibert P, Gaspari L, Sultan C, Paris F, Kalfa N. Prenatal imaging of genital defects: clinical spectrum and predictive factors for severe forms. BJU Int 2019 Nov;124(5):876-882. DOI: 10.1111/bju.14714

253.	Ghaemi MS, Tarca AL, Romero R, Stanley N, Fallahzadeh R, Tanada A, Culos A, Ando K, Han XY, Blumenfeld YJ, Druzin ML, El-Sayed YY, Gibbs RS, Winn VD, Contrepois K, Ling XFB, Wong RJ, Shaw GM, Stevenson DK, Gaudilliere B, Aghaeepour N, Angst MS. Proteomic signatures predict preeclampsia in individual cohorts but not across cohorts - implications for clinical biomarker studies. JOURNAL OF MATERNAL-FETAL & NEONATAL MEDICINE 2021. DOI: 10.1080/14767058.2021.1888915

254.	Houweling TAJ, van Klaveren D, Das S, Azad K, Tripathy P, Manandhar D, Neuman M, de Jonge E, Been JV, Steyerberg E, Costello A. A prediction model for neonatal mortality in low- and middle-income countries: an analysis of data from population surveillance sites in India, Nepal and Bangladesh. INTERNATIONAL JOURNAL OF EPIDEMIOLOGY 2019 FEB;48(1):186-198. DOI: 10.1093/ije/dyy194

255.	Hukkinen M, Kivisaari R, Merras-Salmio L, Koivusalo A, Pakarinen MP. Small Bowel Dilatation Predicts Prolonged Parenteral Nutrition and Decreased Survival in Pediatric Short Bowel Syndrome. Annals of Surgery 2017;266(2):369-375. DOI: 10.1097/SLA.0000000000001893

256.	Samuels N, van de Graaf RA, de Jonge RCJ, Reiss IKM, Vermeulen MJ. Risk factors for necrotizing enterocolitis in neonates: a systematic review of prognostic studies. BMC Pediatr 2017 Apr 14;17(1):105. PMID: PMC5391569

257.	Sokou R, Piovani D, Konstantinidi A, Tsantes AG, Parastatidou S, Lampridou M, Ioakeimidis G, Gounaris A, Iacovidou N, Kriebardis AG, Politou M, Kopterides P, Bonovas S, Tsantes AE. A Risk Score for Predicting the Incidence of Hemorrhage in Critically Ill Neonates: Development and Validation Study. Thrombosis and Haemostasis 2021;121(2):131-139. DOI: 10.1055/s-0040-1715832

258.	Srinivasan S, Treacy R, Herrero T, Olsen R, Leonardo TR, Zhang X, DeHoff P, To C, Poling LG, Fernando A, Leon-Garcia S, Knepper K, Tran V, Meads M, Tasarz J, Vuppala A, Park S, Laurent CD, Bui T, Cheah PS, Overcash RT, Ramos GA, Roeder H, Ghiran I, Parast M, Breakefield XO, Lueth AJ, Rust SR, Dufford MT, Fox AC, Hickok DE, Burchard J, Boniface JJ, Laurent LC. Discovery and Verification of Extracellular miRNA Biomarkers for Non-invasive Prediction of Pre-eclampsia in Asymptomatic Women. Cell Rep Med 2020 May 19;1(2). PMID: PMC7455024

259.	Stock SJ, Bhide A, Richardson H, Black M, Yuill C, Harkness M, Reid M, Wee F, Cheyne H, McCourt C, Rana D, Boyd KA, Sanders J, Heera N, Huddleston J, Denison F, Pasupathy D, Modi N, Smith G, Norrie J. Cervical ripening at home or in-hospital-prospective cohort study and process evaluation (CHOICE) study: a protocol. BMJ Open 2021 May 4;11(5):e050452. PMID: PMC8098973

260.	Townsend R, Sileo FG, Allotey J, Dodds J, Heazell A, Jorgensen L, Kim VB, Magee L, Mol B, Sandall J, Smith GCS, Thilaganathan B, von Dadelszen P, Thangaratinam S, Khalil A. Prediction of stillbirth: an umbrella review of evaluation of prognostic variables. BJOG: An International Journal of Obstetrics and Gynaecology 2021;128(2):238-250. DOI: 10.1111/1471-0528.16510

261.	Usta CS, Atik TK, Ozcaglayan R, Bulbul CB, Camili FE, Adali E. Does the fibrinogen/albumin ratio predict the prognosis of pregnancies with abortus imminens? Saudi Medical Journal 2021;42(3):255-263. DOI: 10.15537/SMJ.2021.42.3.20200695

262.	Akolekar R, Syngelaki A, Gallo DM, Poon LC, Nicolaides KH. Umbilical and fetal middle cerebral artery Doppler at 35-37 weeks' gestation in the prediction of adverse perinatal outcome. Ultrasound Obstet Gynecol 2015 Jul;46(1):82-92. DOI: 10.1002/uog.14842

263.	Allam IS, Abuelghar W, Fathy H, Enin LA, Sayed M. Prediction of neonatal acidosis by ductus venosus Doppler pattern in high risk pregnancies. Middle East Fertility Society Journal 2013 2013/03/01/;18(1):47-52. DOI: <https://doi.org/10.1016/j.mefs.2012.10.003>

264.	Cheema R, Dubiel M, Gudmundsson S. Signs of fetal brain sparing are not related to umbilical cord blood gases at birth. Early Hum Dev 2009 Jul;85(7):467-470. DOI: 10.1016/j.earlhumdev.2009.04.003

265.	Crovetto F, Baffero G, Cesano N, Rossi F, Acerboni S, De Marinis S, Fedele L, Acaia B, Persico N. OP31.08: Fetal middle cerebral artery Doppler at 40 weeks' gestation in the prediction of emergency delivery for fetal distress in labour. Ultrasound in Obstetrics & Gynecology 2016;48(S1):156-156. DOI: <https://doi.org/10.1002/uog.16463>

266.	Dajić BC, Vilendecić R. [Diagnostic efficacy of biophysical tests and cerebral-umbilical index when assessing fetal oxygenation]. Med Pregl 2013 Jul-Aug;66(7-8):292-296. DOI: 10.2298/mpns1308292c

267.	D'Antonio F, Patel D, Chandrasekharan N, Thilaganathan B, Bhide A. Role of cerebroplacental ratio for fetal assessment in prolonged pregnancy. Ultrasound Obstet Gynecol 2013 Aug;42(2):196-200. DOI: 10.1002/uog.12357

268.	Devine PA, Bracero LA, Lysikiewicz A, Evans R, Womack S, Byrne DW. Middle cerebral to umbilical artery Doppler ratio in post-date pregnancies. Obstet Gynecol 1994 Nov;84(5):856-860. DOI:

269.	Duncan J, Tobiasz A, Schenone M, Thompson R, G M. P01.10: The cerebroplacental ratio does not appear to predict severe neonatal outcomes in preterm premature rupture of membranes. Ultrasound in Obstetrics & Gynecology 2017;50(S1):157-. DOI:

270.	Eser A, Zulfıkaroglu E, Eserdag S, Kılıc S, Danısman N. Predictive value of middle cerebral artery to uterine artery pulsatility index ratio in preeclampsia. Arch Gynecol Obstet 2011 Aug;284(2):307-311. DOI: 10.1007/s00404-010-1660-5

271.	Guerrero Casillas MA, Romero Gutiérrez G, Molina Rodríguez R, Guzán Mena G. [Correlation between Doppler fluxometry of middle cerebral artery/umbilical and non stress test as methods of antepartum fetal surveillance]. Ginecol Obstet Mex 2007 Apr;75(4):193-199. DOI:

272.	Gupta U, Chandra S, Narula M. Value of middle cerebral artery to umbilcal artery ratio by doppler velocimetry in pregnancies beyond term. J Obstet Gynaecol India 2006;56(1):37-40. DOI:

273.	Hernandez-Andrade EA, Ahn H, Garcia M, Tarca A, Saker H, Yeo L, Korzeniewski S, Hassan S, Romero R. OC15.07: A reduced cerebral placental ratio in fetuses at risk of preterm delivery is associated with neonatal sepsis and necrotising enterocolitis (NEC). Ultrasound in Obstetrics & Gynecology 2015 09/01;46:33-34. DOI: 10.1002/uog.15048

274.	Hoffmann H, Chaoui R, Bollmann R, Metzner A. [Evaluation of the central hemodynamics of the fetus using pulsed Doppler ultrasound]. Zentralbl Gynakol 1990;112(11):673-678. DOI:

275.	Kant A, Seth N, Rastogi D. Comparison of Outcome of Normal and High-Risk Pregnancies Based Upon Cerebroplacental Ratio Assessed by Doppler Studies. J Obstet Gynaecol India 2017 Jun;67(3):173-177. PMID: PMC5425638

276.	Maeda M, Nomura R, Niigaki J, Liao A, Zugaib M. Accuracy of fetal cerebroplacental ratio to discriminate cases with acidemia at birth (PH<7.15) in placental dysfunction. J Perinat Med 2011;39:100. DOI:

277.	Maeda Mde F, Nomura RM, Niigaki JI, Miyadahira S, Zugaib M. [Cerebroplacental ratio and acidemia to the birth in placental insufficiency detected before 34th week's gestation]. Rev Bras Ginecol Obstet 2010 Oct;32(10):510-515. DOI:

278.	Mari G, Abuhamad AZ, Keller M, et al. Is the fetal brain-sparing effect a risk factor for the development of intraventricular hemorrhage in the preterm infant? American Journal of Obstetrics & Gynecology 1995 Nov;172(1):268. DOI:

279.	Ott WJ. Comparison of the non-stress test with the evaluation of centralization of blood flow for the prediction of neonatal compromise. Ultrasound Obstet Gynecol 1999 Jul;14(1):38-41. DOI: 10.1046/j.1469-0705.1999.14010038.x

280.	Ott WJ. Middle cerebral artery blood flow in the fetus and central nervous system complications in the neonate. J Matern Fetal Neonatal Med 2003 Jul;14(1):26-29. DOI: 10.1080/jmf.14.1.26.29

281.	Prior T, Mullins E, Bennett P, Kumar S. Prediction of intrapartum fetal compromise using the cerebroumbilical ratio: a prospective observational study. Am J Obstet Gynecol 2013 Feb;208(2):124.e121-126. DOI: 10.1016/j.ajog.2012.11.016

282.	Prior T, Paramasivam G, Bennett P, Kumar S. Are fetuses that fail to achieve their growth potential at increased risk of intrapartum compromise? Ultrasound Obstet Gynecol 2015 Oct;46(4):460-464. DOI: 10.1002/uog.14758

283.	Regan J, Masters H, Warshak CR. Association between an abnormal cerebroplacental ratio and the development of severe pre-eclampsia. J Perinatol 2015 May;35(5):322-327. DOI: 10.1038/jp.2014.210

284.	Sabdia S, Greer RM, Prior T, Kumar S. Predicting intrapartum fetal compromise using the fetal cerebro-umbilical ratio. Placenta 2015 May;36(5):594-598. DOI: 10.1016/j.placenta.2015.01.200

285.	Shannon A, Crimmins S, Kopelman J, Harman C, O T. Abnormal MCA dopplers in diabetic patients and the association with stillbirth. Reprod Sci 2017;24(1):250A. DOI:

286.	Shyken J, Lieberman S, Kivikoski A, Smeltzer J. 202 Low Middle Cerebral Artery (MCA) Resistance Index of Pourcelot (Ri) Predicts Neonatal Morbidity in Post-Term Pregnancies. American Journal of Obstetrics & Gynecology 1992;166(1):334. DOI: 10.1016/S0002-9378(12)91368-X

287.	Stoicescu S, Damian O, Ciobanu D, Paicu S, Ianisevskaia O. Clinical considerations and ultrasound examination at premature infants with gestational age less than 32 weeks. Obstetrica si Ginecologie 2013;61(3):159-165. DOI:

288.	Vahanian S, Kinzler WL, Chen A, Saleh I, Chavez M, Vintzileos AM. 745: Fetal surveillance using middle cerebral artery (MCA) Dopplers in pregnancies complicated by diabetes. American Journal of Obstetrics & Gynecology 2016;214(1):S390-S391. DOI: 10.1016/j.ajog.2015.10.793

289.	Weiss E, Ulrich S, Berle P. Blood flow velocity waveforms of the middle cerebral artery and abnormal neurological evaluations in live-born fetuses with absent or reverse end-diastolic flow velocities of the umbilical arteries. Eur J Obstet Gynecol Reprod Biol 1992 Jul 3;45(2):93-100. DOI: 10.1016/0028-2243(92)90223-l

290.	Zhang YN. [Clinical evaluation of fetal blood flow velocity in cerebral artery and umbilical artery by color Doppler imaging]. Zhonghua Fu Chan Ke Za Zhi 1993 Jul;28(7):395-396, 440-391. DOI:

291.	Dane B, Dane C, Batmaz G, Ates S, Dansuk R. First trimester maternal serum pregnancy-associated plasma protein-A is a predictive factor for early preterm delivery in normotensive pregnancies. Gynecol Endocrinol 2013 Jun;29(6):592-595. DOI: 10.3109/09513590.2013.788626

292.	Dugoff L, Hobbins JC, Malone FD, Porter TF, Luthy D, Comstock CH, Hankins G, Berkowitz RL, Merkatz I, Craigo SD, Timor-Tritsch IE, Carr SR, Wolfe HM, Vidaver J, D'Alton ME. First-trimester maternal serum PAPP-A and free-beta subunit human chorionic gonadotropin concentrations and nuchal translucency are associated with obstetric complications: a population-based screening study (the FASTER Trial). Am J Obstet Gynecol 2004 Oct;191(4):1446-1451. DOI: 10.1016/j.ajog.2004.06.052

293.	Jelliffe-Pawlowski LL, Shaw GM, Currier RJ, Stevenson DK, Baer RJ, O'Brodovich HM, Gould JB. Association of early-preterm birth with abnormal levels of routinely collected first- and second-trimester biomarkers. Am J Obstet Gynecol 2013 Jun;208(6):492.e491-411. PMID: PMC3672244

294.	She BQ, Chen SC, Lee FK, Cheong ML, Tsai MS. Low maternal serum levels of pregnancy-associated plasma protein-A during the first trimester are associated with subsequent preterm delivery with preterm premature rupture of membranes. Taiwan J Obstet Gynecol 2007 Sep;46(3):242-247. DOI: 10.1016/s1028-4559(08)60027-3

295.	Giardini V, Allievi S, Fornari C, Rovelli R, Cesana G, Lafranconi A, Vergani P. Management of pregnancy blood pressure increase in the emergency room: role of PlGF-based biochemical markers and relative economic impact. JOURNAL OF MATERNAL-FETAL & NEONATAL MEDICINE 2021 APR 3;34(7):1083-1090. DOI: 10.1080/14767058.2019.1624718
